# Estimating the prevalence of discrepancies between study registrations and publications: A systematic review and meta-analyses

**DOI:** 10.1101/2021.07.07.21259868

**Authors:** TARG Meta-Research Group & Collaborators, Robert T Thibault, Robbie Clark, Hugo Pedder, Olmo van den Akker, Samuel Westwood, Jacqueline Thompson, Marcus Munafo

## Abstract

**Objectives:** Prospectively registering study plans in a permanent time-stamped and publicly accessible document is becoming more common across disciplines and aims to reduce risk of bias and make risk of bias transparent. Selective reporting persists, however, when researchers deviate from their registered plans without disclosure. This systematic review aimed to estimate the prevalence of undisclosed discrepancies between prospectively registered study plans and their associated publication. We further aimed to identify the research disciplines where these discrepancies have been observed, whether interventions to reduce discrepancies have been conducted, and gaps in the literature.

**Design:** Systematic review and meta-analyses.

**Data sources:** Scopus and Web of Knowledge, published up to 15 December 2019.

**Eligibility criteria:** Articles that included quantitative data about discrepancies between registrations or study protocols and their associated publications.

**Data extraction and synthesis:** Each included article was independently coded by two reviewers using a coding form designed for this review (osf.io/728ys). We used random-effects meta-analyses to synthesize the results.

**Results:** We reviewed k = 89 articles, which included k = 70 that reported on primary outcome discrepancies from n = 6314 studies and, k = 22 that reported on secondary outcome discrepancies from n = 1436 studies. Meta-analyses indicated that between 29% to 37% (95% confidence interval) of studies contained at least one primary outcome discrepancy and between 50% to 75% (95% confidence interval) contained at least one secondary outcome discrepancy. Almost all articles assessed clinical literature, and there was considerable heterogeneity. We identified only one article that attempted to correct discrepancies.

**Conclusions:** Many articles did not include information on whether discrepancies were disclosed, which version of a registration they compared publications to, and whether the registration was prospective. Thus, our estimates represent discrepancies broadly, rather than our target of *undisclosed* discrepancies between *prospectively* registered study plans and their associated publications. Discrepancies are common and reduce the trustworthiness of medical research. Interventions to reduce discrepancies could prove valuable.

**Registration:** osf.io/ktmdg. Protocol amendments are listed in Supplementary Material A.

## Introduction

In 2000, ClinicalTrials.gov and the ISRCTN Registry were launched with several aims, including aiding participant recruitment, facilitating knowledge synthesis, and reducing duplication, publication bias and selective reporting (Deborah A. Zarin et al., 2017). In 2005, the International Committee of Medical Journal Editors (ICMJE) made *prospective* registration a condition of consideration for publication (De Angelis et al., 2004). Thousands of journals now claim to follow this policy (ICMJE, 2021). In parallel, the World Health Organization International Clinical Trials Registry Platform established a minimum set of required information for a trial to be considered fully registered, including experimental design elements such as the conditions being studied, intervention, key inclusion and exclusion criteria, sample size, primary outcomes, and key secondary outcomes (Sim et al., 2006). While the relatively widespread uptake of clinical trial registration has substantially improved transparency, many trials remain unregistered, are registered after enrollment of participants begins or analyses are complete (i.e., *retrospective* registration), are never published, or publish outcomes discrepant with those in the registration without disclosing the discrepancy (e.g., Chan et al., 2017; Scott et al., 2015). Nevertheless, the existence of registries allows researchers to identify and quantify these issues.

In research disciplines other than clinical trials, study registration is becoming more common, but remains far from standard practice (Hardwicke et al., 2021; Hardwicke, Wallach, et al., 2020; Scoggins & Robertson, 2023; Thibault et al., 2023). For example, starting around 2011 the field of psychology has increasingly taken the “replication crisis” seriously and many researchers and journals now use registration to reduce bias and make risk of bias transparent. Other disciplines have created dedicated registries, such as PROSPERO for systematic reviews and the American Economic Association RCT Trial Registry.

In this manuscript, we systematically reviewed articles that quantify the prevalence of discrepancies between registrations or study protocols and their associated publications (e.g., in primary outcome measures). Our analysis extended beyond the three systematic reviews already published on this topic in several ways (Dwan, Gamble, et al., 2013; C. W. Jones et al., 2015; G. Li et al., 2018). First, registration has expanded beyond clinical trials; we included all research disciplines and used key word searches for registries including the Open Science Framework, the American Economic Association RCT Trial Registry, and PROSPERO. Second, we extracted more fine-grained information about a wide range of discrepancies (e.g., outcomes, analysis, sample size), as well as which version of the registration was surveyed and whether discrepancies were disclosed (we believe disclosed discrepancies present little reason for concern). Third, our review included over twice as many studies as previous systematic reviews on this topic, provides meta-analytic estimates, and used meta-regression and additional analyses to attempt to identify predictors of discrepancies.

## Methods

### Terminology

We present a systematic review of k = 89 articles that assessed a wide range of outcome discrepancies and non-outcome discrepancies across over n = 7,000 studies. To avoid confusion, this report consistently uses the terms *studies* to refer to the over n = 7,000 individual studies that were *assessed*, and the term *article* to refer to the k = 89 articles that assessed these studies, and that we *reviewed*. We restrict our usage of the term *publication* to refer to the publications stemming from the *studies* (not to refer to the *articles*).

We use the term *discrepancy* to refer to any incongruity between the content of a publication and its associated registration (e.g., on clinicaltrials.gov) or study protocol (e.g., submitted to an ethics review board or funding agency)—see Box 1 for examples. We use the term *prospective registration* broadly to include terms used in different research disciplines, such as prospective trial registration, preregistration, and pre-analysis plans. All these terms indicate the registration of study details *before* commencing a study, or in some cases, before viewing the data or removing the blind. They are in contrast to *retrospective* registration, which occurs after participant enrollment begins or analyses are complete. We use the term outcome discrepancy to indicate a discrepancy in the outcome measure registered versus the outcome measure reported in a publication (not to indicate a discrepancy in the value of a reported outcome between these documents).

##### Box 1. Examples of discrepancies

We coded 10 types of outcome discrepancies (listed in Table 2) and 10 types of non-outcome discrepancies (listed in Table 4). The degree to which the information in a registration is discrepant with the information in a publication can range widely*^a^*. The associated concern about risk of bias can also range widely and often requires domain expertise to assess. We present two examples from Calméjane et al. (2018) in this box. Many more examples are available in the appendix of Calméjane et al. (2018) and at https://www.compare-trials.org/results.

**Registered**: Primary outcome: Visual Acuity [Time Frame: 6 months].

**Published**: Primary outcome: 3-year cumulative incidence rate of myopia. Myopia was defined as a spherical equivalent refractive error (sphere +½ cylinder) of at least -0.50 D

**Coded as**: Timing of outcome measurement changed.

**Registered**: Primary outcome: Severity of Device and Procedure related complications [Time Frame: At the time of ExAblate Transcranial thalamotomy procedure]. Secondary outcome: Effectiveness of the ExAblate Transcranial MRgFUS treatment determined using the Clinical Rating Scale for Tremor (CRST) [Time Frame: Participants will be followed from the date of treatment until study completion, approximately up to 12 months].

**Published**: Primary outcome: Change from baseline to 3 months in the tremor score for the hand derived from the CRST, Part A (three items: resting, postural, and action or intention components of hand tremor), and the CRST, Part B (five tasks involving handwriting, drawing, and pouring).

**Coded as**: Secondary outcome promoted to primary outcome; Timing of outcome measurement changed; Primary outcome omitted.

a Some researchers suggest that people checking for discrepancies should compare publications to clinical trial study protocols instead of registrations (Annals of Internal Medicine Editors, 2016). Study protocols are generally far more thorough and are more likely to be updated when trialists change their plans (which may occur before starting a study, but after registering). Study protocols contain timestamps, yet they often become public only at the time of publication.

### Eligibility criteria

We included articles that reported quantitative data about discrepancies between registrations or study protocols and their associated publication. We excluded conference proceedings and articles written in a language other than English (for full inclusion and exclusion criteria, see our preregistered protocol).

### Study selection

We searched Scopus and Web of Science on 15 December 2019 using the queries in Appendix A and Appendix B of our preregistered protocol (osf.io/ktmdg). Briefly, our queries included (1) variations of the terms preregistration, pre-analysis plans, and prospective registration in the title or keyword fields; (2) terms indicating discrepancies such as “outcome switching” in the title, keywords or abstract; (3) names of registration or protocol repositories such as “clinicaltrials.gov” in the title or keywords; and excluded overlapping but irrelevant terms (e.g., “nursing preregistration”). To limit the number of irrelevant articles, we did not search for variations of the term preregistration or for repository names in the abstract field.

Our search returned 4,283 articles after duplicates were removed (see Figure 1 for a PRISMA flowchart). Articles were screened independently by two reviewers in two stages. In Stage One, reviewers screened titles and, if necessary, briefly examined abstracts of articles to determine inclusion in the systematic review or in a scoping review on prospective registration^1^. If at least one of the reviewers deemed an article potentially relevant, it was included in Stage Two screening. In Stage Two, the reviewers independently examined the remaining 464 abstracts in greater detail for eligibility. Disagreements were resolved through discussion between the two reviewers and eventual consensus. Inter-rater agreemend for the 464 articles was Cohen’s *k* = 0.67 for inclusion in the systematic review (the list of articles and coding is available at osf.io/wa62f). Interrater agreement for all 4,283 articles was Cohen’s *k* = 0.72. We then used a snowball method and identified 33 additional articles that met our inclusion criteria, mostly through citations in G. Li et al. (2018) and C. W. Jones et al. (2015). These 33 additional articles are not included in the inter-rater agreement scores. After After full-text review, we included 89 articles in our systematic review.

**Figure 1.**
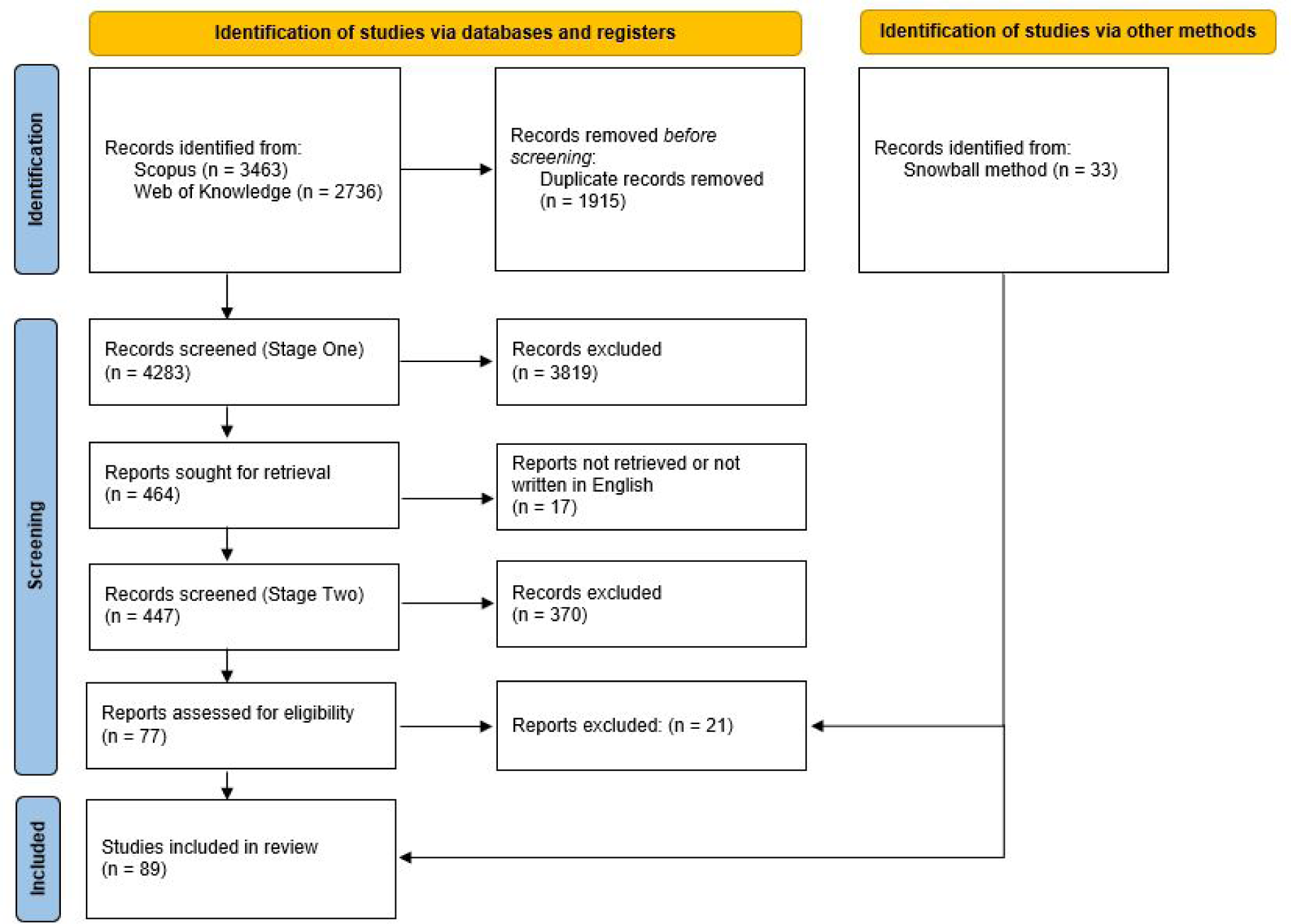
PRISMA flowchart of article inclusion

### Coding items

Each included article was independently coded by two of four reviewers (RTT, RC, OvdA, SW) using a coding form designed for this review. The form consisted of five sections that assessed (1) article characteristics, (2) study registration details, (3) 10 types of outcome discrepancies (listed in Table 2), (4) 10 types of non-outcome discrepancies (listed in Table 4), and (5) any additional descriptive or inferential statistics on discrepancies. The form details the operationalization of each variable we coded, and is available at osf.io/728ys. We chose items to code based on a pilot test of our protocol, as well as the categories used in a seminal paper (Chan, Hr’objartsson, et al., 2004) and a systematic review on discrepancies (G. Li et al., 2018).

The data we extracted was often presented as summary results in a table and sometimes as text in the results section. To be included in our meta-analyses this data had to include at least two of (1) the number of studies assessed (denominator), (2) the number of studies with a given discrepancy (numerator), and (3) the percentage of studies with a given discrepancy—from which we could calculate an unreported numerator or denominator. We did not access the raw data. If an article did not report data for a certain measure (e.g., secondary outcome discrepancies), then we did not include that article in the meta-analysis for that measure (this is why k varies among the meta-analyses we present—see Tables 2 and 4). For the meta-regressions we performed, we could not find data on (1) the version of the registry that publications were compared to for 35 articles, and (2) the number of studies that disclosed discrepancies for 57 articles. We coded these cases as “not reported” and included “not reported” as a factor in the meta-regressions. All other meta-regression data was complete. Further coding details are available in Supplementary Material B.

The complete dataset, including the coding of each reviewer and the resolved coding, is available at osf.io/ue2c6. A cleaned dataset with only the resolved coding is available at osf.io/6cn9m.

### Statistical analyses

We performed two main random effects meta-analyses: one on the proportion of studies with at least one primary outcome discrepancy, and another on the proportion of studies with at least one secondary outcome discrepancy. We used random effects models because they allow for the true effect to vary across the populations the articles sampled from, and the articles we reviewed differ in their methodologies and the research disciplines that they assess. We used a random intercept logistic regression model with the Knapp-Hartung adjustment for the synthesis of proportions (J. Hartung & Knapp, 2003). We used the maximum-likelihood method for estimating the between-study heterogeneity (Tau). We also performed meta-regressions to test whether article characteristics are associated with the proportion of studies with at least one primary or secondary outcome discrepancy.

For pooled estimates, we report both confidence intervals and prediction intervals. Whereas researchers are likely more familiar with confidence intervals, interpreting confidence intervals can be unintuitive (Hoekstra et al., 2014), and their pooled-estimate does not incorporate uncertainty due to the between-article heterogeneity. If we assume that we could resample from our population, 95% of the resampled *meta-analyses* would yield a 95% confidence interval that contains the true value of the parameter being estimated (e.g., proportion of articles with at least one primary outcome discrepancy). Alternatively, if we are interested in the results that would come from another *article* assessing discrepancies, we would want a 95% prediction interval. In other words, of 100 articles drawn from the same population, we could expect the results from 95 of them—on average—to fall within the 95% prediction interval. While prediction intervals are not commonly reported, methodologists recommend reporting them for random effects meta-analysis, particularly when few articles are included or, as in our case, included articles are highly heterogeneous (Higgins et al., 2009; Riley et al., 2011).

Whereas we did not perform a formal risk of bias assessment—because our review differed substantially from the purpose these tools were built for—we shed light on a few potential sources of bias with additional analyses that consider the funding source, statistical significance, and the timing of registration of included *studies*. These additional analyses were not prospectively registered. We made a few amendments to our preregistered study protocol which are listed in Supplementary Material A.

## Results

### Articles characteristics

We identified and reviewed k = 89 articles that report at least one type of discrepancy. Articles that checked for outcome discrepancies assessed a median of 68 studies (IQR: 33 to 112). Article characteristics are outlined in Table 1. All articles except for two, one preprint in economics (Ofosu & Posner, 2019) and one preprint in psychology (Claesen et al., 2019), focused on clinical trials or systematic reviews. All but k = 10 articles were solely observational. Only one article attempted to correct published discrepancies (Goldacre et al., 2019). The authors assessed all trials published in five journals over a six-week period and sent a letter to the editor for each trial that published a discrepant outcome (for 58 letters in total). In the time since our literature search, at least two more interventional studies were published. One reports a trial that attempted to reduce discrepancies at medical journals by sending peer reviewers information about the study registration (C. W. Jones et al., 2022). They found null results. The other was a feasibility study that assigned a peer reviewer to specifically check for discrepancies in manuscripts submitted for publication (TARG Meta-Research Group & Collaborators, 2022). Further details about article characteristics are available in Supplementary Material C.

**Table 1.** Article characteristics

| Article characteristic | k = 89 | n = 6929 |
| --- | --- | --- |
| **Discipline** |  |  |
| medicine | 81 | 6452 |
| dentistry | 3 | 254 |
| psychology | 3 | 68 |
| economics | 1 | 93 |
| physical therapy | 1 | 62 |
| **Source of registration or protocol assessed for discrepancies** |  |  |
| registry | 73 | 6107 |
| ethics application | 7 | 146 |
| other protocol | 5 | 466 |
| marketing application | 2 | 126 |
| grant application | 2 | 84 |
| **Sources searched to identify studies** |  |  |
| journals | 32 | 3264 |
| registries | 23 | 1547 |
| search engines | 16 | 1527 |
| ethics boards | 7 | 146 |
| funders | 3 | 96 |
| registries and search engines | 3 | 27 |
| regulators | 2 | 126 |
| research group | 2 | 44 |
| registries and journals | 1 | 152 |
| **Type of study** |  |  |
| solely observational | 79 | 6344 |
| observational and study authors were contacted | 9 | 518 |
| observational and interventional | 1 | 67 |
| **Version of registry that publications were compared to** |  |  |
| original | 29 | 2607 |
| most recent | 15 | 1281 |
| other version/unclear | 10 | 670 |
| not reported | 35 | 2371 |
| **Number of studies within each article that disclose discrepancies** |  |  |
| one or more | 19 | 1547 |
| none | 9 | 441 |
| article excluded publications with disclosed discrepancies | 4 | 326 |
| not reported | 57 | 4615 |

#### Registration timing

Articles varied in the level of detail they provided about whether and when studies were registered. For example, whereas some articles presented their sample only after selecting for prospectively registered studies, other articles detailed their selection process including how many studies were registered and if so, when they were registered. Using the terminology in the articles we reviewed, articles identified studies that were registered retrospectively (k = 29), registered during participant enrollment (k = 17), registered after participant enrollment was complete (k = 14), and studies that were not registered (k = 36)^2^. Many articles were ambiguous regarding when some studies were registered (k = 47) and whether or not some studies were registered at all (k = 24). While these data do not provide fine-grained detail, they highlight two overarching issues: many studies are not registered, and many registered studies are registered retrospectively. These studies fail to meet the Declaration of Helsinki (World Medical Association, 2013) (item 35) requirement that “Every research study involving human subjects must be registered in a publicly accessible database before recruitment of the first subject” and the equivalent International Committee of Medical Journal Editors (ICMJE) policy (ICMJE, 2019), which thousands of journals claim to follow (ICMJE, 2021).

Eighty of the k = 89 articles we reviewed report at least one type of outcome discrepancy. Of these, 23 report only on studies that were unambiguously prospectively registered, 51 do not unambiguously distinguish between prospectively and retrospectively registered studies, and 6 report outcome discrepancies separately for each of prospectively and retrospectively registered studies. Separate meta-analyses for unambiguously prospectively registered studies and studies with unclear timing of registration are presented in Supplementary Material D.

Forty-six of the k = 89 articles report at least one non-outcome discrepancy (e.g., in sample size or analyses). Of these, 12 report only on studies that were unambiguously prospectively registered, 33 do not unambiguously distinguish between prospectively and retrospectively registered studies, and one reports non-outcome discrepancies separately for each of prospectively and retrospectively registered studies.

### Primary outcome discrepancies

An estimated 29-37% (95% confidence interval) of the population of studies contained at least one primary outcome discrepancy (Figure 2). This equates to a 95% prediction interval of 10-68%.

**Figure 2.**
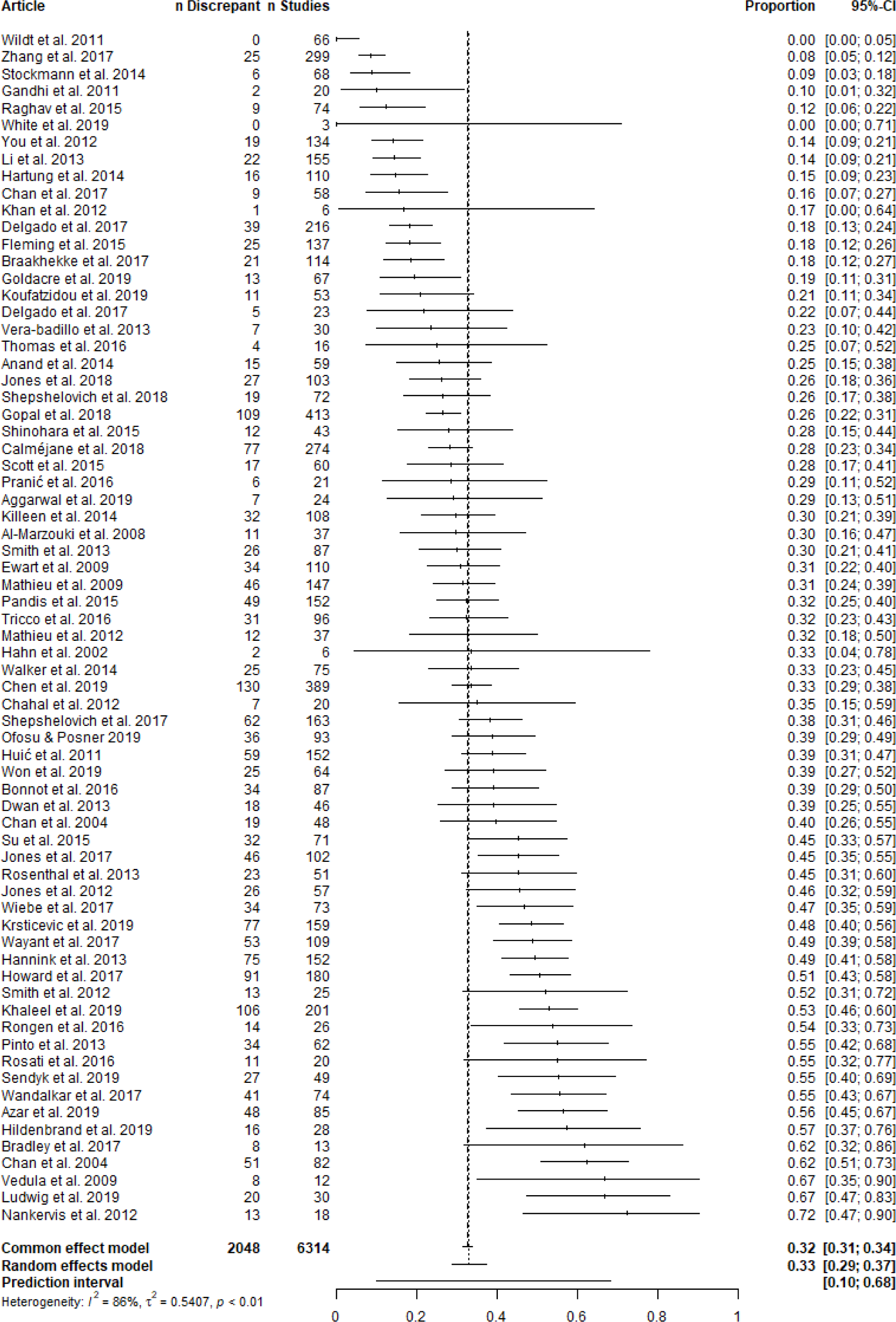
Forest plot of articles reporting the proportion of assessed studies with at least one primary

This meta-analysis had high heterogeneity (*I* ^2^ = 86%), suggesting that the broad range of estimates across the articles stem largely from differences in the methodology of the articles or populations they sample from, rather than from chance. Heterogeneity could not be explained by meta-regression of any of the following article-level characteristics: discipline (p = 0.28)^3^, whether the publications were compared to registry entries versus other protocol formats (e.g., ethics applications) (p = 0.46), sources searched to identify studies (p = 0.65), version of the registry analyzed (p = 0.77), whether discrepancies were disclosed (p = 0.97), and year of article publication (p = 0.83). The meta-regression on discipline had low power because 63 articles assessed medical research and 7 assessed studies across dentistry, psychology, physical therapy, and economics. To increase statistical power, we reran this meta-regression after dichotomizing discipline and found that nonmedical disciplines may have a greater proportion of studies with at least one primary outcome discrepancy (p = 0.09; OR 95% CI: 0.91-3.19). We ran another meta-regression after dichotomizing the source which publications were compared to into registrations versus other protocols and did not find evidence to suggest this moderator played a role (p = 0.42). We also conducted a sensitivity analysis that included all six article-level characteristics in a single meta-regression. We found that publications compared to the most recent version of a registration may have a smaller proportion of studies with at least one primary outcome discrepancy, realtive to publications that were compared to the original version of a registration (p = 0.09; OR 95% CI: 0.32-1.07). All meta-regression model summaries are presented in Supplementary Material E.

The high heterogeneity in this meta-analysis may stem from genuine differences among the articles, including the sub-disciplines surveyed, specific sources searched, definition of a discrepancy (e.g., whereas some articles considered a change in the timing of an outcome as a discrepancy, others did not), and other article characteristics that may or may not have been reported. Our dataset contains more fine-grained information about the specific sub-discipline surveyed and specific sources searched. While we do not further explore these potential moderators in the present report, we note that, whereas some sub-disciplines and sources were highly specific (e.g., cystic fibrosis, lung cancer immunotherapy, Global Resource of Eczema Trials database), others were broad (e.g., medicine, clinicaltrials.gov, core clinical MEDLINE journals). We did not collect information on the exact definitions an article used to identify a primary outcome discrepancy. However, we did collect information on the proportion of articles with sub-categories of outcome discrepancies, which are more strictly defined and listed in Table 2 (e.g., promoting a secondary outcome to a primary outcome). We ran meta-analyses on these sub-categories of outcome discrepancies and found they also had high heterogeneity (Table 2). Thus, varying definitions are unlikely to be the main driver of the high heterogeneity in the present analysis on primary outcome discrepancies.

**Table 2.** Meta-analytic estimates for the proportion of studies that contain various types of outcome discrepancies

|  | Discrepant studies (95% CI) | Discrepant studies (95% PI) | k | n |
| --- | --- | --- | --- | --- |
| Any outcome discrepancy | 41-75% | 7-97% | 18 | 1113 |
| Any primary outcome discrepancy | 29-37% | 10-68% | 70 | 6314 |
| Any secondary outcome discrepancy | 50-75% | 13-95% | 22 | 1436 |
| Primary outcome demoted to secondary outcome | 6-10% | 2-31% | 51 | 4560 |
| Primary outcome omitted | 6-12% | 1-43% | 51 | 4338 |
| Primary outcome added | 7-11% | 2-34% | 54 | 4697 |
| Secondary outcome promoted to primary outcome | 4-6% | 1-19% | 46 | 4135 |
| Secondary outcome omitted | 16-35% | 4-72% | 18 | 1243 |
| Secondary outcome added | 19-43% | 3-83% | 20 | 1305 |
| Timing of outcome measurement changed | 6-16% | 1-62% | 30 | 3472 |

We only coded 'any outcome discrepancy' for articles that checked for both primary and secondary outcome discrepancies in the studies they assessed. Forest plots for all meta-analyses in this table are in Supplementary Material F.

### Secondary outcome discrepancies

An estimated 50-75% (95% confidence interval) of the population of studies contained at least one secondary outcome discrepancy (Figure 3). This equates to a 95% prediction interval of 13-95%.

**Figure 3.**
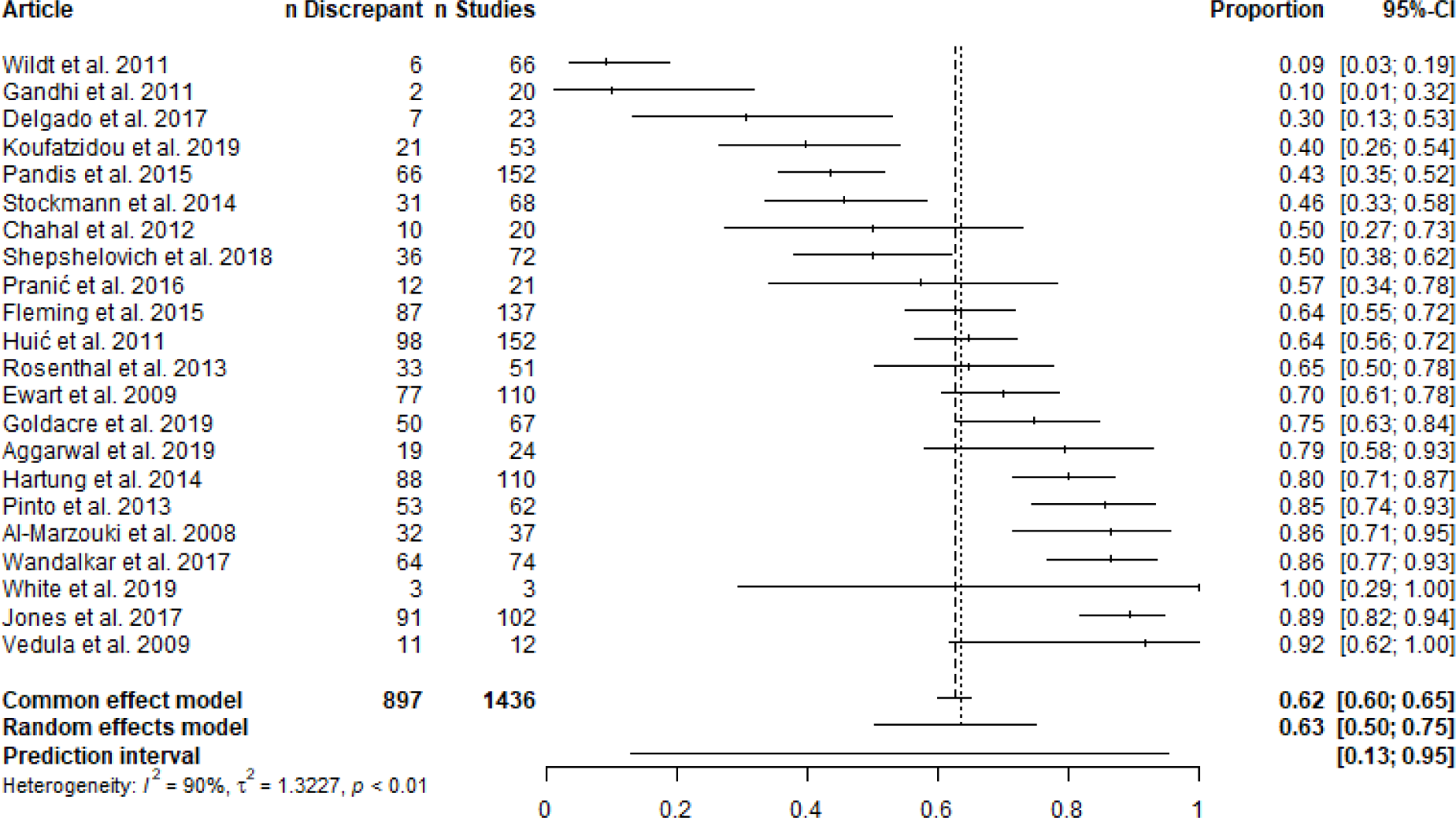
Forest plot of articles reporting the proportion of assessed studies with at least one secondary outcome discrepancy.

This meta-analysis also had high heterogeneity (*I* ^2^ = 90%) which could not be explained by meta-regression of the version of the registry analyzed (p = 0.8) or the year of article publication (p = 0.72). Metaregression of the sources searched to identify studies explained some heterogeneity, in that searches stemming from journals, compared to registries, had a greater proportion of publications with at least one secondary outcome discrepancy (p = 0.03; OR 95% CI: 1.89-13.45). Meta-regressions on discipline (p = 0.29), whether discrepancies were disclosed (p = 0.68), and whether the publications were compared to registry entries versus other protocol formats (p = 0.08) had very low statistical power because almost all articles had the same characteristic. All meta-regression model summaries are included in Supplementary Material E.

Descriptively, omitting secondary outcomes and adding secondary outcomes appears to occur more frequently than omitting primary outcomes, adding primary outcomes, or demoting primary outcomes, which in turn appear to occur more frequently than promoting a secondary outcome (see Table 2).

### Parameters potentially related to outcome discrepancies

A subset of articles contained information on parameters potentially related to the proportion of outcome discrepancies. These include the disclosure of discrepancies, presence of a ‘statistically significant’ result, funding source, and timing of registration (Table 3).

**Table 3.** Additional analyses regarding discrepancies

| Analysis | 95% CI | 95% PI | k | n |
| --- | --- | --- | --- | --- |
| Percentage of studies with at least one outcome discrepancy that disclose an outcome discrepancy | 4-19% | 0.3-74% | 21 | 620 |
| Percentage of outcome discrepancies that favored statistically significant results | 49-66% | 23-86% | 24 | 671 |
| Likelihood ratio of a study with versus without an outcome discrepancy to contain statistically significant results | 0.56-1.06 | 0.42-1.43 | 7 | 405 |
| *Likelihood ratio of a study with versus without a statistically significant results to contain an outcome discrepancy | 0.64-0.99 | 0.59-1.06 | 7 | 405 |
| Likelihood ratio of a study with versus without industry funded to contain an outcome discrepancy | 0.61-0.91 | 0.44-1.27 | 22 | 2623 |
| Likelihood ratio of a prospectively registered study versus retrospectively registered study to contain a primary outcome discrepancy | 0.46-2.57 | 0.13-8.85 | 4 | 260 |

\* This analysis uses the same data as the likelihood ratio analysis before it. Forest plots for all meta-analyses in this table are in Supplementary Material G.

### Non-outcome discrepancies

The meta-analyses for the non-outcome discrepancies had high heterogeneity, and wide confidence intervals and prediction intervals (Table 4). Articles varied in the criteria they used to identify non-outcome discrepancies and there were fewer articles than for outcome discrepancies. Prediction intervals can be particularly imprecise when few articles are included in a meta-analysis (IntHout et al., 2016). Whereas our coding procedure divided outcome discrepancies into 10 sub-categories, it did not employ the same level of granularity for non-outcome discrepancies. Supplementary Material H contains additional information on non-outcome discrepancies.

**Table 4.** Meta-analytic estimates for the proportion of studies that contain various types of non-outcome discrepancies

|  | Discrepant studies (95% CI) | Discrepant studies (95% PI) | k | n |
| --- | --- | --- | --- | --- |
| Eligibility criteria | 25-57% | 5-90% | 15 | 1153 |
| Sample size | 26-44% | 8-78% | 25 | 1398 |
| Randomization | 2-64% | 0.06-98% | 5 | 176 |
| Blinding | 5-42% | 0.04-99% | 3 | 224 |
| Intervention | 3-52% | 0.1-97% | 7 | 550 |
| Study duration | 3-89% | 0.02-99.94% | 4 | 184 |
| Analysis | 19-52% | 4-86% | 12 | 404 |
| Subgroup analysis | 35-93% | 2-99.7% | 9 | 545 |
| Funding | 7-84% | 0.2-99.5% | 5 | 212 |
| Results | 7-82% | 0.2-99% | 6 | 262 |

Values less than 0.5% and greater than 99.5% are rounded to one significant digit from 0 or 100. Forest plots for all meta-analyses in this table are in Supplementary Material I. These results are best interpreted alongside the information provided in Supplementary Material H.

### Gaps in the literature

We identified several gaps in the literature on discrepancies. There exists little research on: (1) the prevalence of discrepancies in fields other than clinical research, (2) the prevalence of discrepancies in a representative sample across clinical disciplines, (3) the level of specificity in registrations, and (4) interventions to reduce undisclosed discrepancies (see Supplementary Material J for additional information about these gaps). We also identified several themes from surveying the conclusions of the articles we reviewed. These include the need for awareness surrounding discrepancies, the need for mandates, enforcement, and/or new initiatives to address discrepancies, and the benefit of registering additional information such as analysis plans (Supplementary Table J1 contains additional details).

## Discussion

We found that outcome measures in registrations and study protocols often differ from published outcome measures, that these discrepancies are rarely disclosed, that the prevalence of discrepancies varies substantially across the articles we reviewed, and that this heterogeneity is not easily assigned to specific article characteristics.

### Limitations

Given the wide range of discrepancy prevalence across individual articles, point estimates and confidence intervals may provide false precision when extrapolating our findings to the registered literature at large. Moreover, because heterogeneity could not be explained by meta-regression of article characteristics, more precise estimates cannot be derived for subsets of the literature. The prediction intervals can reasonably be used to extrapolate to another article in the registered medical literature at large, although the included studies do not necessarily form a representative sample.

### Comparisons to previous research

Our main findings are in line with those from previous systematic reviews. These reviews included 27 articles each and found that 31% of studies had a primary outcome discrepancy in the median article they reviewed (C. W. Jones et al., 2015) and 54% of studies had any outcome discrepancy in the median article they reviewed (G. Li et al., 2018). The latter review did not distinguish between primary and secondary outcomes, and many articles they reviewed only assessed primary outcomes. Our review included all the articles contained in these systematic reviews, except for a few that did not meet our inclusion criteria (e.g., a PhD thesis, an abstract).

### Implications

Our review raises broader issues regarding the efficiency of the research ecosystem and the trustworthiness of research outputs. We identified articles that documented discrepancies between publications and all of registrations, protocols, ethics applications, funding applications, and marketing approval applications. The existence of multiple documents outlining the same study raises the likelihood of discrepancies and, in the absence of a clearly demarcated ‘master’ document, leaves ambiguity regarding which document is ‘correct’. Rehashing the same study details for different audiences may also be an inefficient use of researchers’ time. Identifying a single publicly accessible document as the version of record (this could be the registration) and having all other documents point to this version of record for key information could reduce ambiguity and improve efficiency.

As for trustworthiness, registration has had a clearly positive influence on medical research (DeVito, 2022; Kaplan & Irvin, 2015). At the same time, some registration policies have poor adherence (e.g., many trials are registered retrospectively, and many trial results are never reported (DeVito et al., 2019)). The existence of research policies that are regularly overlooked, rarely monitored, and come with no consequence for non-compliance, can be damaging in at least two ways. They risk devaluing research policies altogether and they can reduce the trustworthiness of research outputs by creating a false impression that rigorous research practices were employed. Conceiving research as a complex ecosystem comprised of various agents with diverse incentives (e.g., funders, publishers, institutions, individual researchers) can help to comprehend why some policies have poor adherence and to develop and implement effective research infrastructure.

## Conclusion

Registrations provide the evidence to detect selective reporting and outcome switching, which we found to be common. Nearly all articles we reviewed focused on documenting issues. Future efforts regarding discrepancies—and research improvement broadly—could prove more fruitful by shifting focus towards developing and testing solutions to these now well-documented issues.

## Supplementary material

osf.io/byqhp

## Transparency statement

Robert Thibault, the manuscript’s guarantor, affirms that the manuscript is an honest, accurate, and transparent account of the study being reported; that no important aspects of the study have been omitted; and that any discrepancies from the study as originally planned have been explained.

Data, data dictionaries, analysis script and materials related to this study are publicly available on the Open Science Framework at https://osf.io/5gfty/. The study protocol and materials were registered on 17 January 2020 at https://osf.io/ktmdg. Discrepancies between this manuscript and the registered protocol are outlined in the Supplementary Material A. To facilitate reproducibility, this manuscript was written by interleaving regular prose and analysis code using R Markdown. The relevant files are available in a Code Ocean container (https://doi.org/10.24433/CO.4753181.v1) which recreates the software environment in which the original analyses were performed. This container allows this manuscript to be reproduced from the data and code with a single button press.

## Contributors

**Conceptualization**: Robert T. Thibault(1,2,†), Jacqueline M. Thompson(2,9), and Marcus R. Munafò(2,3).

**Data curation**: Robert T. Thibault.

**Formal analysis**: Robert T. Thibault and Hugo Pedder(4,5).

**Funding acquisition**: Robert T. Thibault.

**Investigation**: Robert T. Thibault, Robbie Clark(2,3), Olmo van den Akker(6,7), and Samuel Westwood(8).

**Methodology**: Robert T. Thibault, Robbie Clark, and Marcus R. Munafò.

**Project administration**: Robert T. Thibault.

**Software**: Robert T. Thibault and Hugo Pedder.

**Supervision**: Robert T. Thibault and Marcus R. Munafò.

**Validation**: Robert T. Thibault.

**Visualization**: Robert T. Thibault.

**Writing - original draft**: Robert T. Thibault.

**Writing - review & editing**: Robert T. Thibault, Robbie Clark, Hugo Pedder, Olmo van den Akker, Samuel Westwood, Jacqueline M. Thompson, and Marcus R. Munafò.

(1) Meta-Research Innovation Center at Stanford (METRICS), Stanford University
(2) School of Psychological Science, University of Bristol
(3) MRC Integrative Epidemiology Unit at the University of Bristol
(4) Population Health Sciences, University of Bristol
(5) Bristol Medical School, University of Bristol
(6) Department of Methodology & Statistics, Tilburg University
(7) QUEST Center for Responsible Research, Berlin Institute of Health
(8) Department of Psychology, Institute of Psychiatry, Psychology, Neuroscience, King’s College London
(9) Bodleian Libraries, University of Oxford

†Correspondence should be addressed to Robert Thibault

## Footnotes

1 We had planned to report on this scoping review separately. However, the scoping review has been terminated (details provided at https://osf.io/ktmdg).

2 By definition, non-registered studies cannot be assessed for discrepancies between their registration and publication. Several articles identified non-registered studies in their sampling process, but did not include these studies in their final sample.

3 If readers want to find if a study on discrepancies exists within a particular sub-discipline, we invite them to explore the variable ‘disciplineDetail’ in our dataset.

## References

Aggarwal, R., & Oremus, M. (2019). Selective outcome reporting is present in randomized controlled trials in lung cancer immunotherapies. JOURNAL OF CLINICAL EPIDEMIOLOGY, 106, 145–146. https://doi.org/10.1016/j.jclinepi.2018.10.010

Anand, V., Scales, D. C., Parshuram, C. S., & Kavanagh, B. P. (2014). Registration and design alterations of clinical trials in critical care: A cross-sectional observational study. Intensive Care Medicine. https://doi.org/10.1007/s00134-014-3250-7

Annals of Internal Medicine Editors. (2016). Discrepancies Between Prespecified and Reported Outcomes. Annals of Internal Medicine, 164 (5), 374–375. https://doi.org/10.7326/L15-0615

Azar, M., Riehm, K. E., Saadat, N., Sanchez, T., Chiovitti, M., Qi, L., Rice, D. B., Levis, B., Fedoruk, C., Levis, A. W., Kloda, L. A., Kimmelman, J., Benedetti, A., & Thombs, B. D. (2019). Evaluation of Journal Registration Policies and Prospective Registration of Randomized Clinical Trials of Nonregulated Health Care Interventions. JAMA INTERNAL MEDICINE, 179 (5), 624–632. https://doi.org/10.1001/jamainternmed.2018.8009

Bonnot, B., Yavchitz, A., Mantz, J., Paugam-Burtz, C., & Boutron, I. (2016). Selective primary outcome reporting in high-impact journals of anaesthesia and pain. BRITISH JOURNAL OF ANAESTHESIA, 117 (4), 542–543. https://doi.org/10.1093/bja/aew280

Boonacker, C. W. B., Hoes, A. W., Van Liere-Visser, K., Schilder, A. G. M., & Rovers, M. M. (2011). A comparison of subgroup analyses in grant applications and publications. American Journal of Epidemiology. https://doi.org/10.1093/aje/kwr075

Braakhekke, M., Scholten, I., Mol, F., Limpens, J., Mol, B. W., & van der Veen, F. (2017). Selective outcome reporting and sponsorship in randomized controlled trials in IVF and ICSI. HUMAN REPRODUCTION, 32 (10), 2117–2122. https://doi.org/10.1093/humrep/dex273

Bradley, H. A., Rucklidge, J. J., & Mulder, R. T. (2017). A systematic review of trial registration and selective outcome reporting in psychotherapy randomized controlled trials. ACTA PSYCHIATRICA SCANDINAVICA, 135 (1), 65–77. https://doi.org/10.1111/acps.12647

Calm’ejane, L., Dechartres, A., Tran, V. T., & Ravaud, P. (2018). Making protocols available with the article improved evaluation of selective outcome reporting. Journal of Clinical Epidemiology, 104, 95–102. https://doi.org/10.1016/j.jclinepi.2018.08.020

Chahal, J., Tomescu, S. S., Ravi, B., Bach Jr., B. R., Ogilvie-Harris, D., Mohamed, N. N., & Gandhi, R. (2012). Publication of Sports Medicine-Related Randomized Controlled Trials Registered in Clinical-Trials.gov. AMERICAN JOURNAL OF SPORTS MEDICINE, 40 (9), 1970–1977. https://doi.org/10.1177/0363546512448363

Chan, A. W., Hr’objartsson, A., Haahr, M. T., Gøtzsche, P. C., & Altman, D. G. (2004). Empirical evidence for selective reporting of outcomes in randomized trials: Comparison of protocols to published articles. In Journal of the American Medical Association. https://doi.org/10.1001/jama.291.20.2457

Chan, A. W., Hr’objartsson, A., Jørgensen, K. J., Gøtzsche, P. C., & Altman, D. G. (2008). Discrepancies in sample size calculations and data analyses reported in randomised trials: Comparison of publications with protocols. BMJ. https://doi.org/10.1136/bmj.a2299

Chan, A. W., Krleža-Jeri’c, K., Schmid, I., & Altman, D. G. (2004). Outcome reporting bias in randomized trials funded by the Canadian Institutes of Health Research. CMAJ. https://doi.org/10.1503/cmaj.1041086

Chan, A. W., Pello, A., Kitchen, J., Axentiev, A., Virtanen, J. I., Liu, A., & Hemminki, E. (2017). Association of Trial Registration With Reporting of Primary Outcomes in Protocols and Publications. JAMA, 318 (17), 1709. https://doi.org/10.1001/jama.2017.13001

Chen, T., Li, C., Qin, R., Wang, Y., Yu, D., Dodd, J., Wang, D., & Cornelius, V. (2019). Comparison of Clinical Trial Changes in Primary Outcome and Reported Intervention Effect Size Between Trial Registration and Publication. JAMA NETWORK OPEN, 2 (7). https://doi.org/10.1001/jamanetworkopen.2019.7242

Claesen, A., Gomes, S. L. B. T., tuerlinckx, francis, & vanpaemel, wolf. (2019). Preregistration: Comparing Dream to Reality [Preprint]. PsyArXiv. https://doi.org/10.31234/osf.io/d8wex

De Angelis, C., Drazen, J. M., Frizelle, F. A., Haug, C., Hoey, J., Horton, R., Kotzin, S., Laine, C., Marusic, A., Overbeke, A. J. P. M., Schroeder, T. V., Sox, H. C., & Weyden, M. B. V. D. (2004). Clinical Trial Registration: A Statement from the International Committee of Medical Journal Editors. New England Journal of Medicine, 351 (12), 1250–1251. https://doi.org/10.1056/NEJMe048225

Dekkers, O. M., Cevallos, M., Bührer, J., Poncet, A., Ackermann Rau, S., Perneger, T. V., & Egger, M. (2015). Comparison of noninferiority margins reported in protocols and publications showed incomplete and inconsistent reporting. Journal of Clinical Epidemiology. https://doi.org/10.1016/j.jclinepi.2014.09.015

Delgado, A. F., & Delgado, A. F. (2017a). Outcome switching in randomized controlled oncology trials reporting on surrogate endpoints: A cross-sectional analysis. SCIENTIFIC REPORTS, 7. https://doi.org/10.1038/s41598-017-09553-y

Delgado, A. F., & Delgado, A. F. (2017b). Inconsistent Reporting Between Meta-analysis Protocol and Publication - A Cross-Sectional Study. ANTICANCER RESEARCH, 37 (9), 5101–5107. https://doi.org/10.21873/anticanres.11928

DeVito, N. J. (2022). Trial registries for transparency and accountability in clinical research [{{Http://purl.org/dc/dcmitype/ University of Oxford.

DeVito, N. J., Bacon, S., & Goldacre, B. (2019). FDAAA TrialsTracker: A live informatics tool to monitor compliance with FDA requirements to report clinical trial results. bioRxiv, 266452. https://doi.org/10.1101/266452

Dwan, K., Gamble, C., Williamson, P. R., & Kirkham, J. J. (2013). Systematic Review of the Empirical Evidence of Study Publication Bias and Outcome Reporting Bias — An Updated Review. PLOS ONE, 8 (7), e66844. https://doi.org/10.1371/journal.pone.0066844

Dwan, K., Kirkham, J. J., Williamson, P. R., & Gamble, C. (2013). Selective reporting of outcomes in randomised controlled trials in systematic reviews of cystic fibrosis. BMJ OPEN, 3 (6). https://doi.org/10.1136/bmjopen-2013-002709

Ewart, R., Lausen, H., & Millian, N. (2009). Undisclosed changes in outcomes in randomized controlled trials: An observational study. Annals of Family Medicine. https://doi.org/10.1370/afm.1017

Fleming, P. S., Koletsi, D., Dwan, K., & Pandis, N. (2015). Outcome Discrepancies and Selective Reporting: Impacting the Leading Journals? PLOS ONE, 10 (5). https://doi.org/10.1371/journal.pone.0127495

Gandhi, R., Jan, M., Smith, H. N., Mahomed, N. N., & Bhandari, M. (2011). Comparison of published orthopaedic trauma trials following registration in Clinicaltrials.gov. BMC MUSCULOSKELETAL DISORDERS, 12. https://doi.org/10.1186/1471-2474-12-278

Goldacre, B., Drysdale, H., Dale, A., Milosevic, I., Slade, E., Hartley, P., Marston, C., Powell-Smith, A., Heneghan, C., & Mahtani, K. R. (2019). COMPare: A prospective cohort study correcting and monitoring 58 misreported trials in real time. Trials, 20 (1), 1–16. https://doi.org/10.1186/s13063-019-3173-2

Gopal, A. D., Desai, N. R., Tse, T., & Ross, J. S. (2015). Reporting of Noninferiority Trials in ClinicalTrials.gov and Corresponding Publications. JAMA-JOURNAL OF THE AMERICAN MEDICAL ASSOCIATION, 313 (11), 1163–1165. https://doi.org/10.1001/jama.2015.1697

Hahn, S., Williamson, P. R., & Hutton, J. L. (2002). Investigation of within-study selective reporting in clinical research: Follow-up of applications submitted to a local research ethics committee. Journal of Evaluation in Clinical Practice. https://doi.org/10.1046/j.1365-2753.2002.00314.x

Hannink, G., Gooszen, H. G., & Rovers, M. M. (2013). Comparison of Registered and Published Primary Outcomes in Randomized Clinical Trials of Surgical Interventions. ANNALS OF SURGERY, 257 (5), 818–823. https://doi.org/10.1097/SLA.0b013e3182864fa3

Hardwicke, T. E., Serghiou, S., Janiaud, P., Danchev, V., Crüwell, S., Goodman, S. N., & Ioannidis, J. P. A. (2020). Calibrating the Scientific Ecosystem Through Meta-Research. Annual Review of Statistics and Its Application, 7 (1), 11–37. https://doi.org/10.1146/annurev-statistics-031219-041104

Hardwicke, T. E., Thibault, R. T., Kosie, J. E., Wallach, J. D., Kidwell, M. C., & Ioannidis, J. P. A. (2021). Estimating the Prevalence of Transparency and Reproducibility-Related Research Practices in Psychology (2014–2017). Perspectives on Psychological Science, 1745691620979806. https://doi.org/10.1177/1745691620979806

Hardwicke, T. E., Wallach, J. D., Kidwell, M. C., Bendixen, T., Crüwell, S., & Ioannidis, J. P. A. (2020). An empirical assessment of transparency and reproducibility-related research practices in the social sciences (2014–2017). Royal Society Open Science, 7 (2), 190806. https://doi.org/10.1098/rsos.190806

Hartung, D. M., Zarin, D. A., Guise, J.-M., McDonagh, M., Paynter, R., & Helfand, M. (2014). Reporting Discrepancies Between the ClinicalTrials.gov Results Database and Peer-Reviewed Publications. AN-NALS OF INTERNAL MEDICINE, 160 (7), 477+. https://doi.org/10.7326/M13-0480

Hartung, J., & Knapp, G. (2003). An Alternative Test Procedure for Meta-Analysis. In Meta-analysis: New Developments and Applications in Medical and Social Sciences (pp. 53–69). Hogrefe & Huber Publishers.

Hern’andez, A. V., Steyerberg, E. W., Taylor, G. S., Marmarou, A., Habbema, J. D. F., & Maas, A. I. R. (2005). Subgroup analysis and covariate adjustment in randomized clinical trials of traumatic brain injury: A systematic review. In Neurosurgery. https://doi.org/10.1227/01.NEU.0000186039.57548.96

Higgins, J. P. T., Thompson, S. G., & Spiegelhalter, D. J. (2009). A re-evaluation of random-effects meta-analysis. *Journal of the Royal Statistical Society. Series A*, (Statistics in Society), 172 (1), 137–159. https://doi.org/10.1111/j.1467-985X.2008.00552.x

Hildenbrand, A., Conour, C., Straus, J. A., Moufarrej, S., & Palermo, T. M. (2019). Trial Registration and Outcome Reporting in Child and Pediatric Psychology: A Systematic Review. JOURNAL OF PEDIATRIC PSYCHOLOGY, 44 (9), 1024–1033. https://doi.org/10.1093/jpepsy/jsz054

Hoekstra, R., Morey, R. D., Rouder, J. N., & Wagenmakers, E. J. (2014). Robust misinterpretation of confidence intervals. Psychonomic Bulletin and Review, 21 (5), 1157–1164. https://doi.org/10.3758/ s13423-013-0572-3

Howard, B., Scott, J. T., Blubaugh, M., Roepke, B., Scheckel, C., & Vassar, M. (2017). Systematic review: Outcome reporting bias is a problem in high impact factor neurology journals. PLoS ONE, 12 (7), 1–14. https://doi.org/10.1371/journal.pone.0180986

Hui’c, M., Maruši’c, M., & Maruši’c, A. (2011). Completeness and changes in registered data and reporting bias of randomized controlled trials in ICMJE journals after trial registration policy. PLoS ONE. https://doi.org/10.1371/journal.pone.0025258

ICMJE. (2019). Recommendations for the Conduct, Reporting, Editing, and Publication of Scholarly Work in Medical Journals. ICMJE.

ICMJE. (2021). Journals stating that they follow the ICMJE Recommendations. http://www.icmje.org/journals-following-the-icmje-recommendations/.

IntHout, J., Ioannidis, J. P. A., Rovers, M. M., & Goeman, J. J. (2016). Plea for routinely presenting prediction intervals in meta-analysis. BMJ Open, 6 (7), e010247. https://doi.org/10.1136/bmjopen-2015-010247

Jones, C. W., Adams, A., Misemer, B. S., Weaver, M. A., Schroter, S., Khan, H., Margolis, B., Schriger, D. L., & Platts-Mills, T. F. (2022). Peer Reviewed Evaluation of Registered End-Points of Randomised Trials (the PRE-REPORT study): A stepped wedge, cluster-randomised trial. BMJ Open, 12 (9), e066624. https://doi.org/10.1136/bmjopen-2022-066624

Jones, C. W., Keil, L. G., Holland, W. C., Caughey, M. C., & Platts-Mills, T. F. (2015). Comparison of registered and published outcomes in randomized controlled trials: A systematic review. BMC MEDICINE, 13. https://doi.org/10.1186/s12916-015-0520-3

Jones, C. W., Misemer, B. S., Platts-Mills, T. F., Ahn, R., Woodbridge, A., Abraham, A., Saba, S., Korenstein, D., Madden, E., & Keyhani, S. (2018). Primary outcome switching among drug trials with and without principal investigator financial ties to industry: A cross-sectional study. BMJ Open, 8 (2), 1–7. https://doi.org/10.1136/bmjopen-2017-019831

Jones, C. W., & Platts-Mills, T. F. (2012). Quality of Registration for Clinical Trials Published in Emergency Medicine Journals. ANNALS OF EMERGENCY MEDICINE, 60 (4), 458–464. https://doi.org/10.1016/j.annemergmed.2012.02.005

Jones, P. M., Chow, J. T. Y., Arango, M. F., Fridfinnson, J. A., Gai, N., Lam, K., & Turkstra, T. P. (2017). Comparison of Registered anti Reported Outcomes in Randomized Clinical Trials Published in Anesthesiology Journals. ANESTHESIA AND ANALGESIA, 125 (4), 1292–1300. https://doi.org/10.1213/ANE.0000000000002272

Juri’c, D., Boli’c, A., Prani’c, S., & Maruši’c, A. (2020). Drug–drug interaction trials incompletely described drug interventions in ClinicalTrials.gov and published articles: An observational study. Journal of Clinical Epidemiology, 117, 126–137. https://doi.org/10.1016/j.jclinepi.2019.10.002

Kaplan, R. M., & Irvin, V. L. (2015). Likelihood of Null Effects of Large NHLBI Clinical Trials Has Increased over Time. PLOS ONE, 10 (8), e0132382. https://doi.org/10.1371/journal.pone.0132382

Kasenda, B., Schandelmaier, S., Sun, X., Von Elm, E., You, J., Blümle, A., Tomonaga, Y., Saccilotto, R., Amstutz, A., Bengough, T., Meerpohl, J. J., Stegert, M., Olu, K. K., Tikkinen, K. A. O., Neumann, I., Carrasco-Labra, A., Faulhaber, M., Mulla, S. M., Mertz, D., . . . Briel, M. (2014). Subgroup analyses in randomised controlled trials: Cohort study on trial protocols and journal publications. BMJ (Online). https://doi.org/10.1136/bmj.g4539

Khaleel, S., Cleveland, B., Kalapara, A., Sathianathen, N., Balaji, P., & Dahm, P. (2020). The fate of urological systematic reviews registered in PROSPERO. WORLD JOURNAL OF UROLOGY. https://doi.org/10.1007/s00345-019-03032-x

Khan, N. A., Lombeida, J. I., Singh, M., Spencer, H. J., & Torralba, K. D. (2012). Association of industry funding with the outcome and quality of randomized controlled trials of drug therapy for rheumatoid arthritis. Arthritis and Rheumatism. https://doi.org/10.1002/art.34393

Killeen, S., Sourallous, P., Hunter, I. A., Hartley, J. E., & Grady, H. L. O. (2014). Registration Rates, Adequacy of Registration, and a Comparison of Registered and Published Primary Outcomes in Randomized Controlled Trials Published in Surgery Journals. ANNALS OF SURGERY, 259 (1), 193–196. https://doi.org/10.1097/SLA.0b013e318299d00b

Korevaar, D. A., Hooft, L., Askie, L. M., Barbour, V., Faure, H., Gatsonis, C. A., Hunter, K. E., Kressel, H. Y., Lippman, H., McInnes, M. D. F., Moher, D., Rifai, N., Cohen, J. F., & Bossuyt, P. M. M. (2017). Facilitating Prospective Registration of Diagnostic Accuracy Studies: A STARD Initiative. CLINICAL CHEMISTRY, 63 (8), 1331–1341. https://doi.org/10.1373/clinchem.2017.272765

Korevaar, D. A., Ochodo, E. A., Bossuyt, P. M. M., & Hooft, L. (2014). Publication and Reporting of Test Accuracy Studies Registered in ClinicalTrials.gov. CLINICAL CHEMISTRY, 60 (4), 651–659. https://doi.org/10.1373/clinchem.2013.218149

Koufatzidou, M., Koletsi, D., Fleming, P. S., Polychronopoulou, A., & Pandis, N. (2019). Outcome reporting discrepancies between trial entries and published final reports of orthodontic randomized controlled trials. EUROPEAN JOURNAL OF ORTHODONTICS, 41 (3), 225–230. https://doi.org/10.1093/ejo/cjy046

Krsticevic, M., Saric, D., Saric, F., Slapnicar, E., Boric, K., Dosenovic, S., Kadic, A. J., Kegalj, M. J., & Puljak, L. (2019). Selective reporting bias due to discrepancies between registered and published outcomes in osteoarthritis trials. Journal of Comparative Effectiveness Research, 8 (15), 1265–1273. https://doi.org/10.2217/cer-2019-0068

Li, G., Abbade, L. P. F., Nwosu, I., Jin, Y., Leenus, A., Maaz, M., Wang, M., Bhatt, M., Zielinski, L., Sanger, N., Bantoto, B., Luo, C., Shams, I., Shahid, H., Chang, Y., Sun, G., Mbuagbaw, L., Samaan, Z., Levine, M. A. H., . . . Thabane, L. (2018). A systematic review of comparisons between protocols or registrations and full reports in primary biomedical research. BMC Medical Research Methodology, 18 (1), 9. https://doi.org/10.1186/s12874-017-0465-7

Li, X.-Q., Yang, G.-L., Tao, K.-M., Zhang, H.-Q., Zhou, Q.-H., & Ling, C.-Q. (2013). Comparison of registered and published primary outcomes in randomized controlled trials of gastroenterology and hepatology. Scandinavian Journal of Gastroenterology, 48 (12), 1474–1483. https://doi.org/10.3109/00365521.2013.845909

Ludwig, D. S., Ebbeling, C. B., & Heymsfield, S. B. (2019). Discrepancies in the Registries of Diet vs Drug Trials. JAMA Network Open, 2 (11), e1915360. https://doi.org/10.1001/jamanetworkopen.2019.15360

Mathieu, S., Boutron, I., Moher, D., Altman, D. G., & Ravaud, P. (2009). Comparison of Registered and Published Primary Outcomes in Randomized Controlled Trials. JAMA-JOURNAL OF THE AMERICAN MEDICAL ASSOCIATION, 302 (9), 977–984. https://doi.org/10.1001/jama.2009.1242

Mathieu, S., Giraudeau, B., Soubrier, M., & Ravaud, P. (2012). Misleading abstract conclusions in randomized controlled trials in rheumatology: Comparison of the abstract conclusions and the results section. Joint Bone Spine. https://doi.org/10.1016/j.jbspin.2011.05.008

Maund, E., Tendal, B., Hr’objartsson, A., Jørgensen, K. J., Lundh, A., Schroll, J., & Gøtzsche, P. C. (2014). Benefits and harms in clinical trials of duloxetine for treatment of major depressive disorder: Comparison of clinical study reports, trial registries, and publications. BMJ (Online). https://doi.org/10.1136/bmj.g3510

Mhaskar, R., Djulbegovic, B., Magazin, A., Soares, H. P., & Kumar, A. (2012). Published methodological quality of randomized controlled trials does not reflect the actual quality assessed in protocols. Journal of Clinical Epidemiology. https://doi.org/10.1016/j.jclinepi.2011.10.016

Nankervis, H., Baibergenova, A., Williams, H. C., & Thomas, K. S. (2012). Prospective Registration and Outcome-Reporting Bias in Randomized Controlled Trials of Eczema Treatments: A Systematic Review. JOURNAL OF INVESTIGATIVE DERMATOLOGY, 132 (12), 2727–2734. https://doi.org/10.1038/jid.2012.231

Norris, S. L., Holmer, H. K., Fu, R., Ogden, L. A., Viswanathan, M. S., & Abou-Setta, A. M. (2014). Clinical trial registries are of minimal use for identifying selective outcome and analysis reporting. RESEARCH SYNTHESIS METHODS, 5 (3), 273–284. https://doi.org/10.1002/jrsm.1113

Ofosu, G., & Posner, D. N. (2019). *Pre-analysis Plans: A Stocktaking* [Preprint]. MetaArXiv. https://doi.org/10.31222/osf.io/e4pum

Pandis, N., Fleming, P. S., Worthington, H., Dwan, K., & Salanti, G. (2015). Discrepancies in Outcome Reporting Exist Between Protocols and Published Oral Health Cochrane Systematic Reviews. PLOS ONE, 10 (9). https://doi.org/10.1371/journal.pone.0137667

Pinto, R. Z., Elkins, M. R., Moseley, A. M., Sherrington, C., Herbert, R. D., Maher, C. G., Ferreira, P. H., & Ferreira, M. L. (2013). Many Randomized Trials of Physical Therapy Interventions Are Not Adequately Registered: A Survey of 200 Published Trials. PHYSICAL THERAPY, 93 (3), 299–309. https://doi.org/10.2522/ptj.20120206

Prani’c, S., & Maruši’c, A. (2016). Changes to registration elements and results in a cohort of Clinicaltrials.gov trials were not reflected in published articles. Journal of Clinical Epidemiology, 70, 26–37. https://doi.org/10.1016/j.jclinepi.2015.07.007

Raghav, K. P. S., Mahajan, S., Yao, J. C., Hobbs, B. P., Berry, D. A., Pentz, R. D., Tam, A., Hong, W. K., Ellis, L. M., Abbruzzese, J., & Overman, M. J. (2015). From Protocols to Publications: A Study in Selective Reporting of Outcomes in Randomized Trials in Oncology. JOURNAL OF CLINICAL ONCOLOGY, 33 (31), 3583+. https://doi.org/10.1200/JCO.2015.62.4148

Rankin, J., Ross, A., Baker, J., O’Brien, M., Scheckel, C., & Vassar, M. (2017). Selective outcome reporting in obesity clinical trials: A cross-sectional review. CLINICAL OBESITY, 7 (4), 245–254. https://doi.org/10.1111/cob.12199

Redmond, S., Von Elm, E., Blümle, A., Gengler, M., Gsponer, T., & Egger, M. (2013). Cohort study of trials submitted to ethics committee identified discrepant reporting of outcomes in publications. Journal of Clinical Epidemiology. https://doi.org/10.1016/j.jclinepi.2013.06.020

Riehm, K. E., Azar, M., & Thombs, B. D. (2015). Transparency of outcome reporting and trial registration of randomized controlled trials in top psychosomatic and behavioral health journals: A 5-year follow-up. JOURNAL OF PSYCHOSOMATIC RESEARCH, 79 (1), 1–12. https://doi.org/10.1016/j.jpsychores.2015.04.010

Riley, R. D., Higgins, J. P. T., & Deeks, J. J. (2011). Interpretation of random effects meta-analyses. BMJ, 342, d549. https://doi.org/10.1136/bmj.d549

Rising, K., Bacchetti, P., & Bero, L. (2008). Reporting bias in drug trials submitted to the Food and Drug Administration: Review of publication and presentation. In PLoS Medicine. https://doi.org/10.1371/journal.pmed.0050217

Rongen, J. J., & Hannink, G. (2016). Comparison of Registered and Published Primary Outcomes in Randomized Controlled Trials of Orthopaedic Surgical Interventions. JOURNAL OF BONE AND JOINT SURGERY-AMERICAN VOLUME, 98 (5), 403–409. https://doi.org/10.2106/JBJS.15.00400

Rosati, P., Porzsolt, F., Ricciotti, G., Testa, G., Inglese, R., Giustini, F., Fiscarelli, E., Zazza, M., Carlino, C., Balassone, V., Fiorito, R., & D’Amico, R. (2016). Major discrepancies between what clinical trial registries record and paediatric randomised controlled trials publish. TRIALS, 17. https://doi.org/10.1186/s13063-016-1551-6

Rosenthal, R., & Dwan, K. (2013). Comparison of Randomized Controlled Trial Registry Entries and Content of Reports in Surgery Journals. ANNALS OF SURGERY, 257 (6), 1007–1015. https://doi.org/10.1097/SLA.0b013e318283cf7f

Ross, A., George, D., Wayant, C., Hamilton, T., & Vassar, M. (2019). Registration Practices of Randomized Clinical Trials in Rhinosinusitis: A Cross-sectional Review. JAMA OTOLARYNGOLOGY-HEAD & NECK SURGERY, 145 (5), 468–474. https://doi.org/10.1001/jamaoto.2019.0145

Ruegger, C. M., Dawson, J. A., Donath, S. M., Owen, L. S., & Davis, P. G. (2017). Nonpublication and discontinuation of randomised controlled trials in newborns. ACTA PAEDIATRICA, 106 (12), 1940– 1944. https://doi.org/10.1111/apa.14062

Saquib, N., Saquib, J., & Ioannidis, J. P. A. (2013). Practices and impact of primary outcome adjustment in randomized controlled trials: Meta-epidemiologic study. BMJ (Online). https://doi.org/10.1136/bmj.f4313

Scoggins, B., & Robertson, M. P. (2023). Measuring Transparency in the Social Sciences: Political Science and International Relations. ECONSTOR, 14.

Scott, A., Rucklidge, J. J., & Mulder, R. T. (2015). Is mandatory prospective trial registration working to prevent publication of unregistered trials and selective outcome reporting? An observational study of five psychiatry journals that mandate prospective clinical trial registration. PLoS ONE, 10 (8), 1–13. https://doi.org/10.1371/journal.pone.0133718

Sendyk, D. I., Rovai, E. S., Souza, N. V., Deboni, M. C. Z., & Pannuti, C. M. (2019). Selective outcome reporting in randomized clinical trials of dental implants. Journal of Clinical Periodontology, 46 (7), 758–765. https://doi.org/10.1111/jcpe.13128

Shepshelovich, D., Goldvaser, H., Wang, L., Razak, A. R. A., & Bedard, P. L. (2017). Comparison of reporting phase I trial results in ClinicalTrials.gov and matched publications. INVESTIGATIONAL NEW DRUGS, 35 (6), 827–833. https://doi.org/10.1007/s10637-017-0510-8

Shepshelovich, D., Yelin, D., Gafter-Gvili, A., Goldman, S., Avni, T., & Yahav, D. (2018). Comparison of reporting phase III randomized controlled trials of antibiotic treatment for common bacterial infections in ClinicalTrials.gov and matched publications. CLINICAL MICROBIOLOGY AND INFECTION, 24 (11). https://doi.org/10.1016/j.cmi.2018.02.010

Shinohara, K., Tajika, A., Imai, H., Takeshima, N., Hayasaka, Y., & Furukawa, T. A. (2015). Protocol registration and selective outcome reporting in recent psychiatry trials: New antidepressants and cognitive behavioural therapies. ACTA PSYCHIATRICA SCANDINAVICA, 132 (6), 489–498. https://doi.org/10.1111/acps.12502

Sim, I., Chan, A.-W., Gülmezoglu, A. M., Evans, T., & Pang, T. (2006). Clinical trial registration: Transparency is the watchword. The Lancet, 367 (9523), 1631–1633. https://doi.org/10.1016/S0140-6736(06)68708-4

Smith, H. N., Bhandari, M., Mahomed, N. N., Jan, M., & Gandhi, R. (2012). Comparison of Arthroplasty Trial Publications After Registration in ClinicalTrials.gov. JOURNAL OF ARTHROPLASTY, 27 (7), 1283–1288. https://doi.org/10.1016/j.arth.2011.11.005

Smith, S. M., Wang, A. T., Pereira, A., Chang, R. D., McKeown, A., Greene, K., Rowbotham, M. C., Burke, L. B., Coplan, P., Gilron, I., Hertz, S. H., Katz, N. P., Lin, A. H., McDermott, M. P., Papadopoulos, E. J., Rappaport, B. A., Sweeney, M., Turk, D. C., & Dworkin, R. H. (2013). Discrepancies between registered and published primary outcome specifications in analgesic trials: ACTTION systematic review and recommendations. PAIN, 154 (12), 2769–2774. https://doi.org/10.1016/j.pain.2013.08.011

Soares, H. P., Daniels, S., Kumar, A., Clarke, M., Scott, C., Swann, S., & Djulbegovic, B. (2004). Bad reporting does not mean bad methods for randomised trials: Observational study of randomised controlled trials performed by the Radiation Therapy Oncology Group. British Medical Journal. https://doi.org/10.1136/bmj.328.7430.22

Stockmann, C., Ross, J. S., Sherwin, C. M. T., Reilly, C. A., McDowell, B., Fassl, B., Nkoy, F., Maloney, C. G., & Spigarelli, M. G. (2014). Rate of asthma trial outcomes reporting on ClinicalTrials.gov and in the published literature. JOURNAL OF ALLERGY AND CLINICAL IMMUNOLOGY, 134 (6), 1443–1446. https://doi.org/10.1016/j.jaci.2014.09.019

Su, C.-X., Han, M., Ren, J., Li, W.-Y., Yue, S.-J., Hao, Y.-F., & Liu, J.-P. (2015). Empirical evidence for outcome reporting bias in randomized clinical trials of acupuncture: Comparison of registered records and subsequent publications. TRIALS, 16. https://doi.org/10.1186/s13063-014-0545-5

TARG Meta-Research Group & Collaborators. (2022). Discrepancy review: A feasibility study of a novel peer review intervention to reduce undisclosed discrepancies between registrations and publications | Royal Society Open Science. Royal Society Open Science. https://doi.org/10.1098/rsos.220142

Thibault, R. T., Pennington, C. R., & Munaf‘o, M. R. (2023). Reflections on Preregistration: Core Criteria, Badges, Complementary Workflows. Journal of Trial & Error. https://doi.org/10.36850/mr6

Thomas, E. T., Clark, J., & Glasziou, P. (2016). Publication and outcome reporting of homeopathy trials registered in clinicaltrials.gov. Focus on Alternative and Complementary Therapies, 21 (3-4), 127–133. https://doi.org/10.1111/fct.12278

Tricco, A. C., Cogo, E., Page, M. J., Polisena, J., Booth, A., Dwan, K., MacDonald, H., Clifford, T. J., Stewart, L. A., Straus, S. E., & Moher, D. (2016). A third of systematic reviews changed or did not specify the primary outcome: A PROSPERO register study. JOURNAL OF CLINICAL EPIDEMIOLOGY, 79, 46–54. https://doi.org/10.1016/j.jclinepi.2016.03.025

Vedula, S. Swaroop Bero, L., Scherer, R. W., & Dickersin, K. (2009). Outcome Reporting in Industry-Sponsored Trials of Gabapentin for Off-Label Use. NEW ENGLAND JOURNAL OF MEDICINE, 361 (20), 1963–1971. https://doi.org/10.1056/NEJMsa0906126

Vedula, S. Swaroop, Li, T., & Dickersin, K. (2013). Differences in Reporting of Analyses in Internal Company Documents Versus Published Trial Reports: Comparisons in Industry-Sponsored Trials in Off-Label Uses of Gabapentin. PLoS Medicine. https://doi.org/10.1371/journal.pmed.1001378

Walker, K. F., Stevenson, G., & Thornton, J. G. (2014). Discrepancies between registration and publication of randomised controlled trials: An observational study. JRSM Short Reports, 5 (5), 1–4. https://doi.org/10.1177/2042533313517688

Wandalkar, P., Gandhe, P., Pai, A., Limaye, M., Chauthankar, S., Gogtay, N., & Thatte, U. (2017). A study comparing trial registry entries of randomized controlled trials with publications of their results in a high impact factor journal: The Journal of the American Medical Association. Perspectives in Clinical Research, 8 (4), 167–171. https://doi.org/10.4103/2229-3485.215978

Wayant, C., Scheckel, C., Hicks, C., Nissen, T., Leduc, L., Som, M., & Vassar, M. (2017). Evidence of selective reporting bias in hematology journals: A systematic review. PLOS ONE, 12 (6). https://doi.org/10.1371/journal.pone.0178379

White, V. A., Walker, K. F., & Thornton, J. G. (2019). Trials of antenatal corticosteroids for preterm fetal lung maturity: A review of the potential for selective outcome reporting. EUROPEAN JOURNAL OF OBSTETRICS & GYNECOLOGY AND REPRODUCTIVE BIOLOGY, 236, 58–68. https://doi.org/10.1016/j.ejogrb.2019.02.031

Wiebe, J., Detten, G., Scheckel, C., Gearhart, D., Wheeler, D., Sanders, D., & Vassar, M. (2017). The heart of the matter: Outcome reporting bias and registration status in cardio-thoracic surgery. INTERNATIONAL JOURNAL OF CARDIOLOGY, 227, 299–304. https://doi.org/10.1016/j.ijcard.2016.11.098

Wildt, S., Krag, A., & Gluud, L. (2011). Characteristics of randomised trials on diseases in the digestive system registered in ClinicalTrials.gov: A retrospective analysis. BMJ OPEN, 1 (2). https://doi.org/10.1136/bmjopen-2011-000309

Won, J., Kim, S., Bae, I., & Lee, H. (2019). Trial registration as a safeguard against outcome reporting bias and spin? A case study of randomized controlled trials of acupuncture. PLoS ONE, 14 (10). https://doi.org/10.1371/journal.pone.0223305

World Medical Association. (2013). Declaration of Helsinki – Ethical Principles for Medical Research Involving Human Subjects. World Medical Association.

You, B., Gan, H. K., Pond, G., & Chen, E. X. (2012). Consistency in the analysis and reporting of primary end points in oncology randomized controlled trials from registration to publication: A systematic review. In Journal of Clinical Oncology. https://doi.org/10.1200/JCO.2011.37.0890

Zarin, Deborah A., Tse, T., Williams, R. J., Califf, R. M., & Ide, N. C. (2011). The ClinicalTrials.gov Results Database - Update and Key Issues. NEW ENGLAND JOURNAL OF MEDICINE, 364 (9), 852–860. https://doi.org/10.1056/NEJMsa1012065

Zarin, Deborah A., Tse, T., Williams, R. J., & Rajakannan, T. (2017). Update on Trial Registration 11 Years after the ICMJE Policy Was Established. New England Journal of Medicine, 376 (4), 383–391. https://doi.org/10.1056/NEJMsr1601330

Zhang, S., Liang, F., & Li, W. (2017). Comparison between publicly accessible publications, registries, and protocols of phase III trials indicated persistence of selective outcome reporting. JOURNAL OF CLINICAL EPIDEMIOLOGY, 91, 87–94. https://doi.org/10.1016/j.jclinepi.2017.07.010

