## Supplementary Material for "Estimating the prevalence of discrepancies between study registrations and publications: A systematic review and meta-analyses"

### Supplementary materials

#### A. Protocol amendments

As our systematic review was largely exploratory, many elements of the protocol are detailed more precisely in the manuscript than they were in our preregistered study protocol. We made the following amendments to this protocol:

- We included additional contributors and some of the projected contributor roles changed.
- Before beginning data extraction, we operationalized Table 2 from the protocol into a Qualtrics survey form and made the items more precise and more detailed.
- The protocol uses the term "*preregistration-publication discrepancies*". However, because many articles were unclear whether the registrations were prospective, we included data on discrepancies between publications and registrations regardless of the timing of registration. We also include discrepancies between publications and protocols, ethics applications, grant applications, and marketing applications. We did not foresee articles being published on discrepancies between these documents, but believe them to be relevant to our systematic review.
- Whereas the protocol states that the review will be descriptive, we used inferential statistics to run meta-analyses and to test for parameters that might influence the prevalence of discrepancies (i.e., the analyses presented in Table 2). We had preregistered that we would search for "*Additional inferential statistics relevant to preregistration–publication discrepancies*", but did not preregister that we would run our own inferential analyses. We used inferential statistics because we found many more articles than we anticipated and we developed our data extraction form to have more precision than we originally intended.
- The scoping review was terminated, as explained in the updated version of the registration associated with this study.
- When screening, for that sake of simplicity, we did not end up using the bin "*(iv) relevant to preregistration but not directly relevant to our study objectives.*"
- A peer reviewer suggested we run a meta-regression which includes all the article-level characteristics. We ran this meta-regression after knowing the results of all the meta-regressions we ran for single article-level characteristics. We report on this analysis in Supplementary Material E.
- We originally planned to share our data and code on the University of Bristol Data Repository. Instead, we have shared it on osf.io (because more people are familiar with this platform) and on www.codeocean.com (because this platform supports reproducible containers).

#### B. Coding

The coding form is divided into five sections and available at osf.io/728ys. Section One coded the search strategy and design of each article, including how the researchers identified studies (e.g., through registries or journals), first and last year of publication of the assessed studies, whether the article was solely observational, the research discipline assessed (as defined by the Scopus discipline categories outlined in Appendix C of our preregistered protocol), and a verbatim copy of the main conclusion that was relevant to findings on discrepancies.

Section Two coded when the assessed studies were registered (e.g., prospectively, retrospectively, unclear), the version of the registration used when checking for discrepancies (e.g., original version, most recent version), and whether the article checked if authors disclosed reasons for any of the discrepancies.

Section Three coded outcome discrepancies. Whereas some articles made it clear that they only assessed studies that were prospectively registered, others were unclear about when the studies they assessed were registered. We coded discrepancies for unambiguously preregistered studies separate from studies with unclear registration timing or retrospective registration. If an article reported discrepancies separately for prospectively and retrospectively registered studies, we coded both these categories. We divided outcome discrepancies into 10 non-exclusive subcategories: any outcome discrepancy, a primary outcome discrepancy, a secondary outcome discrepancy, demoting a primary outcome to a secondary outcome, omitting a primary outcome, adding a primary outcome, promoting a secondary outcome to a primary outcome, omitting a secondary outcome, adding a secondary outcome, and changing the timing at which an outcome was measured. We only coded *any outcome discrepancy* for articles that checked for both primary and secondary outcome discrepancies in the studies they assessed. If we had also included articles that only checked for primary outcome discrepancies in this sub-category, then we would have biased our estimate downwards because the assessed studies may also have secondary outcome discrepancies, but that information would not be documented. Articles used slightly different methods for identifying and defining primary outcome discrepancies. Rather than use a stringent definition of what constitutes a primary outcome discrepancy across all articles, we coded the number of studies with primary outcome discrepancies based on the data presented in the results section of each article, regardless of the method or definition the article used. A definition used in many articles comes from Chan, Hr'objartsson, et al. (2004) and includes (1) demoting a primary outcome to a secondary outcome, (2) omitting a primary outcome, (3) adding a primary outcome, (4) promoting a secondary outcome to a primary outcome, and (5) changing the timing at which a primary outcome is measured.

Section Four coded non-outcome discrepancies. These included discrepancies in participant eligibility (e.g., inclusions and exclusion criteria), sample size, randomization (e.g., number of experimental arms), method of blinding (e.g., single versus double blind), interventions (e.g., a change in the drug administered), study duration, analyses, subgroup analyses, funding, and reported results. There was heterogeneity between articles in their method for identifying and defining each type of discrepancy. As we did for the outcome discrepancies, we coded the number of studies with each type of discrepancies based on the data presented in the results section of each article, regardless of the method or definition the article used.

Section Five noted whether articles provided any additional descriptive or inferential statistics on discrepancies. In particular, it noted whether discrepancies were related to funding source, statistical significance, or the timing of registration.

For each type of discrepancy, we coded the numbers of publications containing at least one discrepancy of that type, rather than coding the total number of discrepancies of each type per publication (e.g., a single publication may have five discrepancies in secondary outcomes). Of the k = 89 articles we reviewed, 17 reported a measure of the total number of discrepancies. Sixteen of these 17 articles also reported the number of publications with at least one discrepancy, and as such we were able to include them in our analysis. We excluded the one remaining article that did not report the number of publications with at least one discrepancy. We do not analyze the total number of discrepancies in the present report.

After completing the data extraction and analyses described above, we extracted additional information from articles that we had identified as providing information on whether discrepancies were disclosed, as well as whether discrepancies were related to funding source, statistical significance, or the timing of registration. We extracted this information directly into a spreadsheet, we did not create an additional coding form. We divided studies into industry funded or non-industry funded (which included public, private, and no funding). We excluded studies from this analysis if they did not report their funding or were funded by both industry and non-industry sources. We also coded the proportion of discrepancies that favored statistical significance. Favoring statistical significance included when a novel significant outcome was introduced, or a non-significant primary outcome was downgraded to a secondary outcome. It is impossible to know whether omitted outcomes were statistically significant or not. If we were to assume that the majority of omitted outcomes were non-significant, then this analysis would provide an underestimation of the proportion of studies that favor statistical significance.

#### C. Article characteristics

We identified and reviewed k = 89 articles that report at least one type of discrepancy. The vast majority of these articles assessed medical research (k = 81). The remaining articles focused on dentistry (k = 3), psychology (k = 3), physical therapy (k = 1), and economics (k = 1).

Articles compared publications to registrations deposited in conventional registries such as clinicaltrials.gov (k = 73), or to protocols submitted for research ethics review (k = 7), funding applications (k = 2), marketing approval (k = 2), or other purposes (k = 5). The 89 articles we reviewed identified studies through different sources, including journals (k = 33), registries (k = 27), search engines (k = 19), research ethics boards (k = 7), funders (k = 3), regulatory agencies (k = 2), and research groups (k = 2). Four articles used multiple sources. Seventy-one articles reported the year the earliest study they included was published (median = 2009, IQR: 2004-2012), and 80 reported the last year (median = 2013, IQR: 2010-2015).

All but 10 articles were solely observational. Three of these 10 articles emailed authors for registration numbers (Ross et al., 2019; Shinohara et al., 2015; Wiebe et al., 2017) and one emailed authors for full protocols (Saquib et al., 2013). Four articles sent a survey to researchers to ask about the discrepancies (Chan, Hr'objartsson, et al., 2004; Chan, Krleža-Jeri'c, et al., 2004; Redmond et al., 2013; Wayant et al., 2017). One article sent a survey to better understand user experience with pre-analysis plans (Ofosu & Posner, 2019). Only one article attempted to correct published discrepancies. They sent letters to the editor within weeks of a study being published with outcome discrepancies (Goldacre et al., 2019). We do not further discuss the non-observational aspects of these studies in this review.

Whereas k = 29 articles clearly state that they compared the publications to the original registry entry or protocol, 15 used the most recent version of the registry entry, 35 do not mention which registry version they used, and 10 used a different version of the registration or were unclear about the version they used.

#### D. Meta-analyses of each outcome and non-outcome discrepancy separated by timing of registration

Here, we performed two additional sets of random effects meta-analyses: (1) for articles that reported on unambiguously prospectively registered studies (i.e., studies that were clearly prospectively registered), (2) for articles that reported on studies where the timing of registration was unclear.

While some of the outcome discrepancies may appear to occur more frequently in studies that were unambiguously identified as prospectively registered, as compared to those with unclear timing of registration, the confidence intervals are largely overlapping. And yet, we cannot ascertain that prospectively and retrospectively registered studies have a similar frequency of discrepancies because the unclear category consists of both retrospectively and prospectively registered studies in an unknown proportion. We deemed it unreasonable to compare unambiguously prospectively registered studies to studies with unclear timing of registration because there were too few studies and their methods were highly heterogeneous.

Supplementary Table D1. Meta-analytic estimates for the proportion of studies that contain various types of outcome discrepancies and non-outcome discrepancies. Articles are separated into those which unambiguously assessed only prospectively registered studies versus articles where the registration timing of the assessed studies was retrospective, unclear, or mixed. This table is essentially a combination of Table 2 and Table 4 from the manuscript, but grouped by timing of registration.

| | Prospective registration | | | | Unclear registration timing | | | |
|---|---|---|---|---|---|---|---|---|
| | 95% CI | 95% PI | k | n | 95% CI | 95% PI | k | n |
| Any outcome discrepancy | 38-91% | 5-99% | 6 | 279 | 32-70% | 6-95% | 13 | 850 |
| Any primary outcome discrepancy | 25-38% | 11-64% | 24 | 1767 | 28-39% | 9-72% | 51 | 4857 |
| Any secondary outcome discrepancy | 46-87% | 12-98% | 7 | 506 | 44-74% | 11-95% | 16 | 1052 |
| Primary outcome demoted to secondary outcome | 7-18% | 2-44% | 15 | 1287 | 5-9% | 2-27% | 36 | 3273 |
| Primary outcome omitted | 6-16% | 1-56% | 20 | 1600 | 6-11% | 1-36% | 32 | 2860 |
| Primary outcome added | 7-14% | 3-30% | 18 | 1403 | 6-12% | 1-37% | 36 | 3294 |
| Secondary outcome promoted to primary outcome | 3-9% | 1-25% | 14 | 1250 | 3-6% | 1-18% | 32 | 2885 |
| Secondary outcome omitted | 14-62% | 2-93% | 7 | 566 | 14-31% | 5-58% | 12 | 799 |
| Secondary outcome added | 8-80% | 0.3-99% | 6 | 413 | 18-41% | 5-73% | 14 | 892 |
| Timing of outcome measurement changed | 3-18% | 1-55% | 9 | 864 | 6-19% | 1-68% | 21 | 2608 |
| Eligibility criteria | 1-99% | 23-67% | 2 | 50 | 23-59% | 4-92% | 13 | 1103 |
| Sample size | 40-59% | 39-60% | 6 | 174 | 22-43% | 6-78% | 20 | 1244 |
| Randomization | 46-70% | 46-70% | 1 | 58 | 1-56% | 0.02-98% | 4 | 118 |
| Blinding | NA | NA | NA | NA | 5-42% | 0.04-99% | 3 | 224 |
| Intervention | 1-22% | 1-22% | 1 | 27 | 3-63% | 0.1-98% | 6 | 523 |
| Study duration | NA | NA | NA | NA | 3-89% | 0.02-99.94% | 4 | 184 |
| Analysis | 24-71% | 6-92% | 6 | 232 | 7-46% | 2-80% | 6 | 172 |
| Subgroup analysis | 7-99% | 0.01-99.998% | 4 | 215 | 20-97% | 1-99.93% | 5 | 330 |
| Funding | 7-67% | 7-67% | 1 | 7 | 4-93% | 0.01-99.98% | 4 | 205 |
| Results | 4-20% | 4-20% | 1 | 54 | 7-91% | 0.1-99.8% | 5 | 208 |

Values less than 0.5% and greater than 99.5% are rounded to one significant digit from 0 or 100. Prediction intervals cannot be calculated for meta-analyses with fewer than three studies.

**The 38 forest plots that follow align with the meta-analyses presented in Supplementary Table D1.**

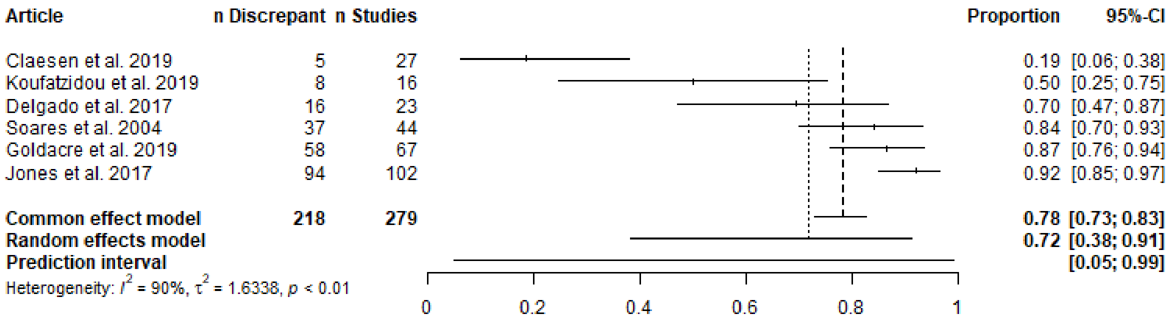


Supplementary Figure D1. Any outcome discrepancy, prospective.

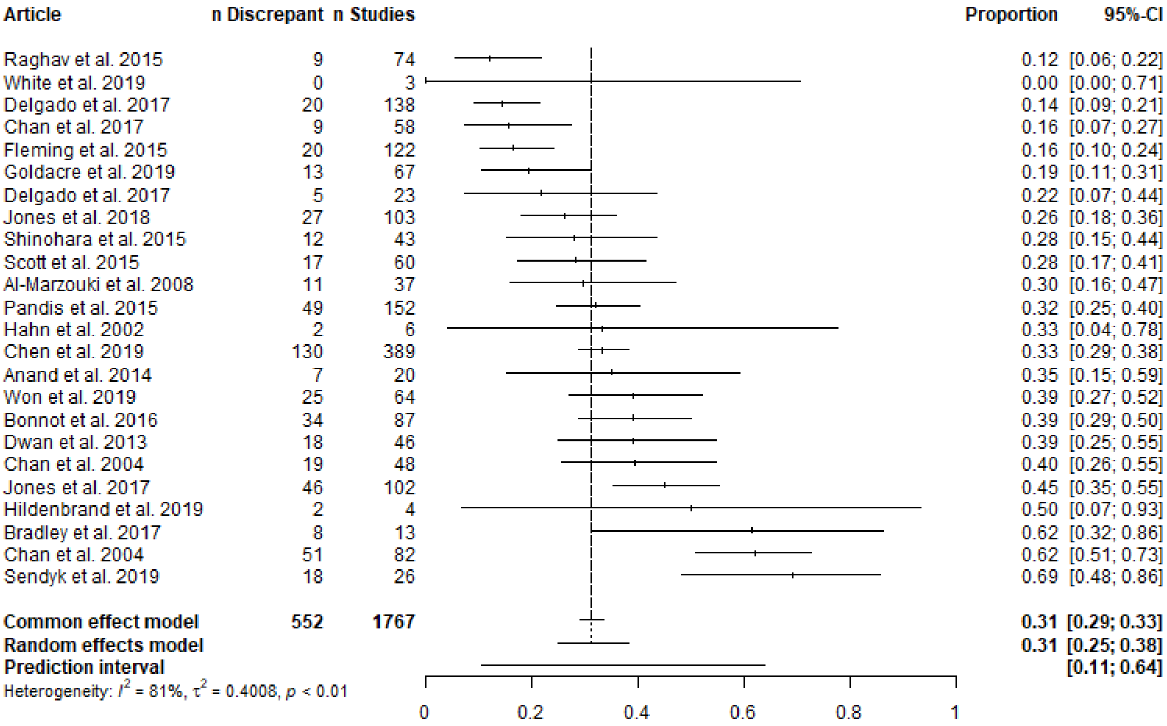


Supplementary Figure D2. Any primary outcome discrepancy, prospective.

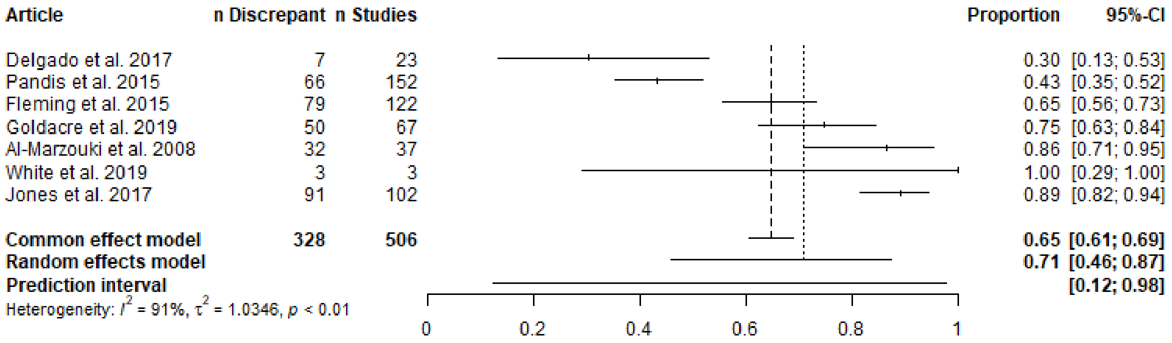


Supplementary Figure D3. Any secondary outcome discrepancy, prospective.

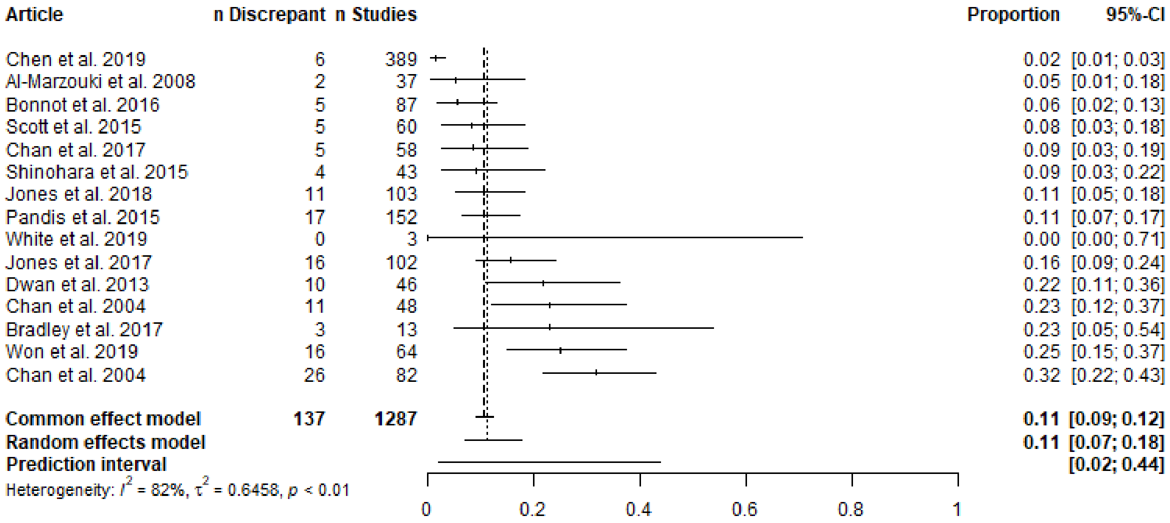


Supplementary Figure D4. Primary outcome demoted to secondary outcome, prospective.

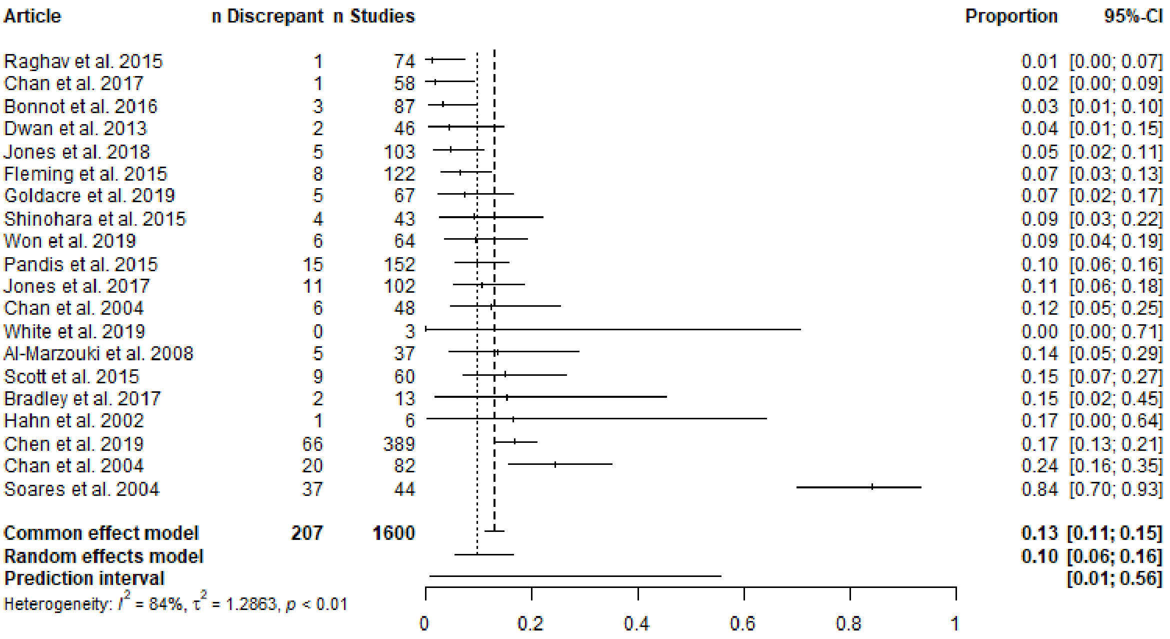


Supplementary Figure D5. Primary outcome omitted, prospective.

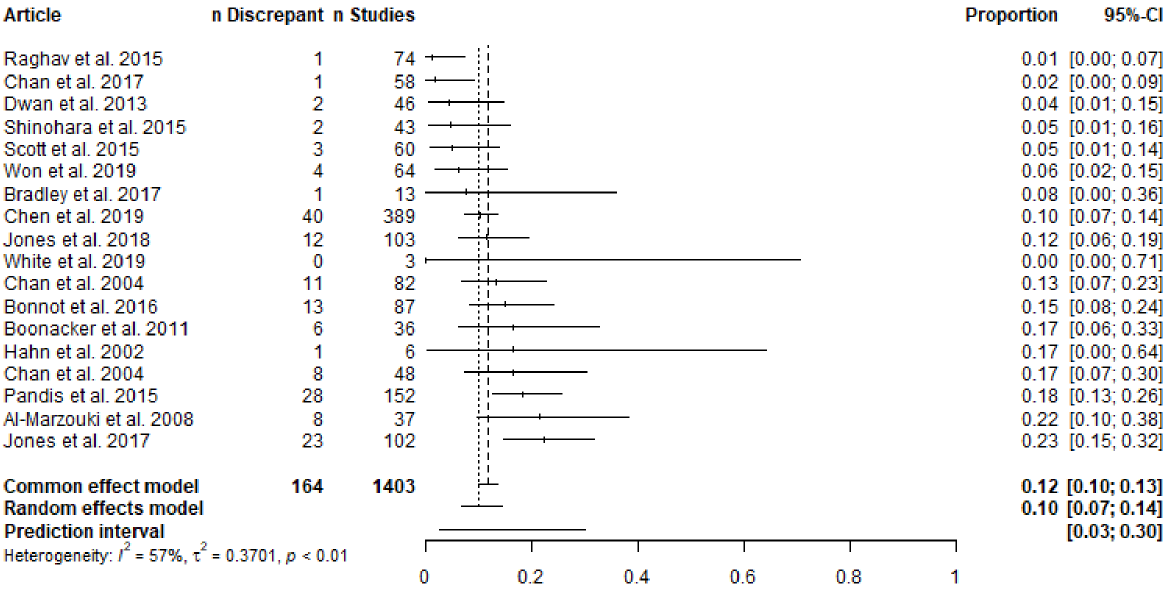


Supplementary Figure D6. Primary outcome added, prospective.

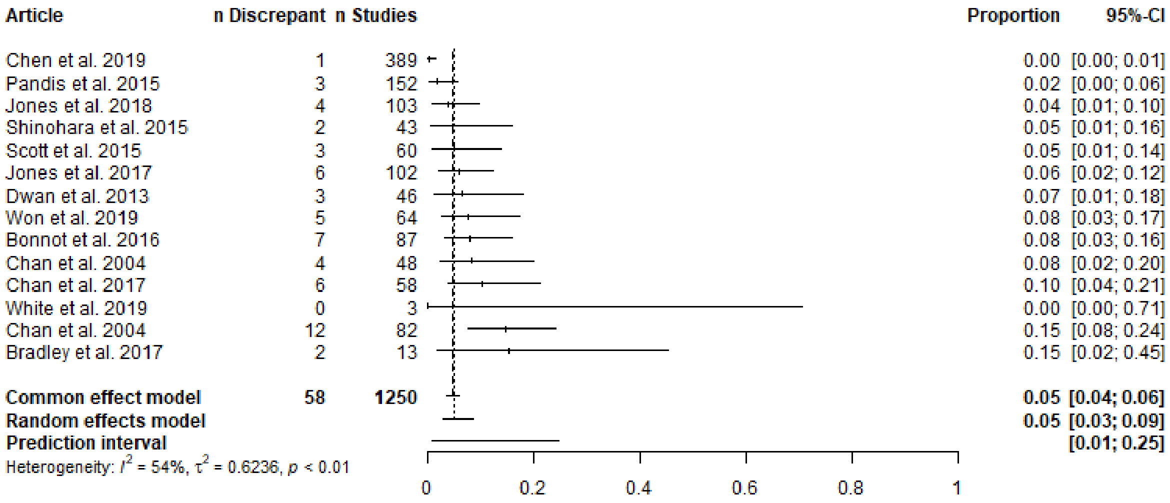


Supplementary Figure D7. Secondary outcome promoted to primary outcome, prospective.

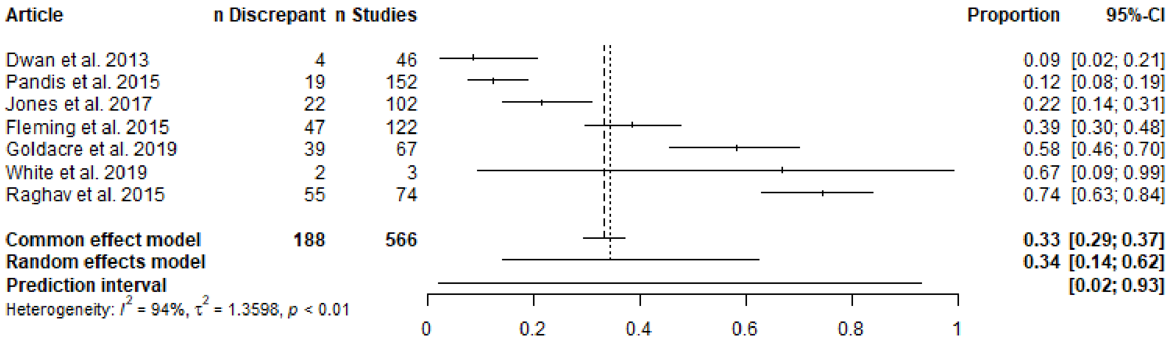


Supplementary Figure D8. Secondary outcome omitted, prospective.

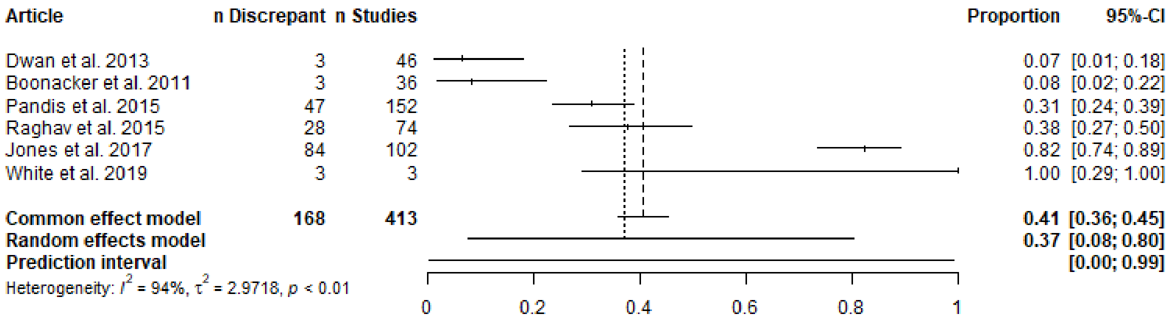


Supplementary Figure D9. Secondary outcome added, prospective.

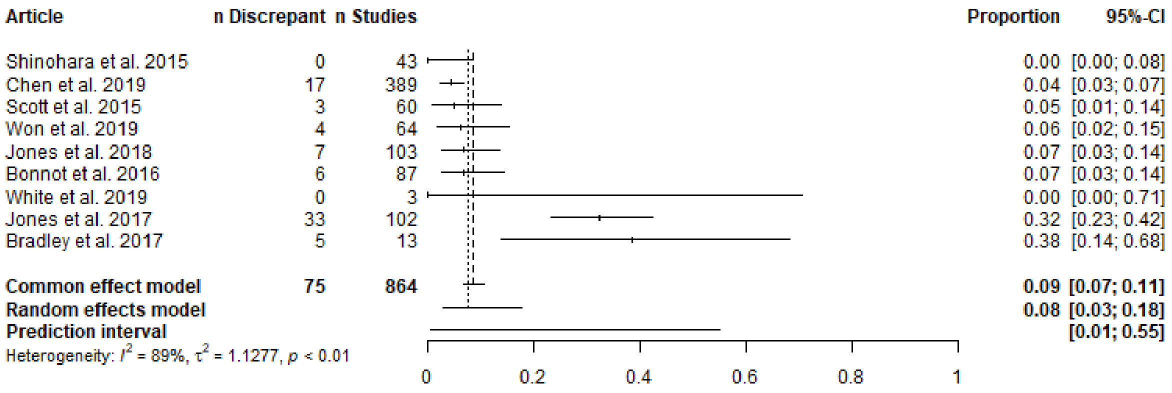


Supplementary Figure D10. Timing of outcome measurement changed, prospective.

| Article | n Discrepant | n Studies | Proportion | 95%-CI |
|---|---|---|---|---|
| Rankin et al. 2017 | 22 | 142 | 0.15 | [0.10; 0.23] |
| Braakhekke et al. 2017 | 21 | 114 | 0.18 | [0.12; 0.27] |
| Gandhi et al. 2011 | 4 | 20 | 0.20 | [0.06; 0.44] |
| Korevaar et al. 2014 | 32 | 131 | 0.24 | [0.17; 0.33] |
| Ross et al. 2019 | 22 | 70 | 0.31 | [0.21; 0.44] |
| Riehm et al. 2015 | 1 | 3 | 0.33 | [0.01; 0.91] |
| Koufatzidou et al. 2019 | 25 | 53 | 0.47 | [0.33; 0.61] |
| Shepshelovich et al. 2017 | 118 | 163 | 0.72 | [0.65; 0.79] |
| Norris et al. 2014 | 27 | 36 | 0.75 | [0.58; 0.88] |
| Aggarwal et al. 2019 | 19 | 24 | 0.79 | [0.58; 0.93] |
| Sendyk et al. 2019 | 39 | 49 | 0.80 | [0.66; 0.90] |
| Chahal et al. 2012 | 16 | 20 | 0.80 | [0.56; 0.94] |
| Smith et al. 2012 | 22 | 25 | 0.88 | [0.69; 0.97] |
| **Common effect model** | **368** | **850** | **0.43** | **[0.40; 0.47]** |
| **Random effects model** | | | **0.52** | **[0.32; 0.70]** |
| **Prediction interval** | | | | **[0.06; 0.95]** |

Heterogeneity: $I^2 = 94\%$, $\tau^2 = 1.5464$, $p < 0.01$

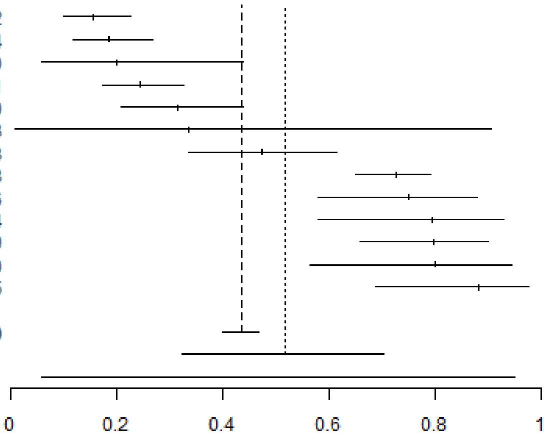


Supplementary Figure D11. Any outcome discrepancy, unclear registration timing.

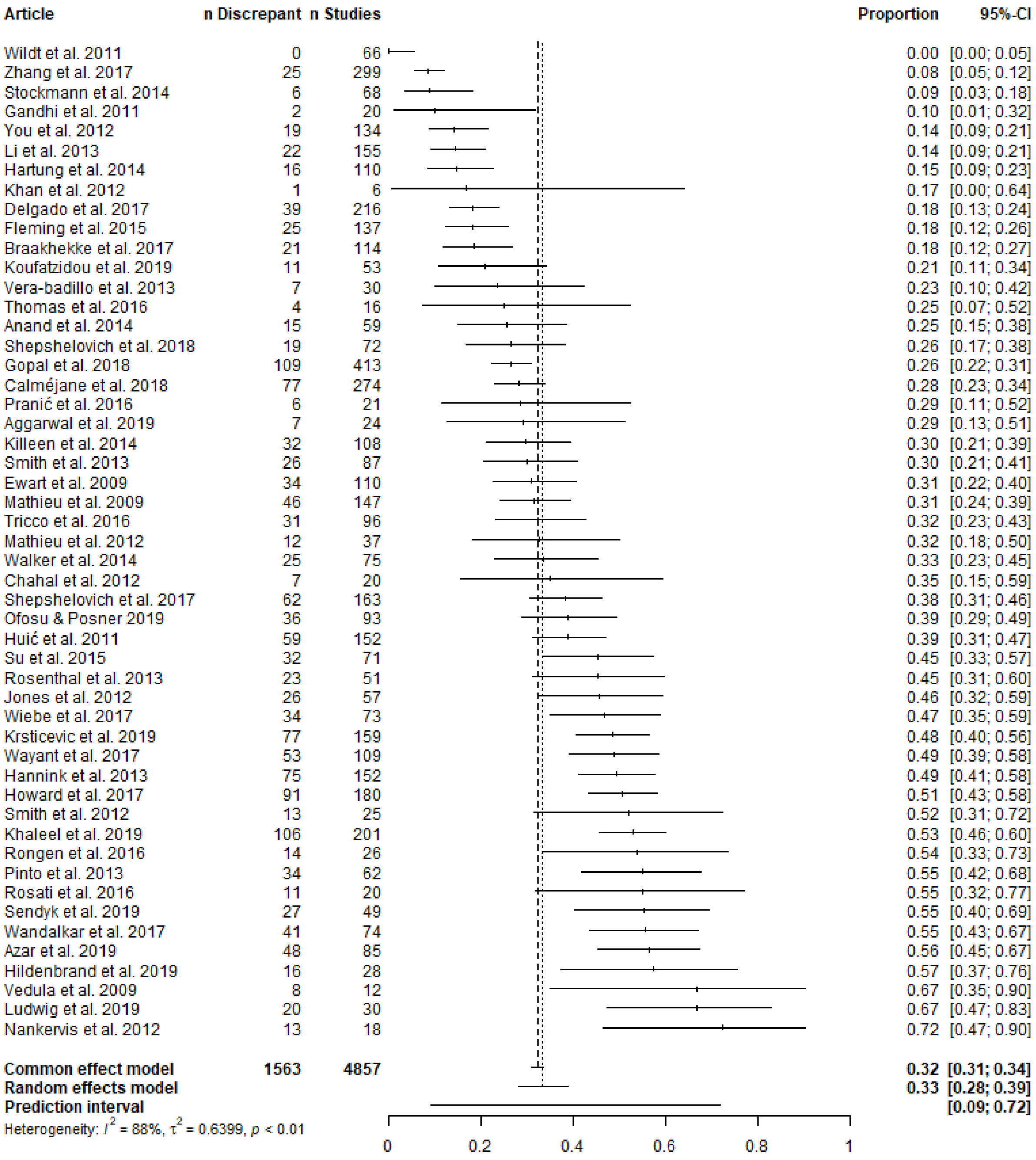


Supplementary Figure D12. Any primary outcome discrepancy, unclear registration timing.

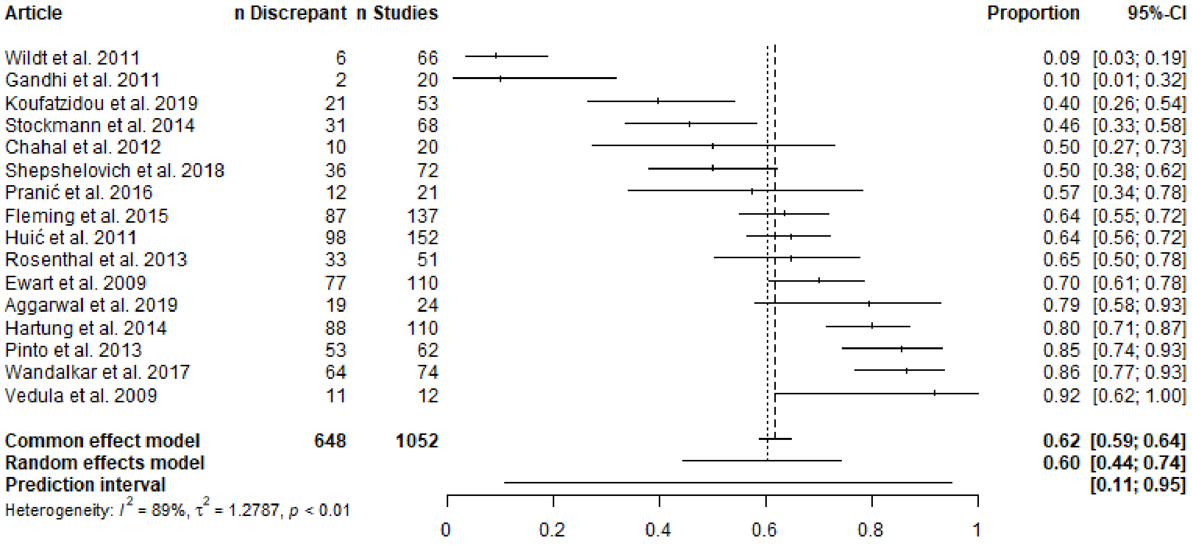


Supplementary Figure D13. Any secondary outcome discrepancy, unclear registration timing.

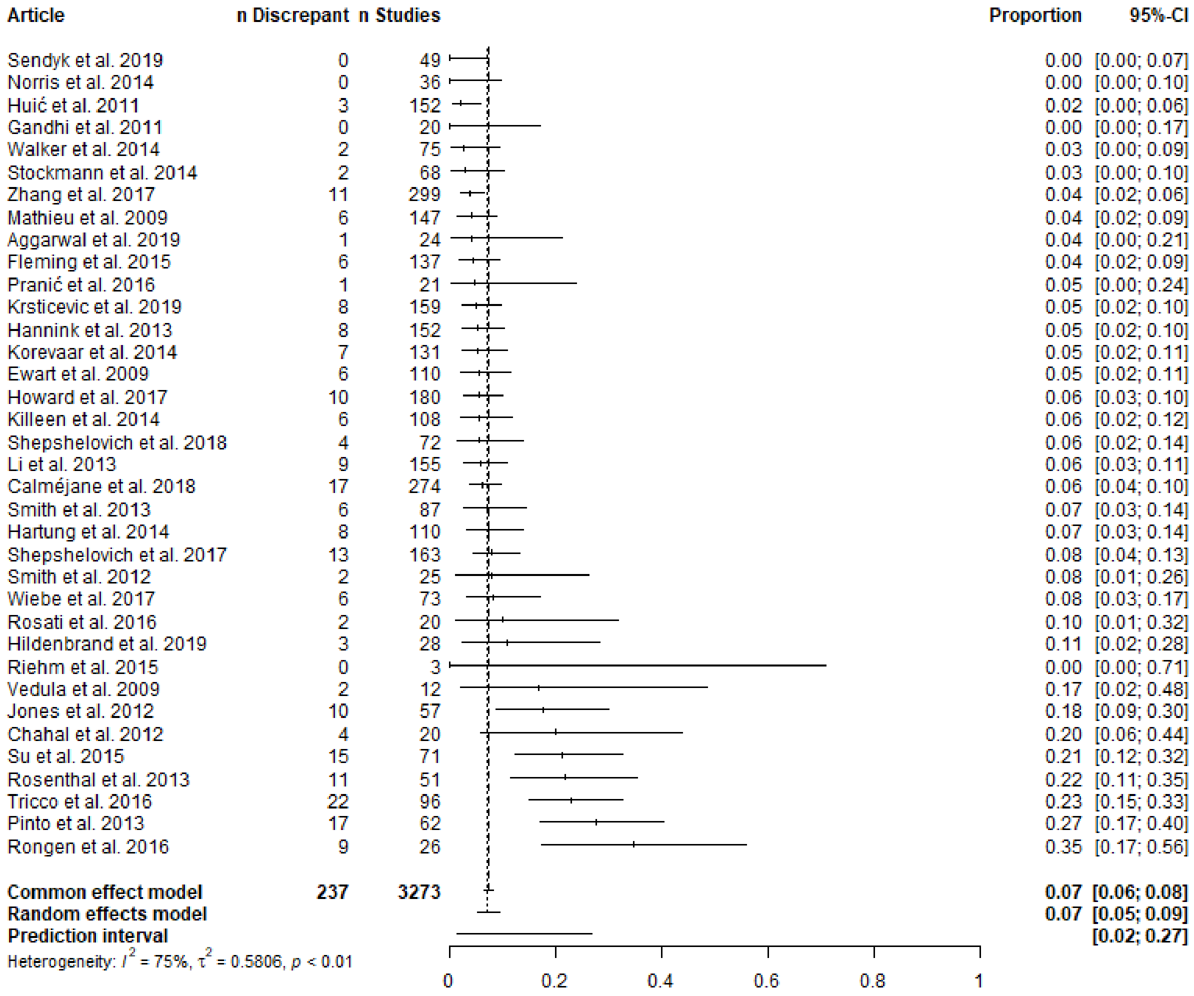


Supplementary Figure D14. Primary outcome demoted to secondary outcome, unclear registration timing.

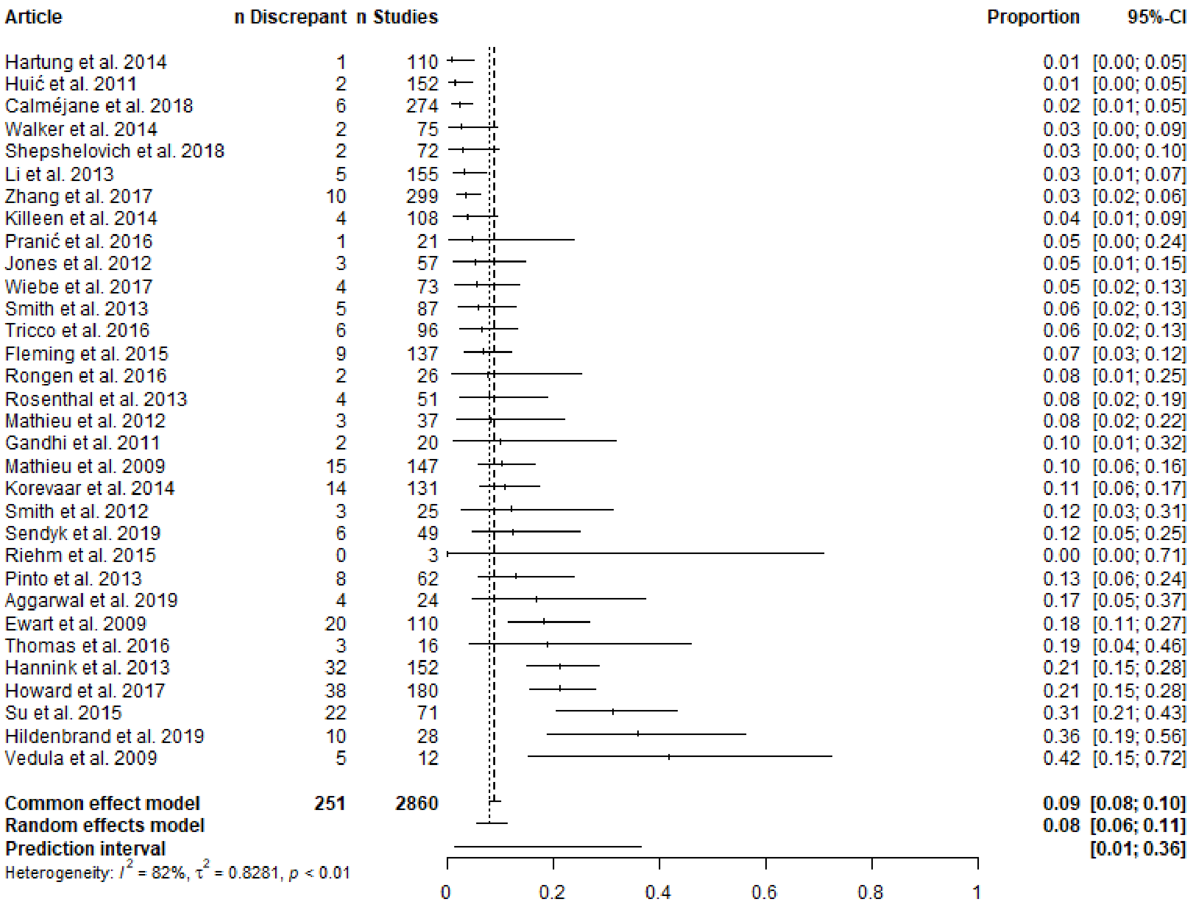


Supplementary Figure D15. Primary outcome omitted, unclear registration timing.

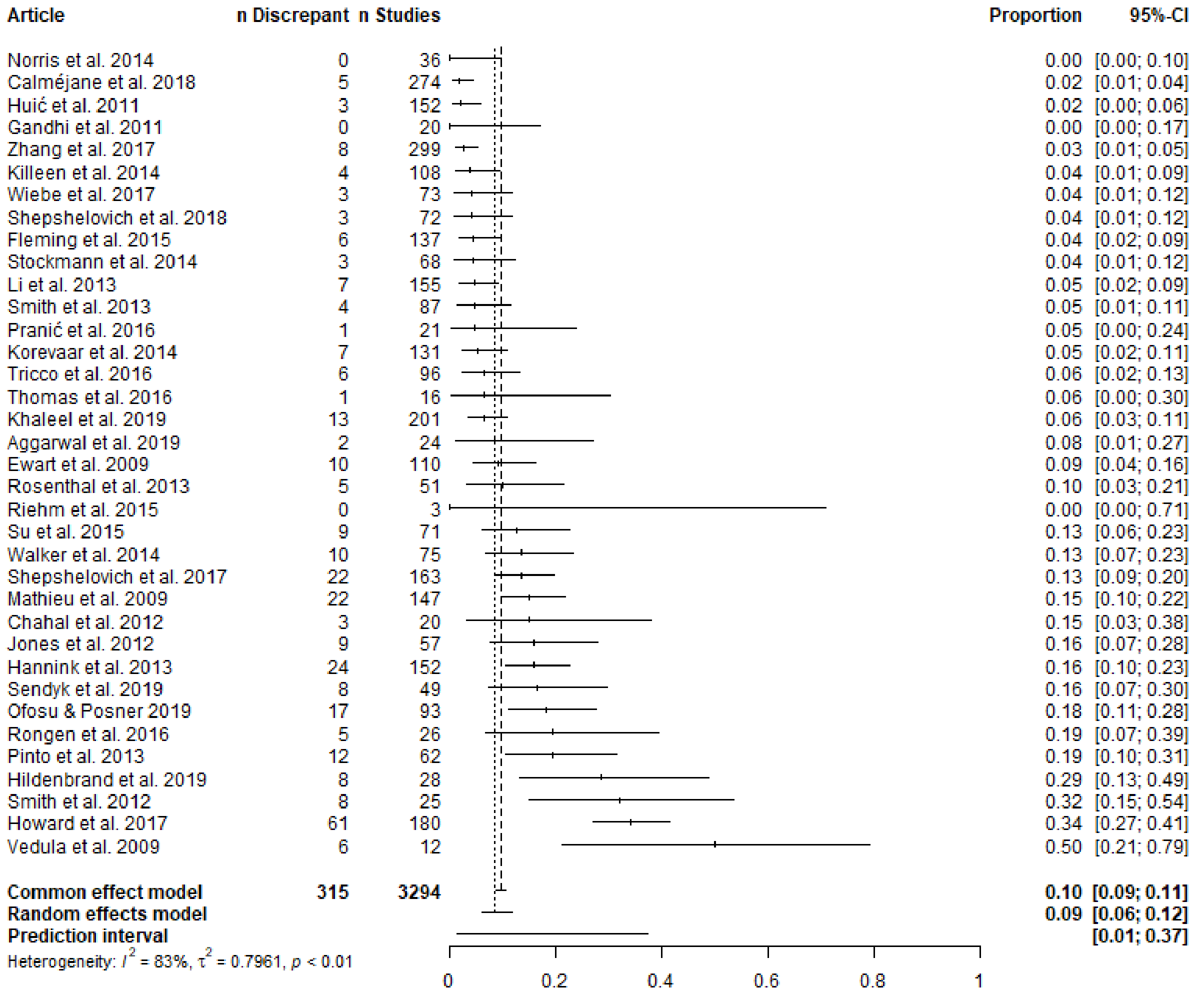


Supplementary Figure D16. Primary outcome added, unclear registration timing.

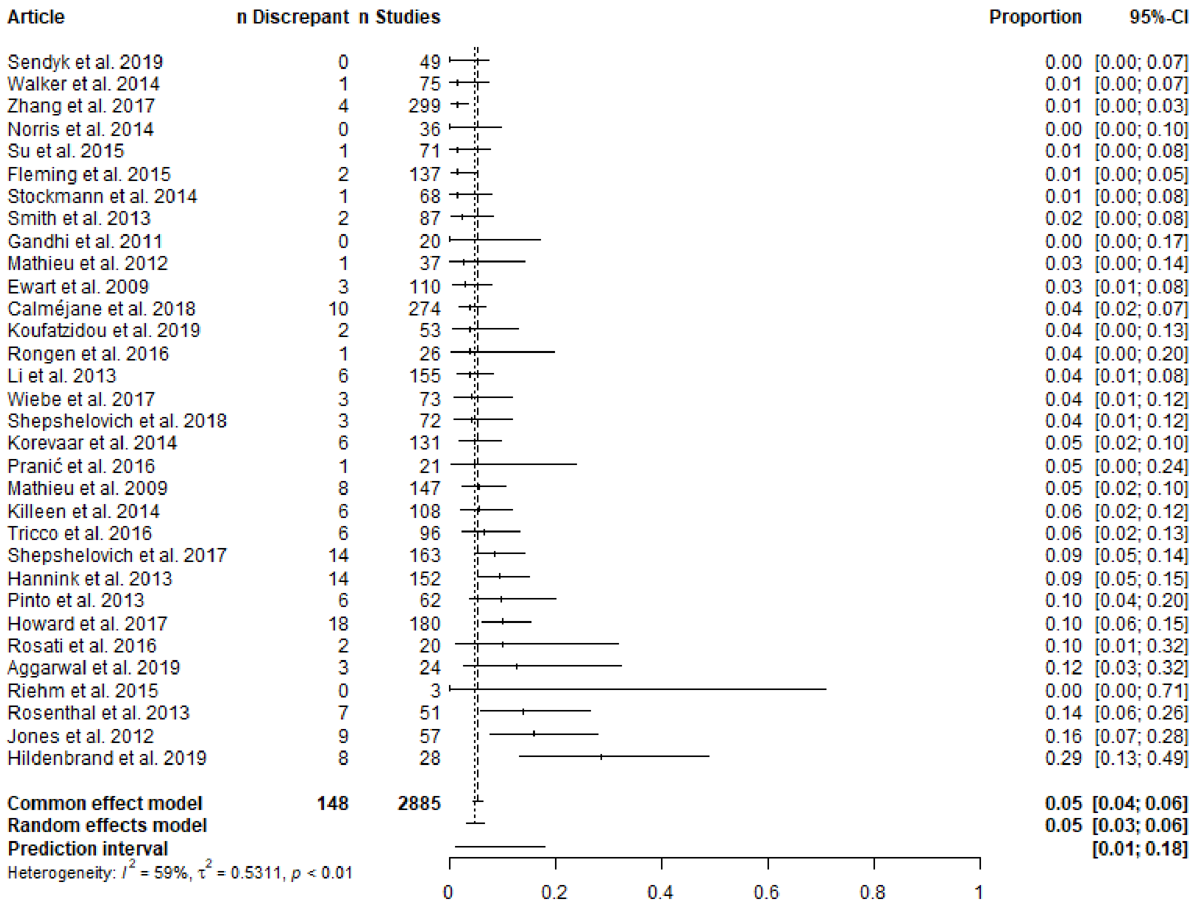


Supplementary Figure D17. Secondary outcome promoted to primary outcome, unclear registration timing.

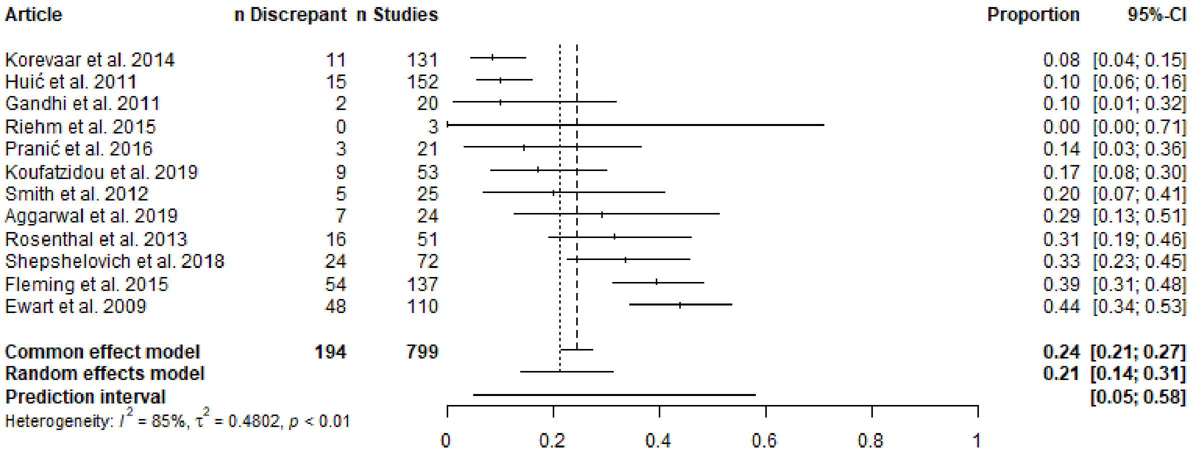


Supplementary Figure D17. Secondary outcome omitted, unclear registration timing.

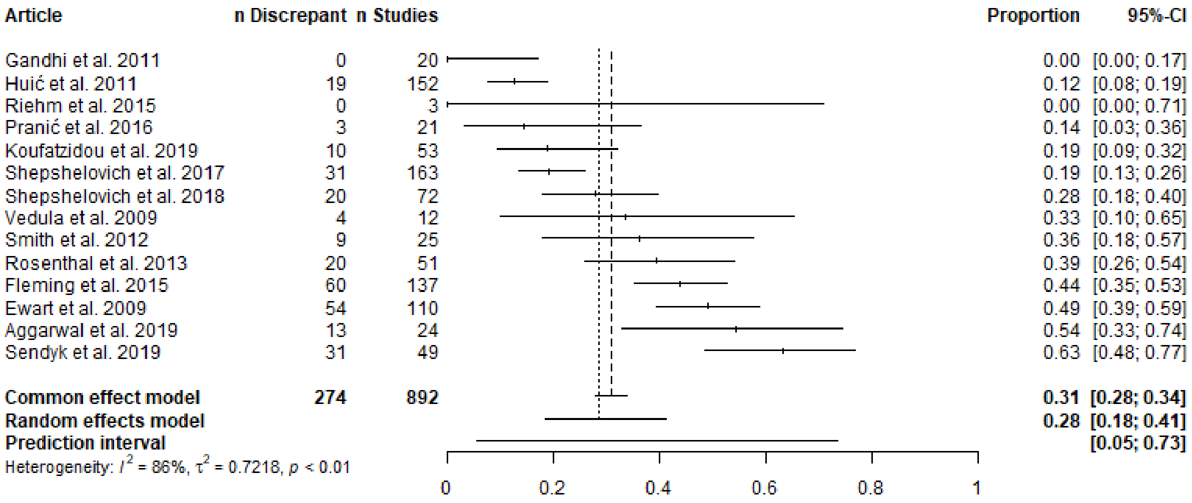


Supplementary Figure D19. Secondary outcome added, unclear registration timing.

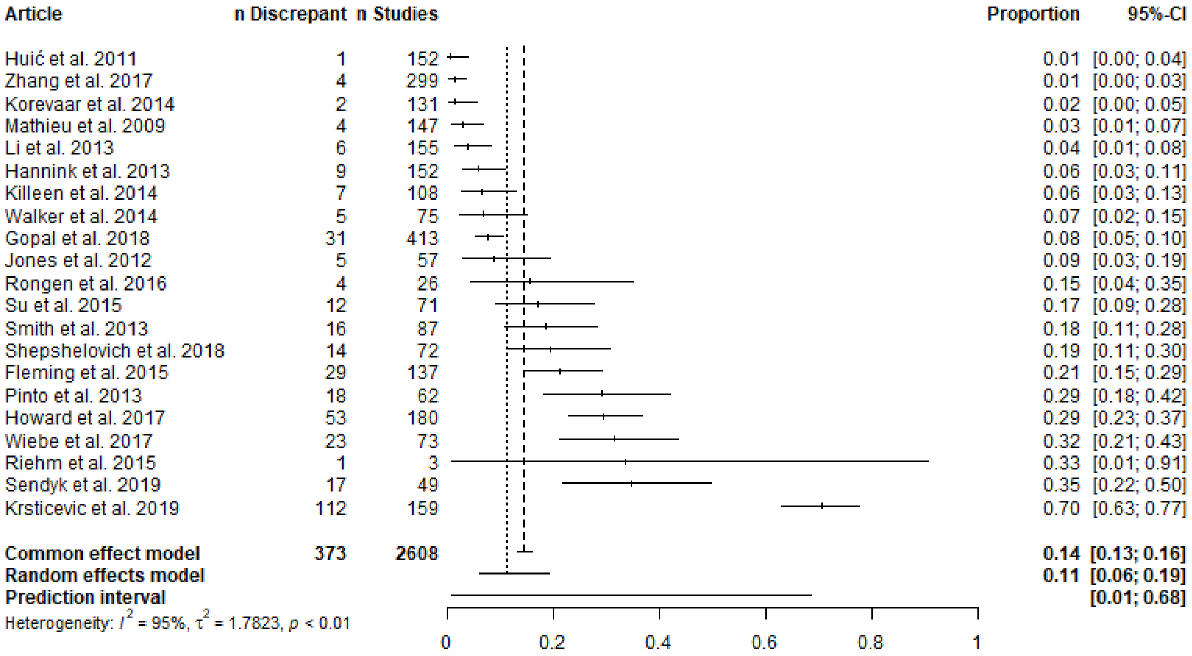


Supplementary Figure D20. Timing of outcome measurement changed, unclear registration timing.

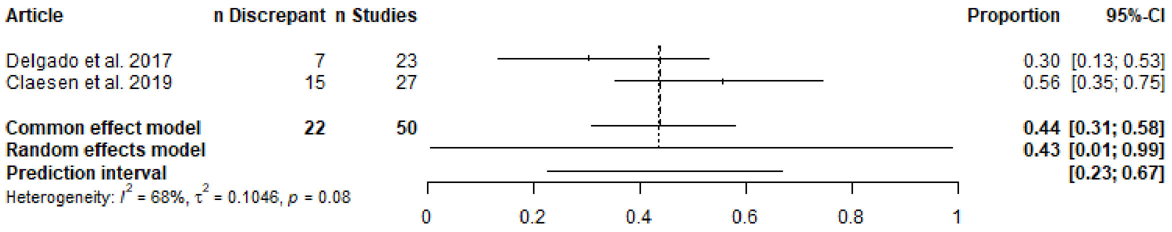


Supplementary Figure D21. Eligibility criteria, prospective.

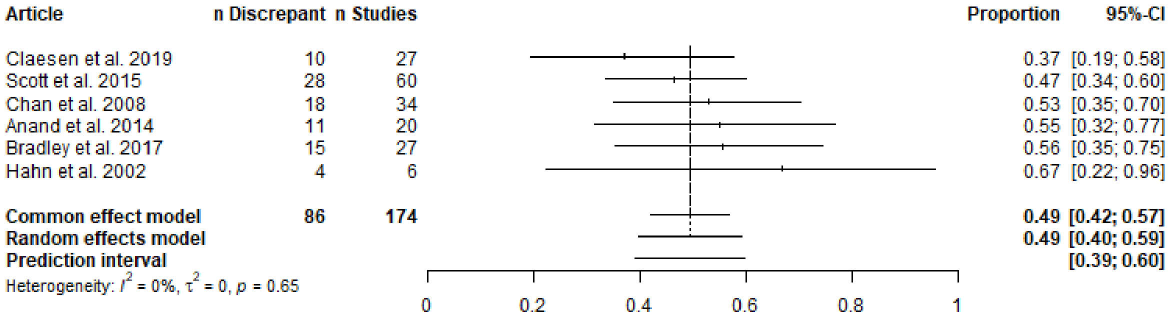


Supplementary Figure D22. Sample size, prospective.

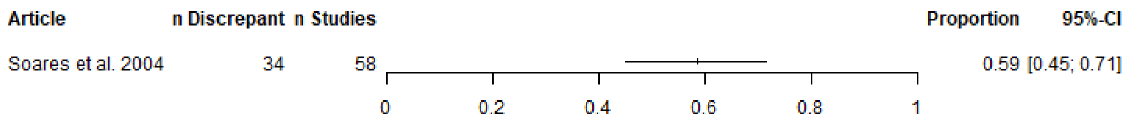


Supplementary Figure D23. Randomization, prospective.

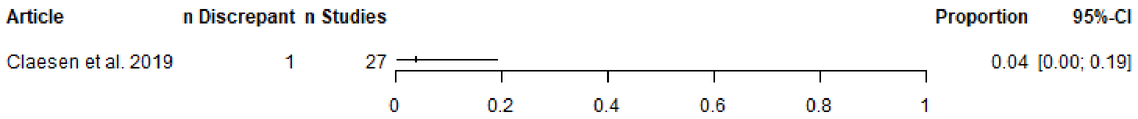


Supplementary Figure D24. Intervention, prospective.

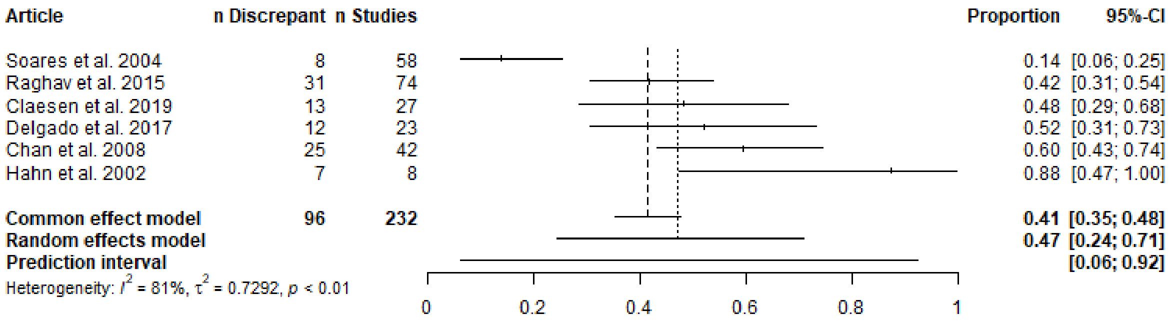


Supplementary Figure D25. Analyses, prospective.

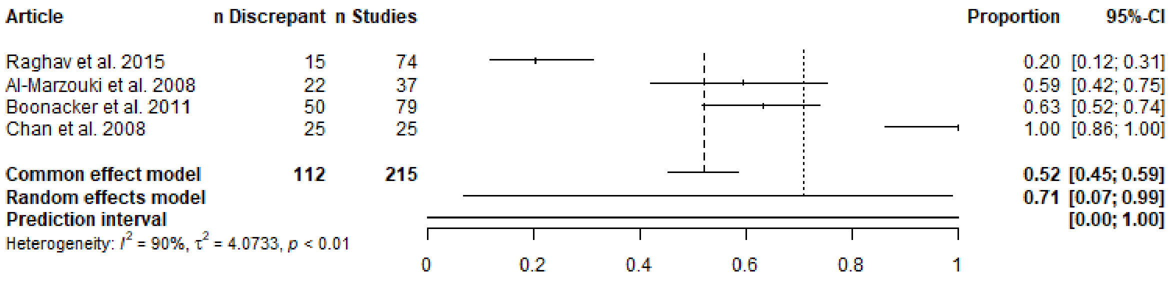


Supplementary Figure D26. Subgroup analyses, prospective.

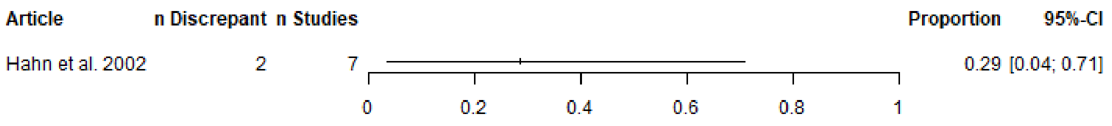


Supplementary Figure D27. Funding, prospective.

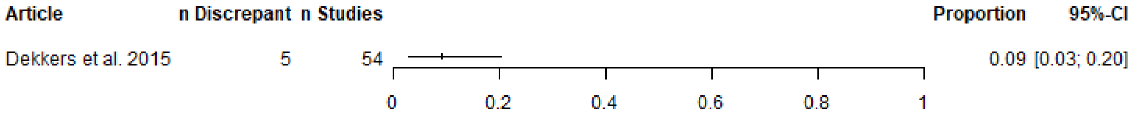


Supplementary Figure D28. Results, prospective.

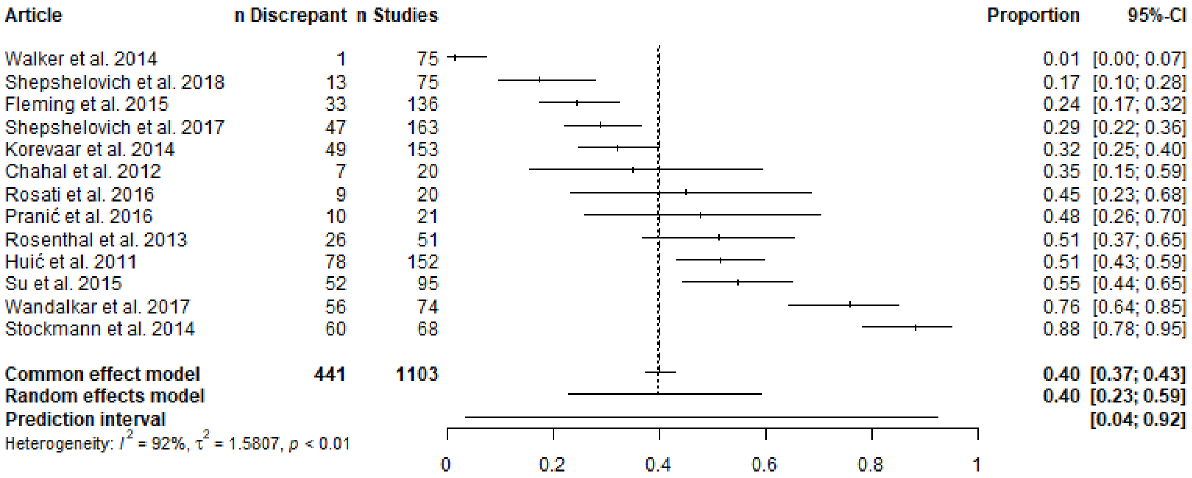


Supplementary Figure D29. Eligibility criteria, unclear registration timing.

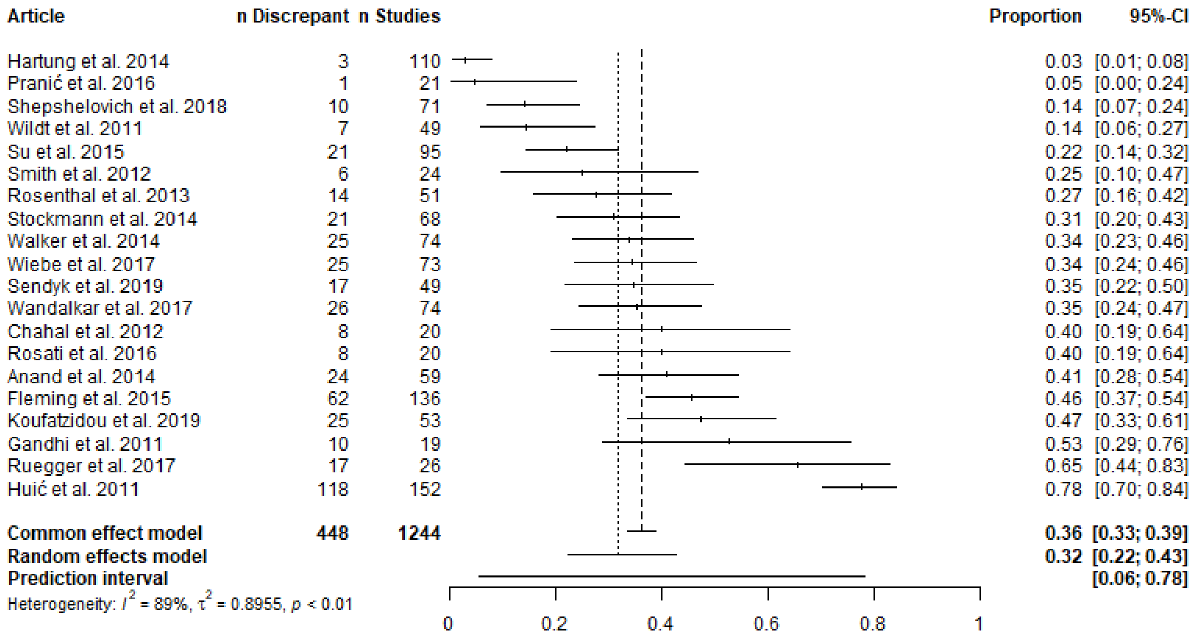


Supplementary Figure D30. Sample size, unclear registration timing.

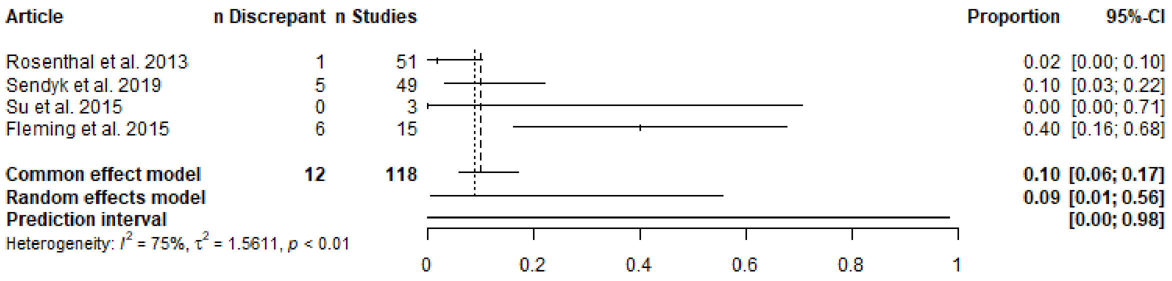


Supplementary Figure D31. Randomization, unclear registration timing.

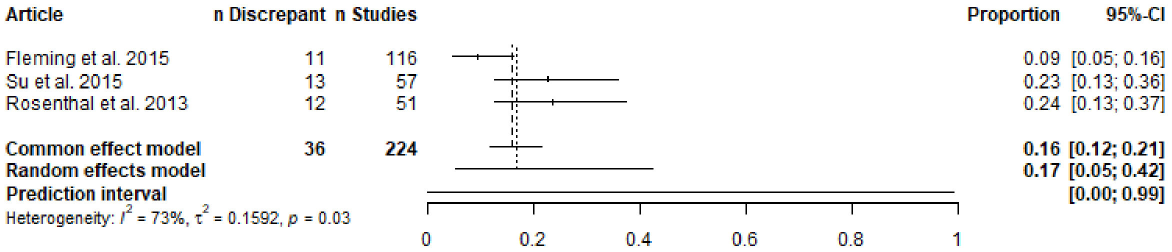


Supplementary Figure D32. Blinding, unclear registration timing.

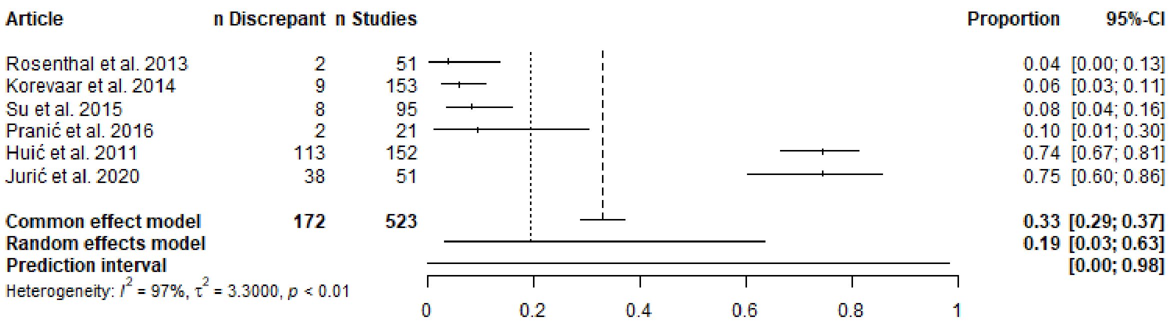


Supplementary Figure D33. Intervention, unclear registration timing.

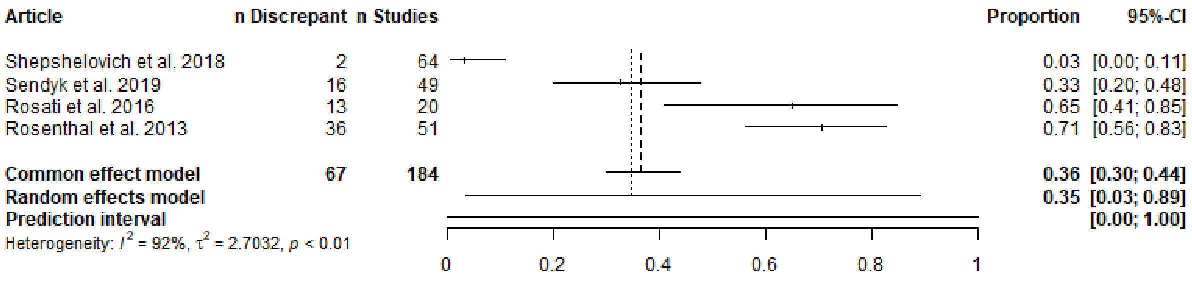


Supplementary Figure D34. Study duration, unclear registration timing.

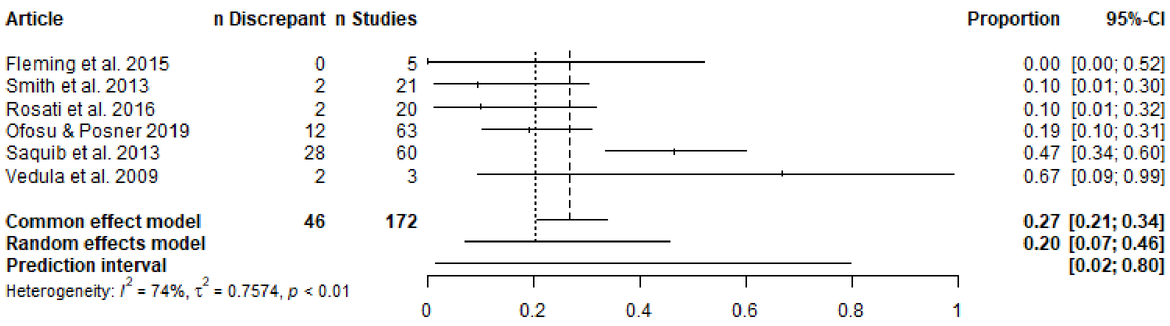


Supplementary Figure D35. Analyses, unclear registration timing.

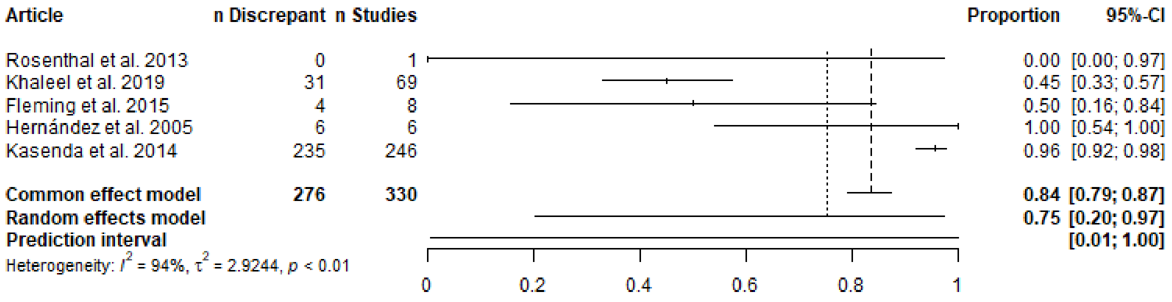


Supplementary Figure D36. Subgroup analyses, unclear registration timing.

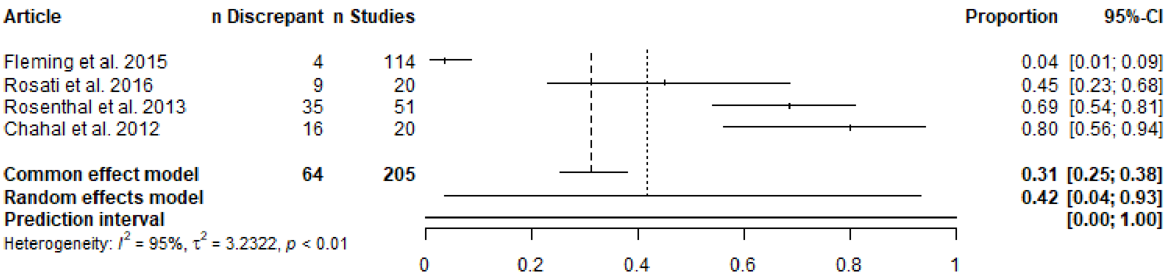


Supplementary Figure D37. Funding, unclear registration timing.

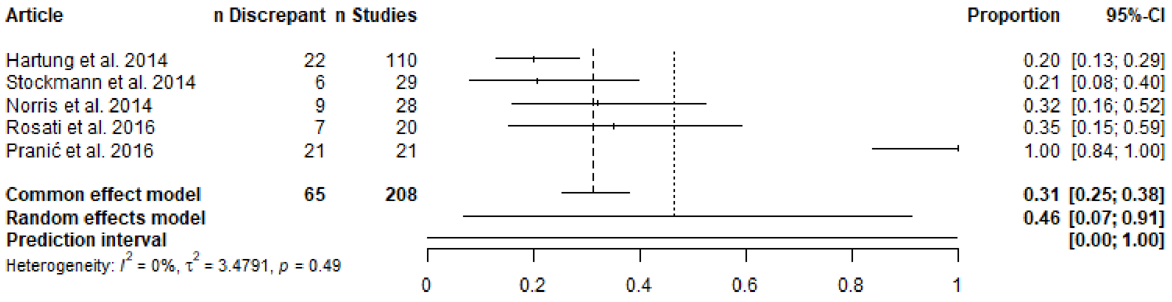


Supplementary Figure D38. Results, unclear registration timing.

### E. Meta-regression model summaries

##### Meta-regression sensitivity analysis

We ran the meta-regressions presented in the manuscript with one predictor at a time. In line with a suggestion from a reviewer, we also ran a meta-regression with all predictors at the same time. We had no a priori thoughts on how the predictors may interact and thus did include the interaction of predictors in our meta-regressions.

This meta-regression revealed 6 predictors with $p < 0.10$. The full output is appended below. Five of the predictors include between 1 to 4 articles in one category and thus may stem from idiosyncrasies of one or more individual studies, rather than from the predictor itself. As Cochrane advises against meta-regression when $k < 10$ (https://training.cochrane.org/handbook/current/chapter-13#section-13-3-5-6), we do not explore these findings further.

This meta-regression revealed one predictor of potential interest. Articles that compared publications to the most recent version of a registration ($k = 15$) may be less likely to have at least one primary outcome discrepancy than articles that compared publications to the original version of a registration ($k = 29$) ($p = 0.09$; OR 95% CI: 0.32-1.07).

```
##
## Mixed-Effects Model (k = 70; tau^2 estimator: ML)
##
## tau^2 (estimated amount of residual heterogeneity):     0.3664
## tau (square root of estimated tau^2 value):             0.6053
## I^2 (residual heterogeneity / unaccounted variability): 86.00%
## H^2 (unaccounted variability / sampling variability):   7.14
##
## Tests for Residual Heterogeneity:
## Wld(df = 50) = 342.2743, p-val < .0001
## LRT(df = 50) = 458.3887, p-val < .0001
##
## Test of Moderators (coefficients 2:20):
## F(df1 = 19, df2 = 50) = 1.3420, p-val = 0.2007
##
## Model Results:
##
##                                                                 estimate
## intrcpt                                                         -42.8460
## df1$pubYear                                                       0.0210
## df1$disciplineDentistry (DENT)                                    0.1016
## df1$disciplineEconomics, Econometrics and Finance (ECON)          0.4086
## df1$disciplineHealth Professions (HEAL)                           1.4988
## df1$disciplinePsychology (PSYC)                                   1.2335
## df1$sourceOther: ethics committee                                 0.4129
## df1$sourceOther: funder                                           2.4149
## df1$sourcereg                                                    -0.3632
## df1$sourcereg,journal                                             0.6233
## df1$sourcereg,searchEngine                                       -0.8627
## df1$sourcesearchEngine                                           -0.2628
## df1$versionMost recent version                                   -0.5331
## df1$versionNo                                                    -0.1020
## df1$versionOther                                                 -0.2049
## df1$disclosedexcludedDisclosedDiscrepancies                       0.3049
## df1$disclosedNo                                                   0.0273
```

```
## df1$disclosedYes, and NONE of the discrepancies were disclosed    0.2042
## df1$comparatorgrant                                              -2.3269
## df1$comparatorprotocol                                           -0.9981
##                                                                      se
## intrcpt                                                         62.0366
## df1$pubYear                                                      0.0308
## df1$disciplineDentistry (DENT)                                   0.4287
## df1$disciplineEconomics, Econometrics and Finance (ECON)         0.7221
## df1$disciplineHealth Professions (HEAL)                          0.7510
## df1$disciplinePsychology (PSYC)                                  0.6494
## df1$sourceOther: ethics committee                                0.5356
## df1$sourceOther: funder                                          1.0151
## df1$sourcereg                                                    0.2454
## df1$sourcereg,journal                                            0.7028
## df1$sourcereg,searchEngine                                       0.7230
## df1$sourcesearchEngine                                           0.2496
## df1$versionMost recent version                                   0.3005
## df1$versionNo                                                    0.2152
## df1$versionOther                                                 0.3291
## df1$disclosedexcludedDisclosedDiscrepancies                      0.4616
## df1$disclosedNo                                                  0.2301
## df1$disclosedYes, and NONE of the discrepancies were disclosed   0.3577
## df1$comparatorgrant                                              1.2765
## df1$comparatorprotocol                                           0.4604
##                                                                    tval  df
## intrcpt                                                         -0.6907  50
## df1$pubYear                                                      0.6828  50
## df1$disciplineDentistry (DENT)                                   0.2370  50
## df1$disciplineEconomics, Econometrics and Finance (ECON)         0.5659  50
## df1$disciplineHealth Professions (HEAL)                          1.9957  50
## df1$disciplinePsychology (PSYC)                                  1.8993  50
## df1$sourceOther: ethics committee                                0.7709  50
## df1$sourceOther: funder                                          2.3789  50
## df1$sourcereg                                                   -1.4799  50
## df1$sourcereg,journal                                            0.8869  50
## df1$sourcereg,searchEngine                                      -1.1933  50
## df1$sourcesearchEngine                                          -1.0527  50
## df1$versionMost recent version                                  -1.7741  50
## df1$versionNo                                                   -0.4740  50
## df1$versionOther                                                -0.6224  50
## df1$disclosedexcludedDisclosedDiscrepancies                      0.6604  50
## df1$disclosedNo                                                  0.1188  50
## df1$disclosedYes, and NONE of the discrepancies were disclosed   0.5709  50
## df1$comparatorgrant                                             -1.8229  50
## df1$comparatorprotocol                                          -2.1680  50
##                                                                   pval
## intrcpt                                                         0.4930
## df1$pubYear                                                     0.4979
## df1$disciplineDentistry (DENT)                                  0.8136
## df1$disciplineEconomics, Econometrics and Finance (ECON)        0.5740
## df1$disciplineHealth Professions (HEAL)                         0.0514
## df1$disciplinePsychology (PSYC)                                 0.0633
## df1$sourceOther: ethics committee                               0.4444
## df1$sourceOther: funder                                         0.0212
```

```
## df1$sourcereg                                           0.1452
## df1$sourcereg,journal                                   0.3794
## df1$sourcereg,searchEngine                              0.2384
## df1$sourcesearchEngine                                  0.2975
## df1$versionMost recent version                          0.0821
## df1$versionNo                                           0.6376
## df1$versionOther                                        0.5365
## df1$disclosedexcludedDisclosedDiscrepancies             0.5120
## df1$disclosedNo                                         0.9059
## df1$disclosedYes, and NONE of the discrepancies were disclosed  0.5706
## df1$comparatorgrant                                     0.0743
## df1$comparatorprotocol                                  0.0349
##                                                            ci.lb
## intrcpt                                                -167.4502
## df1$pubYear                                              -0.0408
## df1$disciplineDentistry (DENT)                           -0.7595
## df1$disciplineEconomics, Econometrics and Finance (ECON)     -1.0417
## df1$disciplineHealth Professions (HEAL)                  -0.0096
## df1$disciplinePsychology (PSYC)                          -0.0710
## df1$sourceOther: ethics committee                        -0.6629
## df1$sourceOther: funder                                   0.3760
## df1$sourcereg                                            -0.8561
## df1$sourcereg,journal                                    -0.7882
## df1$sourcereg,searchEngine                               -2.3149
## df1$sourcesearchEngine                                   -0.7641
## df1$versionMost recent version                           -1.1366
## df1$versionNo                                            -0.5343
## df1$versionOther                                         -0.8659
## df1$disclosedexcludedDisclosedDiscrepancies              -0.6224
## df1$disclosedNo                                          -0.4348
## df1$disclosedYes, and NONE of the discrepancies were disclosed    -0.5142
## df1$comparatorgrant                                      -4.8908
## df1$comparatorprotocol                                   -1.9227
##                                                           ci.ub
## intrcpt                                                 81.7581
## df1$pubYear                                              0.0829
## df1$disciplineDentistry (DENT)                           0.9626
## df1$disciplineEconomics, Econometrics and Finance (ECON)      1.8589
## df1$disciplineHealth Professions (HEAL)                  3.0072  .
## df1$disciplinePsychology (PSYC)                          2.5379  .
## df1$sourceOther: ethics committee                        1.4888
## df1$sourceOther: funder                                  4.4538  *
## df1$sourcereg                                            0.1297
## df1$sourcereg,journal                                    2.0348
## df1$sourcereg,searchEngine                               0.5895
## df1$sourcesearchEngine                                   0.2386
## df1$versionMost recent version                           0.0704  .
## df1$versionNo                                            0.3303
## df1$versionOther                                         0.4562
## df1$disclosedexcludedDisclosedDiscrepancies              1.2321
## df1$disclosedNo                                          0.4895
## df1$disclosedYes, and NONE of the discrepancies were disclosed   0.9227
## df1$comparatorgrant                                      0.2370  .
## df1$comparatorprotocol                                  -0.0734  *
```

```
##
## ---
## Signif. codes:  0 '***' 0.001 '**' 0.01 '*' 0.05 '.' 0.1 ' ' 1
```

**Discipline, primary outcomes**

```
##
## Mixed-Effects Model (k = 70; tau^2 estimator: ML)
##
## tau^2 (estimated amount of residual heterogeneity):     0.4973
## tau (square root of estimated tau^2 value):             0.7052
## I^2 (residual heterogeneity / unaccounted variability): 89.80%
## H^2 (unaccounted variability / sampling variability):   9.80
##
## Tests for Residual Heterogeneity:
## Wld(df = 65) = 484.2655, p-val < .0001
## LRT(df = 65) = 619.5987, p-val < .0001
##
## Test of Moderators (coefficients 2:5):
## F(df1 = 4, df2 = 65) = 1.3093, p-val = 0.2759
##
## Model Results:
##
##                                                          estimate      se
## intrcpt                                                   -0.7695  0.0982
## df1$disciplineDentistry (DENT)                             0.1419  0.4466
## df1$disciplineEconomics, Econometrics and Finance (ECON)   0.3052  0.7434
## df1$disciplineHealth Professions (HEAL)                    0.9665  0.7569
## df1$disciplinePsychology (PSYC)                            1.1512  0.6119
##                                                             tval  df    pval
## intrcpt                                                  -7.8374  65  <.0001
## df1$disciplineDentistry (DENT)                            0.3178  65  0.7517
## df1$disciplineEconomics, Econometrics and Finance (ECON)  0.4106  65  0.6827
## df1$disciplineHealth Professions (HEAL)                   1.2768  65  0.2062
## df1$disciplinePsychology (PSYC)                           1.8813  65  0.0644
##                                                            ci.lb    ci.ub
## intrcpt                                                  -0.9656  -0.5734  ***
## df1$disciplineDentistry (DENT)                           -0.7501   1.0339
## df1$disciplineEconomics, Econometrics and Finance (ECON) -1.1795   1.7900
## df1$disciplineHealth Professions (HEAL)                  -0.5452   2.4782
## df1$disciplinePsychology (PSYC)                          -0.0709   2.3733    .
##
## ---
## Signif. codes:  0 '***' 0.001 '**' 0.01 '*' 0.05 '.' 0.1 ' ' 1
```

**Comparator, primary outcomes**

```
##
## Mixed-Effects Model (k = 70; tau^2 estimator: ML)
##
## tau^2 (estimated amount of residual heterogeneity):     0.5031
## tau (square root of estimated tau^2 value):             0.7093
## I^2 (residual heterogeneity / unaccounted variability): 89.96%
## H^2 (unaccounted variability / sampling variability):   9.96
##
## Tests for Residual Heterogeneity:
## Wld(df = 66) = 439.1139, p-val < .0001
## LRT(df = 66) = 546.7582, p-val < .0001
##
## Test of Moderators (coefficients 2:4):
## F(df1 = 3, df2 = 66) = 0.8637, p-val = 0.4644
##
## Model Results:
##
##                                     estimate      se     tval  df    pval
## intrcpt                              -0.6892  0.0989  -6.9690  66  <.0001
## droplevels(df1$comparator)ethics      0.1085  0.4982   0.2177  66  0.8283
## droplevels(df1$comparator)grant       0.2585  0.7757   0.3333  66  0.7400
## droplevels(df1$comparator)protocol   -0.6395  0.4155  -1.5389  66  0.1286
##                                       ci.lb    ci.ub
## intrcpt                             -0.8866  -0.4917  ***
## droplevels(df1$comparator)ethics    -0.8862   1.1031
## droplevels(df1$comparator)grant     -1.2902   1.8072
## droplevels(df1$comparator)protocol  -1.4691   0.1902
##
## ---
## Signif. codes:  0 '***' 0.001 '**' 0.01 '*' 0.05 '.' 0.1 ' ' 1
```

**Source, primary outcomes**

```
##
## Mixed-Effects Model (k = 70; tau^2 estimator: ML)
##
## tau^2 (estimated amount of residual heterogeneity):     0.5197
## tau (square root of estimated tau^2 value):             0.7209
## I^2 (residual heterogeneity / unaccounted variability): 89.95%
## H^2 (unaccounted variability / sampling variability):   9.95
##
## Tests for Residual Heterogeneity:
## Wld(df = 63) = 491.6691, p-val < .0001
## LRT(df = 63) = 629.6278, p-val < .0001
##
## Test of Moderators (coefficients 2:7):
## F(df1 = 6, df2 = 63) = 0.6989, p-val = 0.6515
##
## Model Results:
##
##                                                 estimate      se     tval  df
## intrcpt                                          -0.6406  0.1456  -4.3983  63
## droplevels(df1$source)Other: ethics committee     0.0578  0.5158   0.1121  63
## droplevels(df1$source)Other: funder               0.6779  0.6178   1.0973  63
## droplevels(df1$source)reg                        -0.3005  0.2346  -1.2808  63
## droplevels(df1$source)reg,journal                 0.1826  0.7542   0.2421  63
## droplevels(df1$source)reg,searchEngine           -0.6840  0.7832  -0.8734  63
## droplevels(df1$source)searchEngine               -0.0312  0.2515  -0.1239  63
##                                                    pval    ci.lb    ci.ub
## intrcpt                                          <.0001  -0.9316  -0.3495  ***
## droplevels(df1$source)Other: ethics committee    0.9111  -0.9729   1.0886
## droplevels(df1$source)Other: funder              0.2767  -0.5566   1.9124
## droplevels(df1$source)reg                        0.2050  -0.7693   0.1684
## droplevels(df1$source)reg,journal                0.8095  -1.3245   1.6897
## droplevels(df1$source)reg,searchEngine           0.3858  -2.2491   0.8810
## droplevels(df1$source)searchEngine               0.9018  -0.5338   0.4715
##
## ---
## Signif. codes:  0 '***' 0.001 '**' 0.01 '*' 0.05 '.' 0.1 ' ' 1
```

**Version, primary outcomes**

```
##
## Mixed-Effects Model (k = 70; tau^2 estimator: ML)
##
## tau^2 (estimated amount of residual heterogeneity):     0.5251
## tau (square root of estimated tau^2 value):             0.7247
## I^2 (residual heterogeneity / unaccounted variability): 90.05%
## H^2 (unaccounted variability / sampling variability):   10.05
##
## Tests for Residual Heterogeneity:
## Wld(df = 66) = 485.3920, p-val < .0001
## LRT(df = 66) = 612.4631, p-val < .0001
##
## Test of Moderators (coefficients 2:4):
## F(df1 = 3, df2 = 66) = 0.3719, p-val = 0.7735
##
## Model Results:
##
##                                 estimate      se     tval  df    pval    ci.lb
## intrcpt                          -0.6048  0.1599  -3.7832  66  0.0003  -0.9239
## df1$versionMost recent version   -0.2477  0.2642  -0.9376  66  0.3519  -0.7752
## df1$versionNo                    -0.1046  0.2299  -0.4551  66  0.6505  -0.5636
## df1$versionOther                 -0.2433  0.3301  -0.7372  66  0.4636  -0.9023
##                                   ci.ub
## intrcpt                         -0.2856  ***
## df1$versionMost recent version   0.2798
## df1$versionNo                    0.3543
## df1$versionOther                 0.4157
##
## ---
## Signif. codes:  0 '***' 0.001 '**' 0.01 '*' 0.05 '.' 0.1 ' ' 1
```

##### Disclosed, primary outcomes

```
##
## Mixed-Effects Model (k = 70; tau^2 estimator: ML)
##
## tau^2 (estimated amount of residual heterogeneity):     0.5403
## tau (square root of estimated tau^2 value):             0.7350
## I^2 (residual heterogeneity / unaccounted variability): 90.43%
## H^2 (unaccounted variability / sampling variability):   10.45
##
## Tests for Residual Heterogeneity:
## Wld(df = 66) = 494.1488, p-val < .0001
## LRT(df = 66) = 644.7748, p-val < .0001
##
## Test of Moderators (coefficients 2:4):
## F(df1 = 3, df2 = 66) = 0.0739, p-val = 0.9738
##
## Model Results:
##
##                                                                 estimate
## intrcpt                                                          -0.7709
## df1$disclosedexcludedDisclosedDiscrepancies                       0.2332
## df1$disclosedNo                                                   0.0600
## df1$disclosedYes, and NONE of the discrepancies were disclosed    0.0611
##                                                                     se     tval
## intrcpt                                                         0.2012  -3.8314
## df1$disclosedexcludedDisclosedDiscrepancies                     0.5094   0.4578
## df1$disclosedNo                                                 0.2362   0.2541
## df1$disclosedYes, and NONE of the discrepancies were disclosed  0.3419   0.1788
##                                                                 df    pval
## intrcpt                                                         66  0.0003
## df1$disclosedexcludedDisclosedDiscrepancies                     66  0.6486
## df1$disclosedNo                                                 66  0.8002
## df1$disclosedYes, and NONE of the discrepancies were disclosed  66  0.8587
##                                                                   ci.lb
## intrcpt                                                         -1.1725
## df1$disclosedexcludedDisclosedDiscrepancies                     -0.7838
## df1$disclosedNo                                                 -0.4115
## df1$disclosedYes, and NONE of the discrepancies were disclosed  -0.6215
##                                                                   ci.ub
## intrcpt                                                         -0.3692  ***
## df1$disclosedexcludedDisclosedDiscrepancies                      1.2502
## df1$disclosedNo                                                  0.5315
## df1$disclosedYes, and NONE of the discrepancies were disclosed   0.7437
##
## ---
## Signif. codes:  0 '***' 0.001 '**' 0.01 '*' 0.05 '.' 0.1 ' ' 1
```

**Publication year, primary outcomes**

```
##
## Mixed-Effects Model (k = 70; tau^2 estimator: ML)
##
## tau^2 (estimated amount of residual heterogeneity):     0.5405
## tau (square root of estimated tau^2 value):             0.7352
## I^2 (residual heterogeneity / unaccounted variability): 90.44%
## H^2 (unaccounted variability / sampling variability):   10.46
##
## Tests for Residual Heterogeneity:
## Wld(df = 68) = 505.5885, p-val < .0001
## LRT(df = 68) = 647.2738, p-val < .0001
##
## Test of Moderators (coefficient 2):
## F(df1 = 1, df2 = 68) = 0.0438, p-val = 0.8348
##
## Model Results:
##
##              estimate       se     tval  df    pval      ci.lb    ci.ub
## intrcpt      -11.8783  53.3266  -0.2227  68  0.8244  -118.2899  94.5332
## df1$pubYear    0.0055   0.0265   0.2093  68  0.8348    -0.0473   0.0584
##
## ---
## Signif. codes:  0 '***' 0.001 '**' 0.01 '*' 0.05 '.' 0.1 ' ' 1
```

**Discpline binary, primary outcomes**

```
##
## Mixed-Effects Model (k = 70; tau^2 estimator: ML)
##
## tau^2 (estimated amount of residual heterogeneity):     0.5175
## tau (square root of estimated tau^2 value):             0.7194
## I^2 (residual heterogeneity / unaccounted variability): 90.11%
## H^2 (unaccounted variability / sampling variability):   10.11
##
## Tests for Residual Heterogeneity:
## Wld(df = 68) = 497.9543, p-val < .0001
## LRT(df = 68) = 634.4749, p-val < .0001
##
## Test of Moderators (coefficient 2):
## F(df1 = 1, df2 = 68) = 2.8890, p-val = 0.0938
##
## Model Results:
##
##                                 estimate      se     tval  df    pval    ci.lb
## intrcpt                          -0.7703  0.0999  -7.7124  68  <.0001  -0.9696
## dfDiscipline1$disciplineother     0.5342  0.3143   1.6997  68  0.0938  -0.0929
##                                   ci.ub
## intrcpt                         -0.5710  ***
## dfDiscipline1$disciplineother    1.1613    .
##
## ---
## Signif. codes:  0 '***' 0.001 '**' 0.01 '*' 0.05 '.' 0.1 ' ' 1
```

##### Comparator binary, primary outcomes

```
##
## Mixed-Effects Model (k = 70; tau^2 estimator: ML)
##
## tau^2 (estimated amount of residual heterogeneity):     0.5305
## tau (square root of estimated tau^2 value):             0.7284
## I^2 (residual heterogeneity / unaccounted variability): 90.37%
## H^2 (unaccounted variability / sampling variability):   10.38
##
## Tests for Residual Heterogeneity:
## Wld(df = 68) = 491.3839, p-val < .0001
## LRT(df = 68) = 609.5590, p-val < .0001
##
## Test of Moderators (coefficient 2):
## F(df1 = 1, df2 = 68) = 0.6521, p-val = 0.4222
##
## Model Results:
##
##                              estimate      se     tval  df    pval    ci.lb
## intrcpt                       -0.9418  0.2945  -3.1978  68  0.0021  -1.5296
## df1Comparator$comparatorreg    0.2516  0.3115   0.8076  68  0.4222  -0.3701
##                                 ci.ub
## intrcpt                       -0.3541  **
## df1Comparator$comparatorreg    0.8733
##
## ---
## Signif. codes:  0 '***' 0.001 '**' 0.01 '*' 0.05 '.' 0.1 ' ' 1
```

**Discipline, secondary outcomes**

```
##
## Mixed-Effects Model (k = 22; tau^2 estimator: ML)
##
## tau^2 (estimated amount of residual heterogeneity):     1.1358
## tau (square root of estimated tau^2 value):             1.0658
## I^2 (residual heterogeneity / unaccounted variability): 92.81%
## H^2 (unaccounted variability / sampling variability):   13.91
##
## Tests for Residual Heterogeneity:
## Wld(df = 19) = 152.4015, p-val < .0001
## LRT(df = 19) = 232.6890, p-val < .0001
##
## Test of Moderators (coefficients 2:3):
## F(df1 = 2, df2 = 19) = 1.3121, p-val = 0.2926
##
## Model Results:
##
##                                          estimate      se     tval  df    pval
## intrcpt                                    0.5830  0.2642   2.2067  19  0.0398
## df2$disciplineDentistry (DENT)            -0.9288  0.8150  -1.1396  19  0.2686
## df2$disciplineHealth Professions (HEAL)    1.2320  1.1580   1.0639  19  0.3007
##                                            ci.lb   ci.ub
## intrcpt                                   0.0300  1.1359  *
## df2$disciplineDentistry (DENT)           -2.6346  0.7771
## df2$disciplineHealth Professions (HEAL)  -1.1917  3.6557
##
## ---
## Signif. codes:  0 '***' 0.001 '**' 0.01 '*' 0.05 '.' 0.1 ' ' 1
```

**Comparator, secondary outcomes**

```
##
## Mixed-Effects Model (k = 22; tau^2 estimator: ML)
##
## tau^2 (estimated amount of residual heterogeneity):     1.1341
## tau (square root of estimated tau^2 value):             1.0650
## I^2 (residual heterogeneity / unaccounted variability): 93.39%
## H^2 (unaccounted variability / sampling variability):   15.12
##
## Tests for Residual Heterogeneity:
## Wld(df = 20) = 188.4912, p-val < .0001
## LRT(df = 20) = 268.8685, p-val < .0001
##
## Test of Moderators (coefficient 2):
## F(df1 = 1, df2 = 20) = 3.4451, p-val = 0.0782
##
## Model Results:
##
##                                    estimate      se    tval  df    pval
## intrcpt                              0.4164  0.2535  1.6425  20  0.1161
## droplevels(df2$comparator)protocol    1.7751  0.9564  1.8561  20  0.0782
##                                       ci.lb   ci.ub
## intrcpt                             -0.1124  0.9452
## droplevels(df2$comparator)protocol  -0.2198  3.7700  .
##
## ---
## Signif. codes:  0 '***' 0.001 '**' 0.01 '*' 0.05 '.' 0.1 ' ' 1
```

**Source, secondary outcomes**

```
##
## Mixed-Effects Model (k = 22; tau^2 estimator: ML)
##
## tau^2 (estimated amount of residual heterogeneity):     0.6838
## tau (square root of estimated tau^2 value):             0.8269
## I^2 (residual heterogeneity / unaccounted variability): 88.00%
## H^2 (unaccounted variability / sampling variability):   8.33
##
## Tests for Residual Heterogeneity:
## Wld(df = 16) = 135.1594, p-val < .0001
## LRT(df = 16) = 187.0805, p-val < .0001
##
## Test of Moderators (coefficients 2:6):
## F(df1 = 5, df2 = 16) = 3.3793, p-val = 0.0284
##
## Model Results:
##
##                             estimate      se     tval  df    pval    ci.lb
## intrcpt                      -0.3517  0.3203  -1.0981  16  0.2884  -1.0306
## df2$sourcejournal             1.6186  0.4626   3.4989  16  0.0030   0.6379
## df2$sourceOther: funder       2.9203  1.4041   2.0798  16  0.0540  -0.0563
## df2$sourcereg,journal         0.9517  0.9029   1.0540  16  0.3075  -0.9624
## df2$sourcereg,searchEngine    2.1280  0.9400   2.2639  16  0.0378   0.1353
## df2$sourcesearchEngine        0.5912  0.6049   0.9774  16  0.3429  -0.6911
##                              ci.ub
## intrcpt                     0.3272
## df2$sourcejournal           2.5993  **
## df2$sourceOther: funder     5.8968   .
## df2$sourcereg,journal       2.8658
## df2$sourcereg,searchEngine  4.1206   *
## df2$sourcesearchEngine      1.8735
##
## ---
## Signif. codes:  0 '***' 0.001 '**' 0.01 '*' 0.05 '.' 0.1 ' ' 1
```

**Version, secondary outcomes**

```
##
## Mixed-Effects Model (k = 22; tau^2 estimator: ML)
##
## tau^2 (estimated amount of residual heterogeneity):     1.2281
## tau (square root of estimated tau^2 value):             1.1082
## I^2 (residual heterogeneity / unaccounted variability): 93.15%
## H^2 (unaccounted variability / sampling variability):   14.60
##
## Tests for Residual Heterogeneity:
## Wld(df = 18) = 164.5429, p-val < .0001
## LRT(df = 18) = 247.8955, p-val < .0001
##
## Test of Moderators (coefficients 2:4):
## F(df1 = 3, df2 = 18) = 0.3299, p-val = 0.8038
##
## Model Results:
##
##                                estimate      se     tval  df    pval    ci.lb
## intrcpt                          0.3831  0.4475   0.8560  18  0.4033  -0.5571
## df2$versionMost recent version   0.5515  0.6480   0.8512  18  0.4058  -0.8098
## df2$versionNo                   -0.0300  0.6500  -0.0462  18  0.9637  -1.3956
## df2$versionOther                 0.1605  0.9210   0.1743  18  0.8636  -1.7744
##                                 ci.ub
## intrcpt                        1.3233
## df2$versionMost recent version 1.9128
## df2$versionNo                  1.3356
## df2$versionOther               2.0954
##
## ---
## Signif. codes:  0 '***' 0.001 '**' 0.01 '*' 0.05 '.' 0.1 ' ' 1
```

**Disclosed, secondary outcomes**

```
##
## Mixed-Effects Model (k = 22; tau^2 estimator: ML)
##
## tau^2 (estimated amount of residual heterogeneity):     1.2148
## tau (square root of estimated tau^2 value):             1.1022
## I^2 (residual heterogeneity / unaccounted variability): 93.11%
## H^2 (unaccounted variability / sampling variability):   14.52
##
## Tests for Residual Heterogeneity:
## Wld(df = 18) = 173.0004, p-val < .0001
## LRT(df = 18) = 241.2204, p-val < .0001
##
## Test of Moderators (coefficients 2:4):
## F(df1 = 3, df2 = 18) = 0.5040, p-val = 0.6843
##
## Model Results:
##
##                                                             estimate
## intrcpt                                                       0.2683
## df2$disclosedexcludedDisclosedDiscrepancies                   0.7064
## df2$disclosedNo                                               0.3885
## df2$disclosedYes, and NONE of the discrepancies were disclosed  -0.5224
##                                                                 se     tval
## intrcpt                                                     0.6643   0.4039
## df2$disclosedexcludedDisclosedDiscrepancies                 1.0390   0.6799
## df2$disclosedNo                                             0.7324   0.5304
## df2$disclosedYes, and NONE of the discrepancies were disclosed  1.0664  -0.4899
##                                                             df    pval
## intrcpt                                                     18  0.6910
## df2$disclosedexcludedDisclosedDiscrepancies                 18  0.5052
## df2$disclosedNo                                             18  0.6023
## df2$disclosedYes, and NONE of the discrepancies were disclosed  18  0.6301
##                                                               ci.lb   ci.ub
## intrcpt                                                     -1.1273  1.6639
## df2$disclosedexcludedDisclosedDiscrepancies                 -1.4765  2.8894
## df2$disclosedNo                                             -1.1503  1.9272
## df2$disclosedYes, and NONE of the discrepancies were disclosed  -2.7627  1.7179
##
## intrcpt
## df2$disclosedexcludedDisclosedDiscrepancies
## df2$disclosedNo
## df2$disclosedYes, and NONE of the discrepancies were disclosed
##
## ---
## Signif. codes:  0 '***' 0.001 '**' 0.01 '*' 0.05 '.' 0.1 ' ' 1
```

**Publication year, secondary outcomes**

```
##
## Mixed-Effects Model (k = 22; tau^2 estimator: ML)
##
## tau^2 (estimated amount of residual heterogeneity):     1.3181
## tau (square root of estimated tau^2 value):             1.1481
## I^2 (residual heterogeneity / unaccounted variability): 93.94%
## H^2 (unaccounted variability / sampling variability):   16.51
##
## Tests for Residual Heterogeneity:
## Wld(df = 20) = 199.2193, p-val < .0001
## LRT(df = 20) = 284.0615, p-val < .0001
##
## Test of Moderators (coefficient 2):
## F(df1 = 1, df2 = 20) = 0.1305, p-val = 0.7217
##
## Model Results:
##
##              estimate        se     tval  df    pval      ci.lb     ci.ub
## intrcpt      -55.6213  155.4756  -0.3577  20  0.7243  -379.9378  268.6951
## df2$pubYear    0.0279    0.0772   0.3613  20  0.7217    -0.1331    0.1889
##
## ---
## Signif. codes:  0 '***' 0.001 '**' 0.01 '*' 0.05 '.' 0.1 ' ' 1
```

**F. Forest plots for all meta-analyses in Table 2 of the manuscript (outcome discrepancies)**

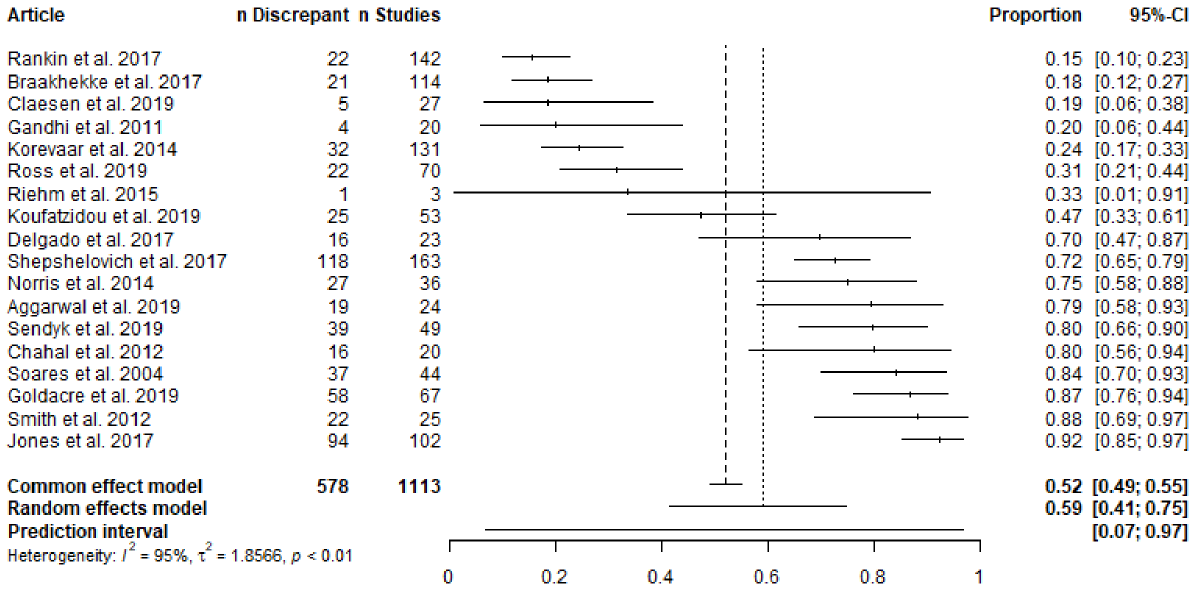


Supplementary Figure F1. Any outcome discrepancy.

| Article | n Discrepant | n Studies | Proportion | 95%-CI |
|---|---|---|---|---|
| Wildt et al. 2011 | 0 | 66 | 0.00 | [0.00; 0.05] |
| Zhang et al. 2017 | 25 | 299 | 0.08 | [0.05; 0.12] |
| Stockmann et al. 2014 | 6 | 68 | 0.09 | [0.03; 0.18] |
| Gandhi et al. 2011 | 2 | 20 | 0.10 | [0.01; 0.32] |
| Raghav et al. 2015 | 9 | 74 | 0.12 | [0.06; 0.22] |
| White et al. 2019 | 0 | 3 | 0.00 | [0.00; 0.71] |
| You et al. 2012 | 19 | 134 | 0.14 | [0.09; 0.21] |
| Li et al. 2013 | 22 | 155 | 0.14 | [0.09; 0.21] |
| Hartung et al. 2014 | 16 | 110 | 0.15 | [0.09; 0.23] |
| Chan et al. 2017 | 9 | 58 | 0.16 | [0.07; 0.27] |
| Khan et al. 2012 | 1 | 6 | 0.17 | [0.00; 0.64] |
| Delgado et al. 2017 | 39 | 216 | 0.18 | [0.13; 0.24] |
| Fleming et al. 2015 | 25 | 137 | 0.18 | [0.12; 0.26] |
| Braakhekke et al. 2017 | 21 | 114 | 0.18 | [0.12; 0.27] |
| Goldacre et al. 2019 | 13 | 67 | 0.19 | [0.11; 0.31] |
| Koufatzidou et al. 2019 | 11 | 53 | 0.21 | [0.11; 0.34] |
| Delgado et al. 2017 | 5 | 23 | 0.22 | [0.07; 0.44] |
| Vera-badillo et al. 2013 | 7 | 30 | 0.23 | [0.10; 0.42] |
| Thomas et al. 2016 | 4 | 16 | 0.25 | [0.07; 0.52] |
| Anand et al. 2014 | 15 | 59 | 0.25 | [0.15; 0.38] |
| Jones et al. 2018 | 27 | 103 | 0.26 | [0.18; 0.36] |
| Shepshelovich et al. 2018 | 19 | 72 | 0.26 | [0.17; 0.38] |
| Gopal et al. 2018 | 109 | 413 | 0.26 | [0.22; 0.31] |
| Shinohara et al. 2015 | 12 | 43 | 0.28 | [0.15; 0.44] |
| Calméjane et al. 2018 | 77 | 274 | 0.28 | [0.23; 0.34] |
| Scott et al. 2015 | 17 | 60 | 0.28 | [0.17; 0.41] |
| Pranić et al. 2016 | 6 | 21 | 0.29 | [0.11; 0.52] |
| Aggarwal et al. 2019 | 7 | 24 | 0.29 | [0.13; 0.51] |
| Killeen et al. 2014 | 32 | 108 | 0.30 | [0.21; 0.39] |
| Al-Marzouki et al. 2008 | 11 | 37 | 0.30 | [0.16; 0.47] |
| Smith et al. 2013 | 26 | 87 | 0.30 | [0.21; 0.41] |
| Ewart et al. 2009 | 34 | 110 | 0.31 | [0.22; 0.40] |
| Mathieu et al. 2009 | 46 | 147 | 0.31 | [0.24; 0.39] |
| Pandis et al. 2015 | 49 | 152 | 0.32 | [0.25; 0.40] |
| Tricco et al. 2016 | 31 | 96 | 0.32 | [0.23; 0.43] |
| Mathieu et al. 2012 | 12 | 37 | 0.32 | [0.18; 0.50] |
| Hahn et al. 2002 | 2 | 6 | 0.33 | [0.04; 0.78] |
| Walker et al. 2014 | 25 | 75 | 0.33 | [0.23; 0.45] |
| Chen et al. 2019 | 130 | 389 | 0.33 | [0.29; 0.38] |
| Chahal et al. 2012 | 7 | 20 | 0.35 | [0.15; 0.59] |
| Shepshelovich et al. 2017 | 62 | 163 | 0.38 | [0.31; 0.46] |
| Ofosu & Posner 2019 | 36 | 93 | 0.39 | [0.29; 0.49] |
| Huić et al. 2011 | 59 | 152 | 0.39 | [0.31; 0.47] |
| Won et al. 2019 | 25 | 64 | 0.39 | [0.27; 0.52] |
| Bonnot et al. 2016 | 34 | 87 | 0.39 | [0.29; 0.50] |
| Dwan et al. 2013 | 18 | 46 | 0.39 | [0.25; 0.55] |
| Chan et al. 2004 | 19 | 48 | 0.40 | [0.26; 0.55] |
| Su et al. 2015 | 32 | 71 | 0.45 | [0.33; 0.57] |
| Jones et al. 2017 | 46 | 102 | 0.45 | [0.35; 0.55] |
| Rosenthal et al. 2013 | 23 | 51 | 0.45 | [0.31; 0.60] |
| Jones et al. 2012 | 26 | 57 | 0.46 | [0.32; 0.59] |
| Wiebe et al. 2017 | 34 | 73 | 0.47 | [0.35; 0.59] |
| Krsticevic et al. 2019 | 77 | 159 | 0.48 | [0.40; 0.56] |
| Wayant et al. 2017 | 53 | 109 | 0.49 | [0.39; 0.58] |
| Hannink et al. 2013 | 75 | 152 | 0.49 | [0.41; 0.58] |
| Howard et al. 2017 | 91 | 180 | 0.51 | [0.43; 0.58] |
| Smith et al. 2012 | 13 | 25 | 0.52 | [0.31; 0.72] |
| Khaleel et al. 2019 | 106 | 201 | 0.53 | [0.46; 0.60] |
| Rongen et al. 2016 | 14 | 26 | 0.54 | [0.33; 0.73] |
| Pinto et al. 2013 | 34 | 62 | 0.55 | [0.42; 0.68] |
| Rosati et al. 2016 | 11 | 20 | 0.55 | [0.32; 0.77] |
| Sendyk et al. 2019 | 27 | 49 | 0.55 | [0.40; 0.69] |
| Wandalkar et al. 2017 | 41 | 74 | 0.55 | [0.43; 0.67] |
| Azar et al. 2019 | 48 | 85 | 0.56 | [0.45; 0.67] |
| Hildenbrand et al. 2019 | 16 | 28 | 0.57 | [0.37; 0.76] |
| Bradley et al. 2017 | 8 | 13 | 0.62 | [0.32; 0.86] |
| Chan et al. 2004 | 51 | 82 | 0.62 | [0.51; 0.73] |
| Vedula et al. 2009 | 8 | 12 | 0.67 | [0.35; 0.90] |
| Ludwig et al. 2019 | 20 | 30 | 0.67 | [0.47; 0.83] |
| Nankervis et al. 2012 | 13 | 18 | 0.72 | [0.47; 0.90] |
| **Common effect model** | **2048** | **6314** | **0.32** | **[0.31; 0.34]** |
| **Random effects model** | | | **0.33** | **[0.29; 0.37]** |
| **Prediction interval** | | | | **[0.10; 0.68]** |

Heterogeneity: $I^2 = 86\%$, $\tau^2 = 0.5407$, $p < 0.01$

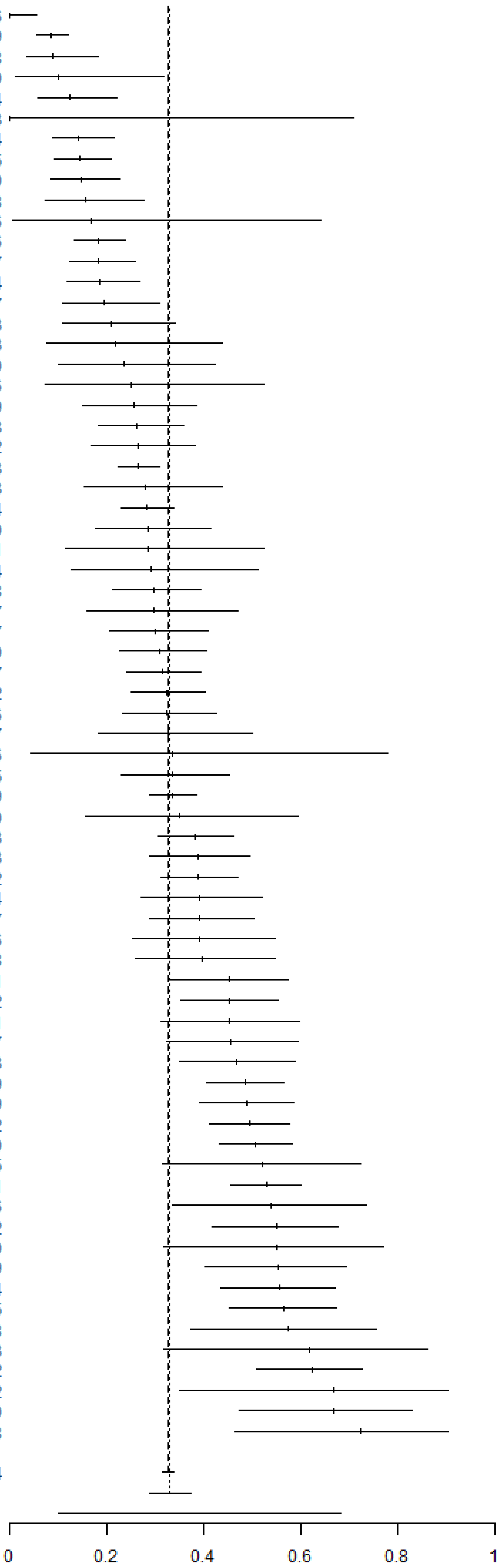


Supplementary Figure F2. Any primary outcome discrepancy.

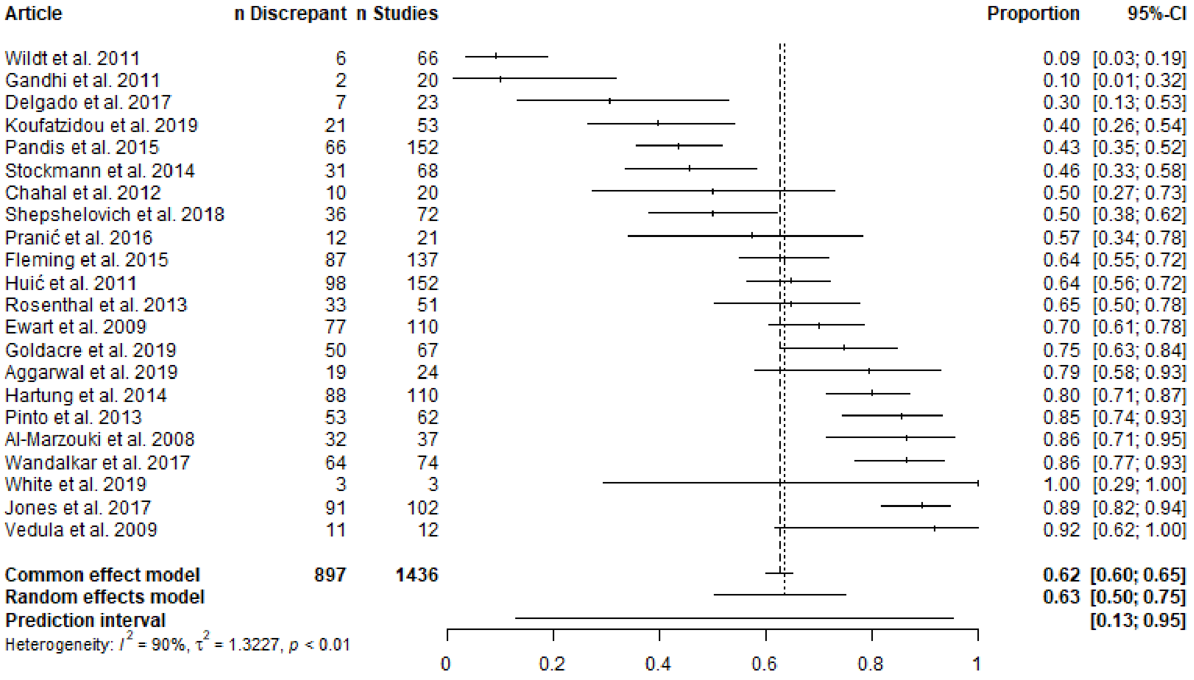


Supplementary Figure F3. Any secondary outcome discrepancy.

| Article | n Discrepant | n Studies | Proportion | 95%-CI |
| --- | --- | --- | --- | --- |
| Sendyk et al. 2019 | 0 | 49 | 0.00 | [0.00; 0.07] |
| Norris et al. 2014 | 0 | 36 | 0.00 | [0.00; 0.10] |
| Chen et al. 2019 | 6 | 389 | 0.02 | [0.01; 0.03] |
| Huić et al. 2011 | 3 | 152 | 0.02 | [0.00; 0.06] |
| Gandhi et al. 2011 | 0 | 20 | 0.00 | [0.00; 0.17] |
| Walker et al. 2014 | 2 | 75 | 0.03 | [0.00; 0.09] |
| Stockmann et al. 2014 | 2 | 68 | 0.03 | [0.00; 0.10] |
| Zhang et al. 2017 | 11 | 299 | 0.04 | [0.02; 0.06] |
| Mathieu et al. 2009 | 6 | 147 | 0.04 | [0.02; 0.09] |
| Aggarwal et al. 2019 | 1 | 24 | 0.04 | [0.00; 0.21] |
| Fleming et al. 2015 | 6 | 137 | 0.04 | [0.02; 0.09] |
| Pranić et al. 2016 | 1 | 21 | 0.05 | [0.00; 0.24] |
| Krsticevic et al. 2019 | 8 | 159 | 0.05 | [0.02; 0.10] |
| Hannink et al. 2013 | 8 | 152 | 0.05 | [0.02; 0.10] |
| Korevaar et al. 2014 | 7 | 131 | 0.05 | [0.02; 0.11] |
| Al-Marzouki et al. 2008 | 2 | 37 | 0.05 | [0.01; 0.18] |
| Ewart et al. 2009 | 6 | 110 | 0.05 | [0.02; 0.11] |
| Howard et al. 2017 | 10 | 180 | 0.06 | [0.03; 0.10] |
| Killeen et al. 2014 | 6 | 108 | 0.06 | [0.02; 0.12] |
| Shepshelovich et al. 2018 | 4 | 72 | 0.06 | [0.02; 0.14] |
| Bonnot et al. 2016 | 5 | 87 | 0.06 | [0.02; 0.13] |
| Li et al. 2013 | 9 | 155 | 0.06 | [0.03; 0.11] |
| Calméjane et al. 2018 | 17 | 274 | 0.06 | [0.04; 0.10] |
| Smith et al. 2013 | 6 | 87 | 0.07 | [0.03; 0.14] |
| Hartung et al. 2014 | 8 | 110 | 0.07 | [0.03; 0.14] |
| Shepshelovich et al. 2017 | 13 | 163 | 0.08 | [0.04; 0.13] |
| Smith et al. 2012 | 2 | 25 | 0.08 | [0.01; 0.26] |
| Wiebe et al. 2017 | 6 | 73 | 0.08 | [0.03; 0.17] |
| Scott et al. 2015 | 5 | 60 | 0.08 | [0.03; 0.18] |
| Chan et al. 2017 | 5 | 58 | 0.09 | [0.03; 0.19] |
| Shinohara et al. 2015 | 4 | 43 | 0.09 | [0.03; 0.22] |
| Rosati et al. 2016 | 2 | 20 | 0.10 | [0.01; 0.32] |
| Jones et al. 2018 | 11 | 103 | 0.11 | [0.05; 0.18] |
| Hildenbrand et al. 2019 | 3 | 28 | 0.11 | [0.02; 0.28] |
| Pandis et al. 2015 | 17 | 152 | 0.11 | [0.07; 0.17] |
| Riehm et al. 2015 | 0 | 3 | 0.00 | [0.00; 0.71] |
| White et al. 2019 | 0 | 3 | 0.00 | [0.00; 0.71] |
| Jones et al. 2017 | 16 | 102 | 0.16 | [0.09; 0.24] |
| Vedula et al. 2009 | 2 | 12 | 0.17 | [0.02; 0.48] |
| Jones et al. 2012 | 10 | 57 | 0.18 | [0.09; 0.30] |
| Chahal et al. 2012 | 4 | 20 | 0.20 | [0.06; 0.44] |
| Su et al. 2015 | 15 | 71 | 0.21 | [0.12; 0.32] |
| Rosenthal et al. 2013 | 11 | 51 | 0.22 | [0.11; 0.35] |
| Dwan et al. 2013 | 10 | 46 | 0.22 | [0.11; 0.36] |
| Chan et al. 2004 | 11 | 48 | 0.23 | [0.12; 0.37] |
| Tricco et al. 2016 | 22 | 96 | 0.23 | [0.15; 0.33] |
| Bradley et al. 2017 | 3 | 13 | 0.23 | [0.05; 0.54] |
| Won et al. 2019 | 16 | 64 | 0.25 | [0.15; 0.37] |
| Pinto et al. 2013 | 17 | 62 | 0.27 | [0.17; 0.40] |
| Chan et al. 2004 | 26 | 82 | 0.32 | [0.22; 0.43] |
| Rongen et al. 2016 | 9 | 26 | 0.35 | [0.17; 0.56] |
| **Common effect model** | **374** | **4560** | **0.08** | **[0.07; 0.09]** |
| **Random effects model** | | | **0.08** | **[0.06; 0.10]** |
| **Prediction interval** | | | | **[0.02; 0.31]** |

Heterogeneity: $I^2 = 79\%$, $\tau^2 = 0.6630$, $p < 0.01$

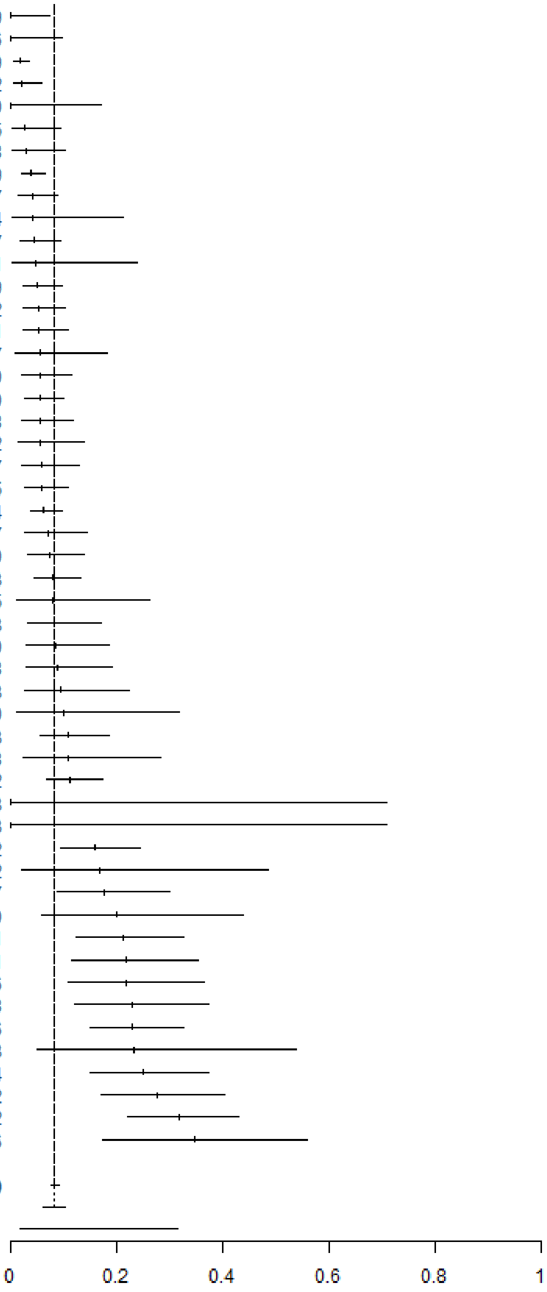


Supplementary Figure F4. Primary outcome demoted to secondary outcome.

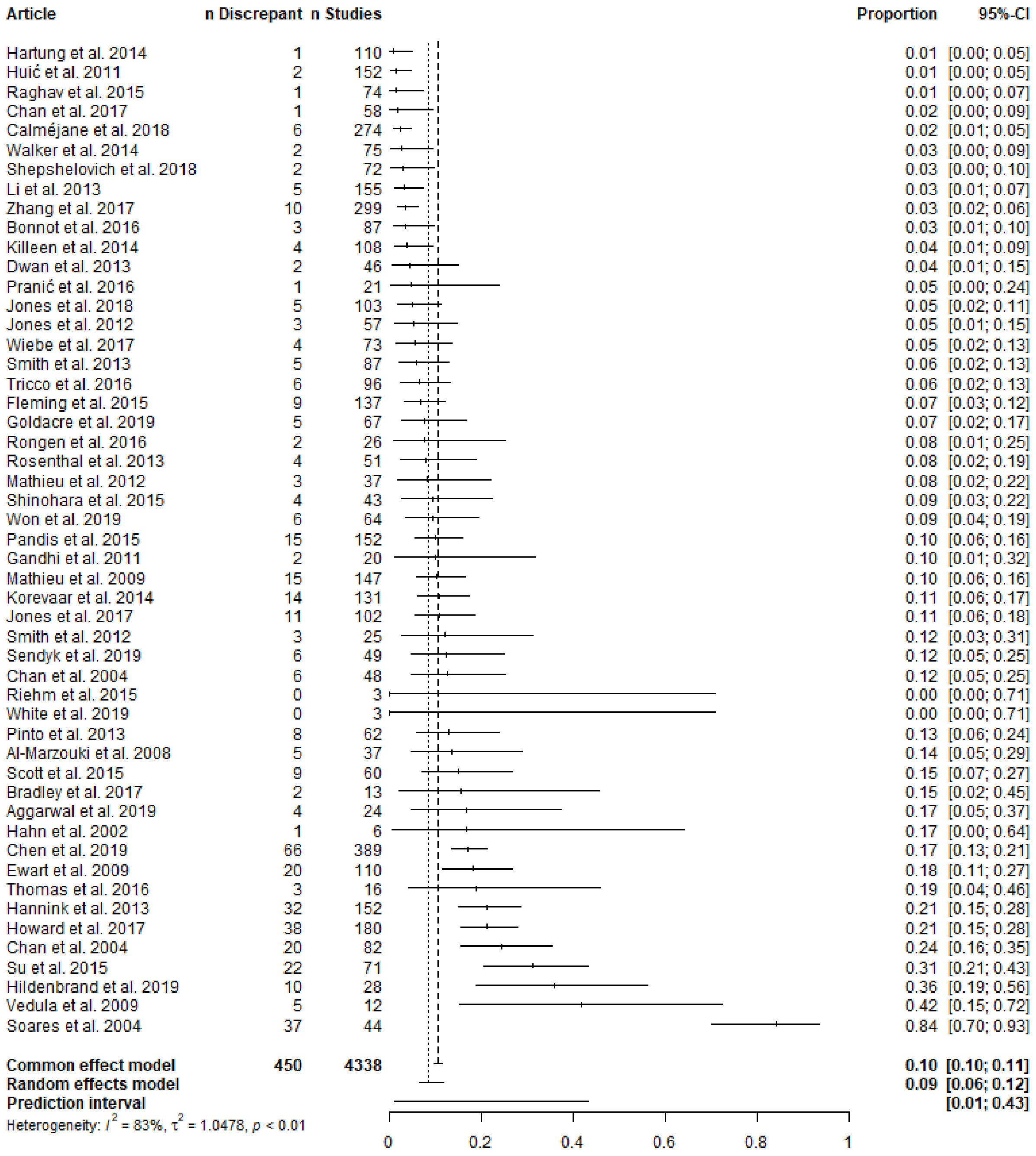


Supplementary Figure F5. Primary outcome omitted.

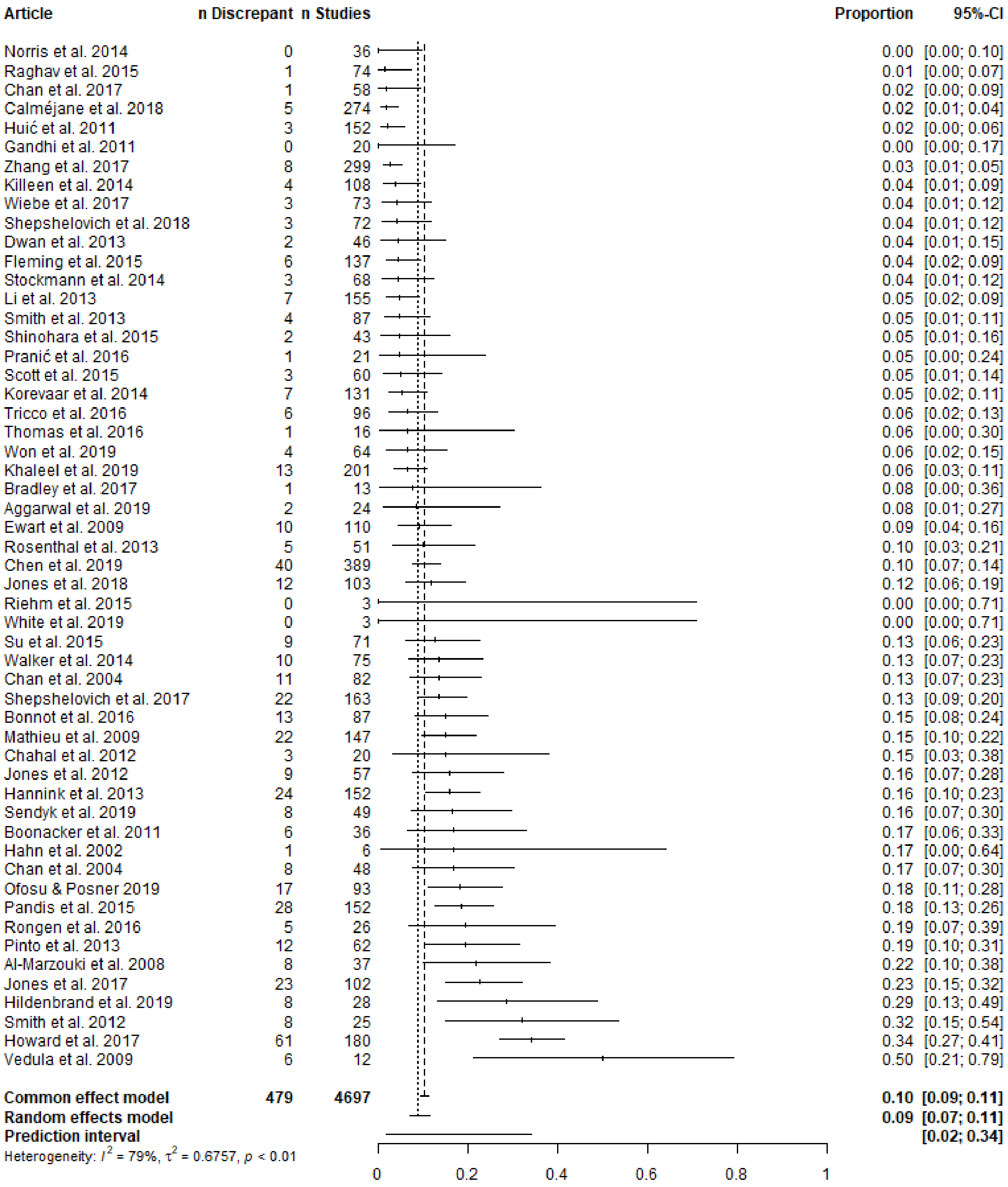


Supplementary Figure F6. Primary outcome added.

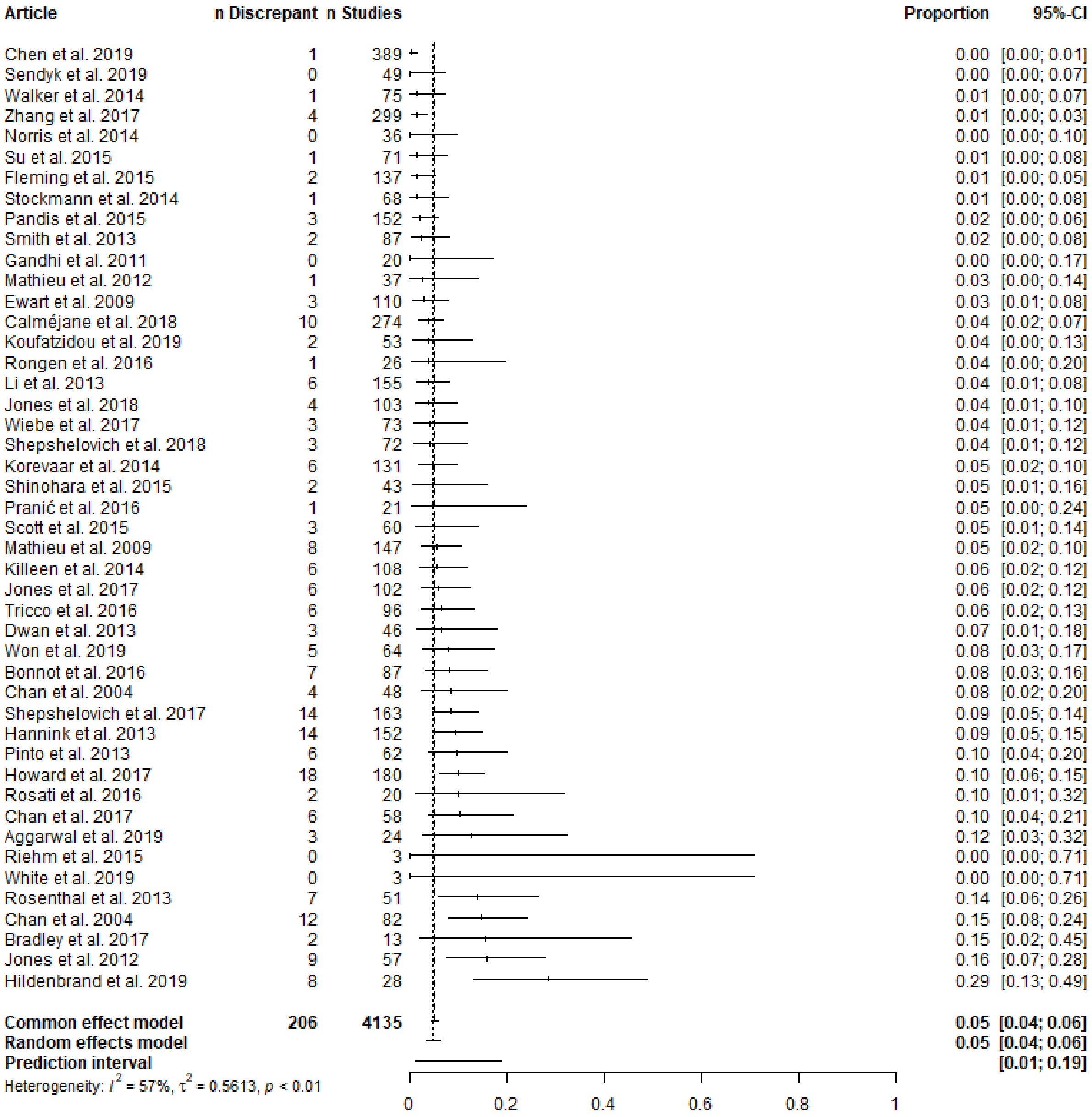


Supplementary Figure F7. Secondary outcome promoted to primary outcome.

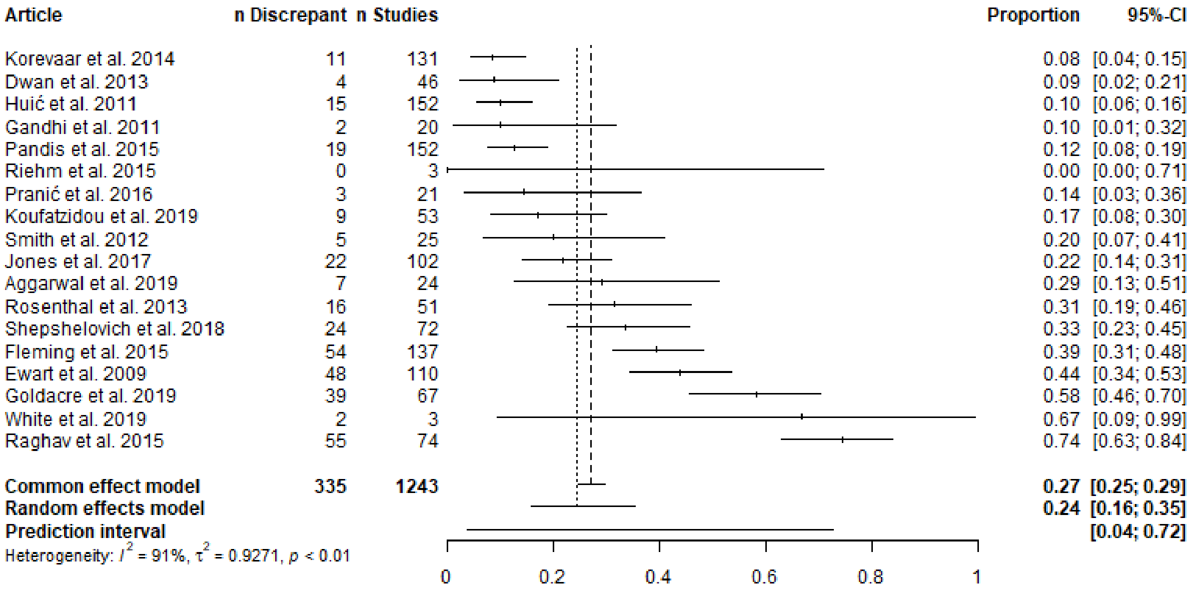


Supplementary Figure F8. Secondary outcome omitted.

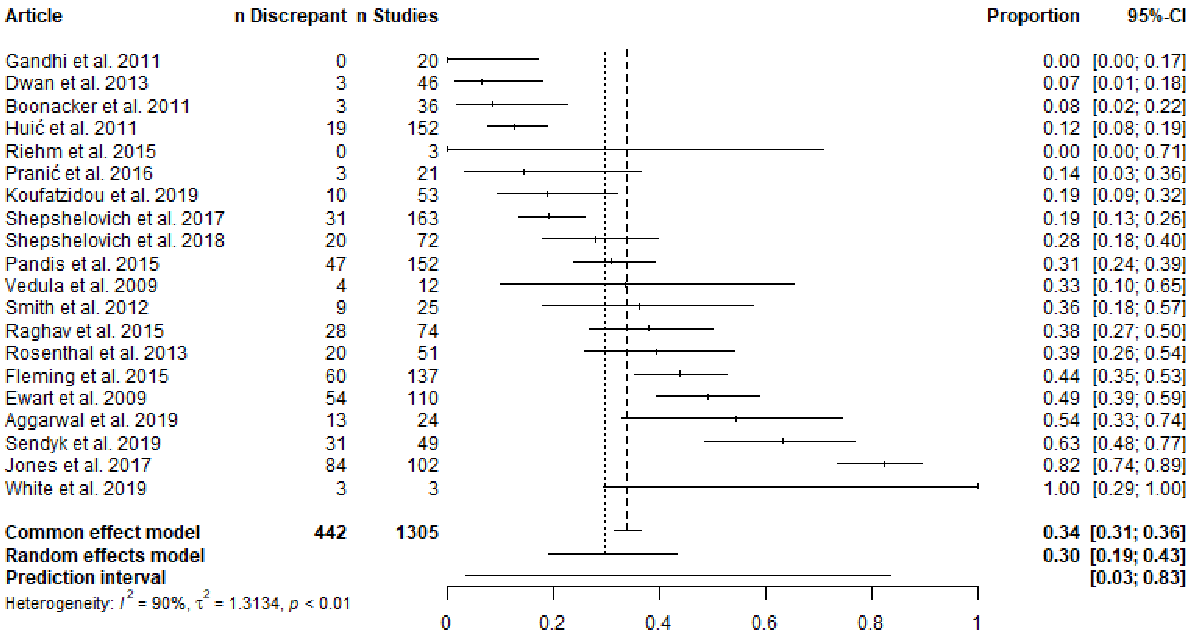


Supplementary Figure F9. Secondary outcome added.

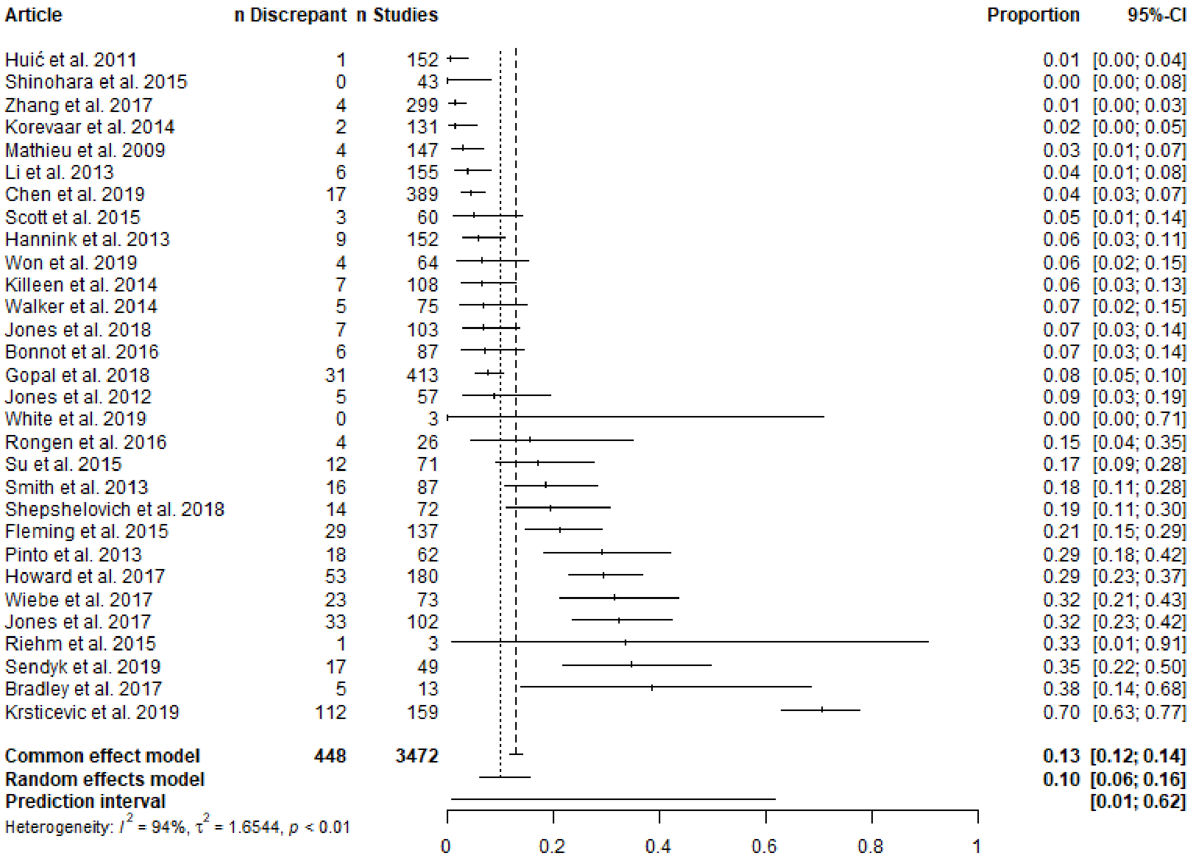


Supplementary Figure F10. Timing of outcome measurement changed.

#### G. Parameters potentially related to discrepancies

##### Disclosure of discrepancies

Some publications disclose the discrepancies between their registration or study protocol and publication. Of the articles we reviewed, k = 8 state that none of the publications disclose their primary outcome discrepancies, k = 13 state that one or more publication disclosed one or more primary outcome discrepancy, k = 68 do not report how many publications disclosed their primary outcome discrepancies, and k = 4 included only publications with undisclosed discrepancies in their sample. A random effects meta-analysis of the k = 21 articles that report the number of disclosed primary outcome discrepancies estimates that 4-19% (95% CI) (95% prediction interval: 0.3-74%) of publications with a discrepancy at least mention that a discrepancy is present (see Supplementary Figure G1 for a forest plot). Notably, a mention does not imply that the publication justified that discrepancy (e.g., Pandis et al., 2015). A few articles identified a much higher proportion of publications that disclosed discrepancies compared to the other articles. These articles focused on acupuncture (Su et al., 2015), meta-analyses (Delgado & Delgado, 2017b), and systematic reviews (Dwan, Kirkham, et al., 2013).

In our initial coding form, we identified 19 articles reporting that the studies they assessed disclosed one or more discrepancy and 9 articles reporting that the studies they assessed disclosed no discrepancies (28 articles total). Only 21 of these 28 articles reported the exact number of studies assessed and the number of discrepancies disclosed (or percentages from which we could calculate these values). Supplementary Figure G1 only includes data from those 21 articles.

##### Statistical significance

k = 24 articles checked whether discrepancies favored statistical significance. Of the studies for which favoring statistical significance could be determined, 49-66% (95% CI) (95% prediction interval: 23-86%) of discrepancies favored statistical significance (see Supplementary Figure G2 for a forest plot).

k = 7 articles provided information on the prevalence of discrepancies in relation to statistical significance. Studies with discrepancies may be *less* likely than studies without discrepancies to report a statistically significant result—likelihood ratio: 0.56-1.06 (95% CI) (95% prediction interval: 0.42-1.43) (see Supplementary Figure G3 for a forest plot). Likewise, rearranging the analysis on this same data shows that studies that report a statistically significant finding are *less* likely to contain a discrepancy—likelihood ratio: 0.64-0.99 (95% CI) (95% prediction interval: 0.59-1.06) (see Supplementary Figure G4 for a forest plot). Whereas these findings may appear counterintuitive, a sampling bias may influence these results—in the sense that the incentive to switch outcomes is less apparent when the registered outcome reaches statistical significance.

##### Industry funding

k = 22 articles provided information on the prevalence of discrepancies in relation to the type of funding a study received. A meta-analysis of these articles shows that industry funded studies are perhaps *less* likely to contain discrepancies compared to non-industry funded studies—likelihood ratio: 0.61-0.91 (95% CI) (95% prediction interval: 0.44-1.27) (see Supplementary Figure G5 for a forest plot).

##### Timing of registration

k = 4 articles compared whether prospectively registered studies were more likely to have primary outcome discrepancies compared to retrospectively registered studies. A meta-analysis of these studies did not find a difference—likelihood ratio: 0.46-2.57 (95% CI) (95% prediction interval: 0.13-8.85) (see Supplementary Figure G6 for a forest plot).

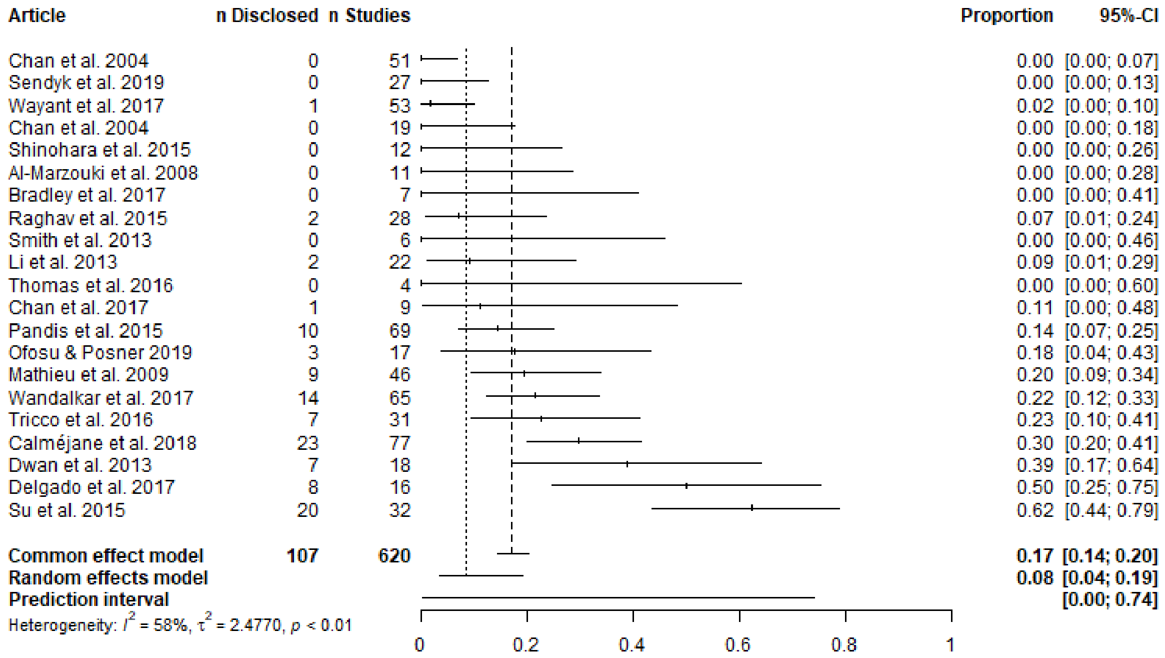


Supplementary Figure G1. Proportion of studies with at least one outcome discrepancy that also disclose an outcome discrepancy.

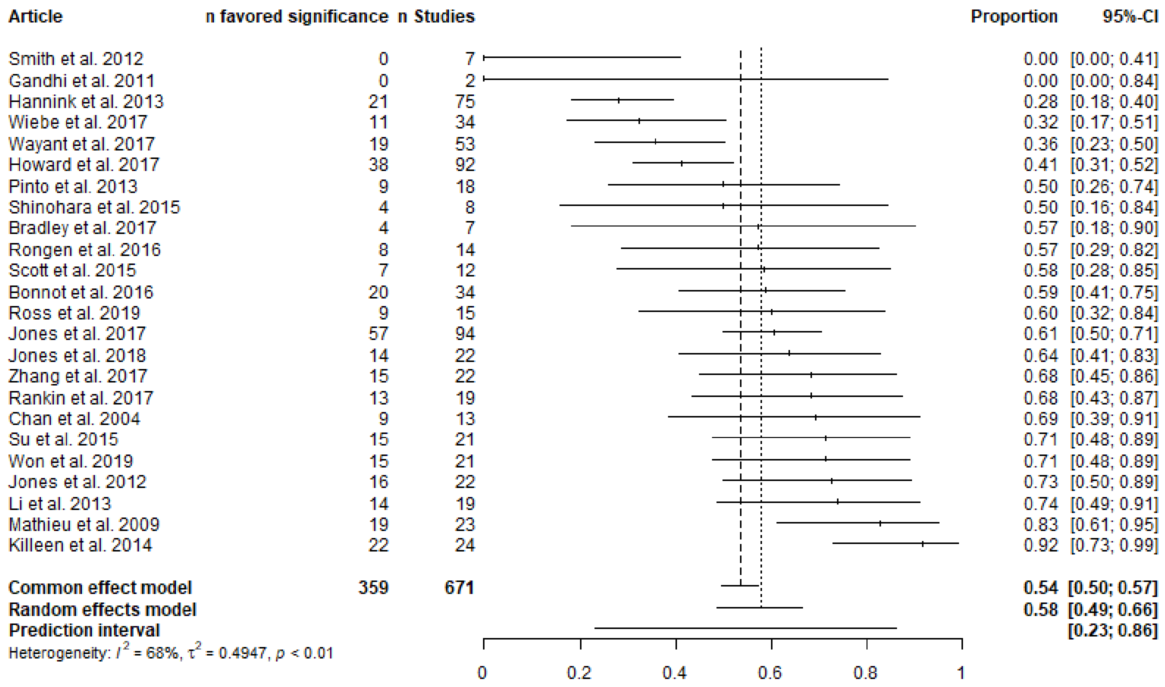


Supplementary Figure G2. Proportion of outcome discrepancies that favor statistically significant results.

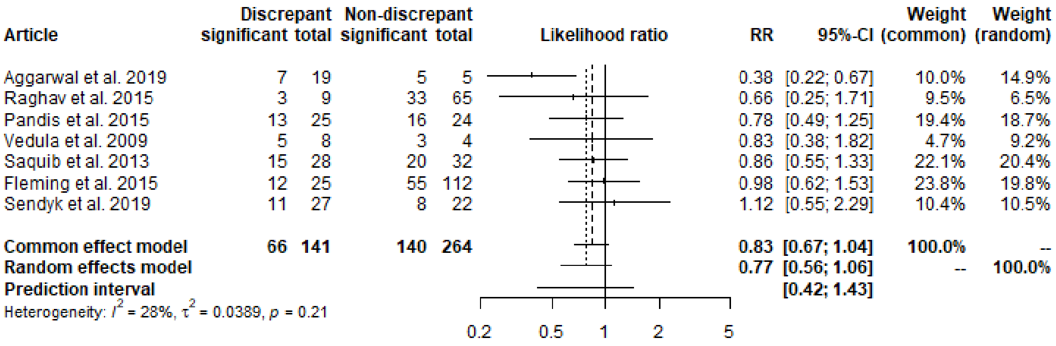


Supplementary Figure G3. Likelihood that a publication with at least one outcome discrepancy, compared to a publication with no outcome discrepancy, reports statistically significant results.

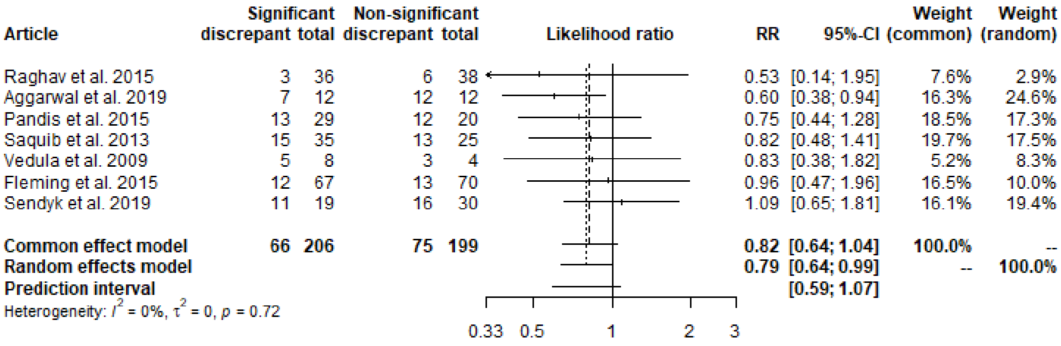


Supplementary Figure G4. Likelihood that a publication reporting a statistically significant result, compared to a publication not reporting a statistically significant results, contains at least one outcome discrepancy.

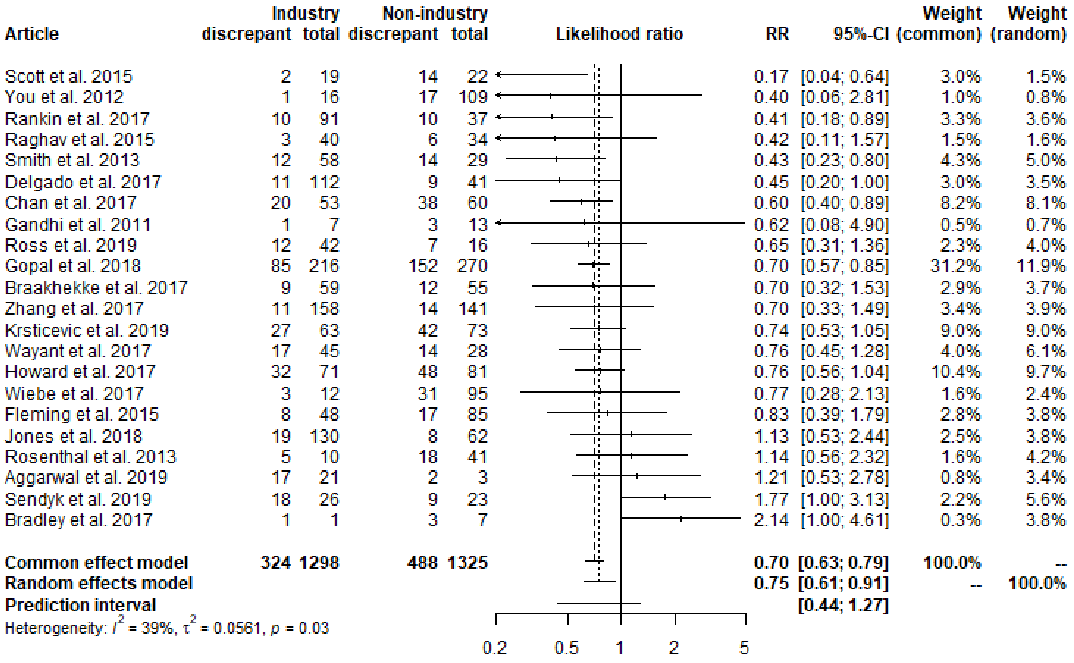


Supplementary Figure G5. Likelihood that a study funded by industry, compared to a study not funded by industry, includes at least one outcome discrepancy.

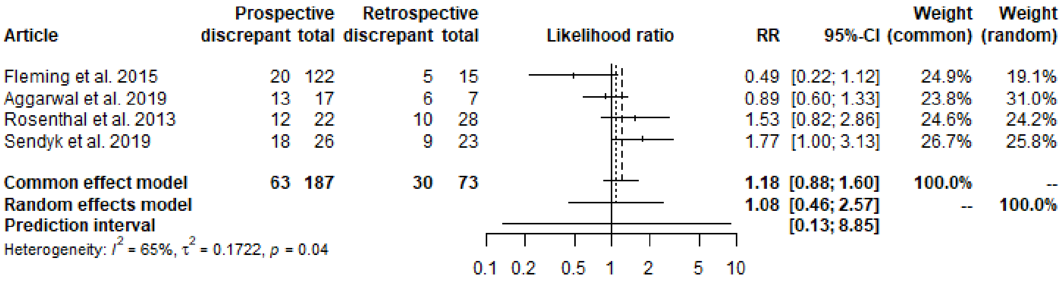


Supplementary Figure G6. Likelihood that a prospectively registered study, compared to a retrospectively registered study, includes at least one primary outcome discrepancy.

#### H. Non-outcome discrepancies

##### Eligibility

Discrepancies in eligibility were sometimes reported separately for inclusion and exclusion criteria. In this case, we took the larger of the two numbers because at least that many studies had a discrepancy in eligibility. Other definitions such as a "change in study population" were also used. While our meta-analysis (k = 15) estimates that 25-57% (95% CI) (95% prediction interval: 5-90%) of publications have at least one discrepancy in eligibility criteria, a broader definition of eligibility would yield an estimate in the higher end of this interval.

##### Sample size

Of the k = 25 articles that report on discrepancies in sample size in our meta-analysis, two required a sample size difference of at least 20%, 7 required a sample size difference of at least 10%, one required a sample size difference of at least 6%, and 15 had no mention of a minimum magnitude of change in sample size. A few articles reported both any discrepancy in sample size and discrepancies surpassing a specified magnitude. In these cases, we counted the discrepancies that reached the largest magnitude cut-off identified in the article. An estimated 26-44% (95% CI) (95% prediction interval: 8-78%) of studies had discrepancies in their sample size.

##### Randomization and blinding

Articles varied greatly in the way they counted discrepancies in randomization. One article surveyed for "a general statement about randomization" (Rosenthal & Dwan, 2013); one counted discrepancies in randomization if there was a change in the number of arms of a study (Sendyk et al., 2019); and one assessed only three studies with generated allocation sequences (Su et al., 2015). These articles found few discrepancies. Other articles counted studies that did not report randomization (Soares et al., 2004) or checked for discrepancies in the randomization procedure (Fleming et al., 2015). These articles found many discrepancies. Few studies include details about randomization in both their registry and publication (Fleming et al., 2015).

Only three of the k = 89 articles we reviewed reported discrepancies in blinding by checking who was blinded (Fleming et al., 2015; Rosenthal & Dwan, 2013; Su et al., 2015). Methods and results were less heterogeneous as compared to discrepancies in randomization.

##### Intervention

Seven articles reported on discrepancies in interventions. Three of these articles only counted high-level discrepancies (e.g., the name of the intervention) and found few discrepancies (Korevaar et al., 2014; Rosenthal & Dwan, 2013; Su et al., 2015). Three other articles counted more fine-grained discrepancies in interventions (e.g., dosage, duration of administration) and found that discrepancies were common (Hui'c et al., 2011; Juri'c et al., 2020; Prani'c & Maruši'c, 2016).

##### Study duration

Four articles reported on discrepancies in study duration. Although the definition of a discrepancy was similar among three of the studies, their results varied substantially—between 3% to 65% discrepant (Rosati et al., 2016; Rosenthal & Dwan, 2013; Shepshelovich et al., 2018).

**Analyses and subgroup analyses**

Twelve articles checked for discrepancies in analysis plans in general, and k = 9 checked specifically for discrepancies in subgroup analyses. Very few studies included a general analysis plan, or a subgroup analysis plan, in both their registration and publication (e.g., Fleming et al., 2015; Rosenthal & Dwan, 2013). Some articles we reviewed only evaluated discrepancies for studies that included an analysis in both their registration and publication. Three of the articles with low estimates for analysis discrepancies only checked whether plans to conduct intention-to-treat analyses were followed (Fleming et al., 2015; Rosati et al., 2016; Soares et al., 2004). Other articles checked for discrepancies in the statistical test for the primary outcome (Chan et al., 2008) or in model adjustments (Saquib et al., 2013). If articles evaluated a broader spectrum of discrepancies in the analyses they checked, and if they considered the addition of an analyses as a discrepancy, the resulting point estimate would be in the higher end of this interval: 19-52% (95% CI) (95% prediction interval: 4-86%). Discrepancies in subgroup analyses were more common: 35-93% (95% CI) (95% prediction interval: 2-99.7%).

**Funding and results**

Discrepancies in funding source and reported results are not directly relevant to the main theme of this paper because they do not imply a deviation from a prospectively planned study protocol. We coded for these discrepancies nonetheless, but the intervals are too wide to draw conclusions about their prevalence (Table 4).

### I. Forest plots for all meta-analyses in Table 4 of the manuscript (non-outcome discrepancies)

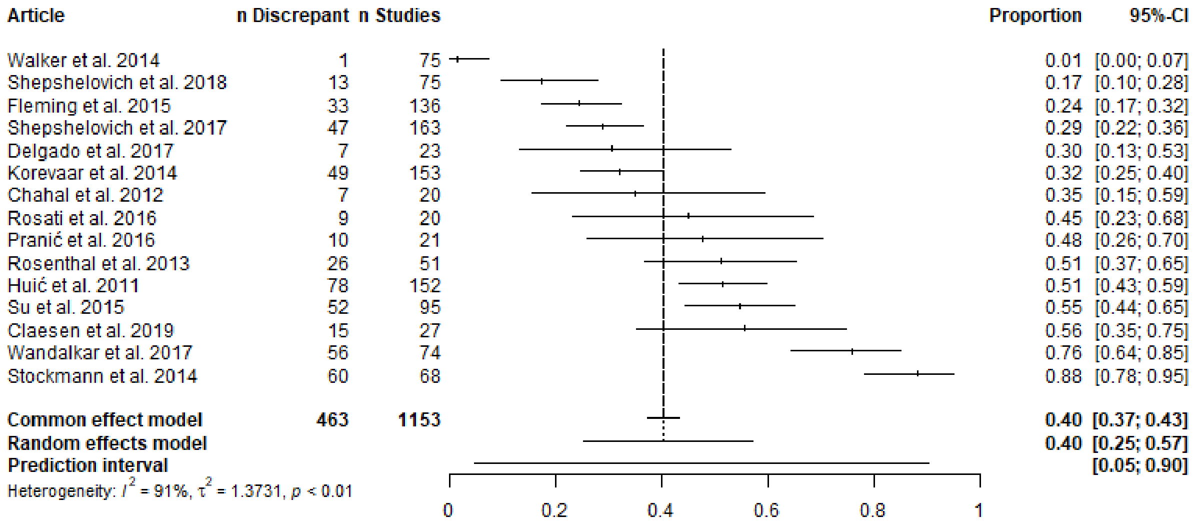


Supplementary Figure I1. Eligibility criteria.

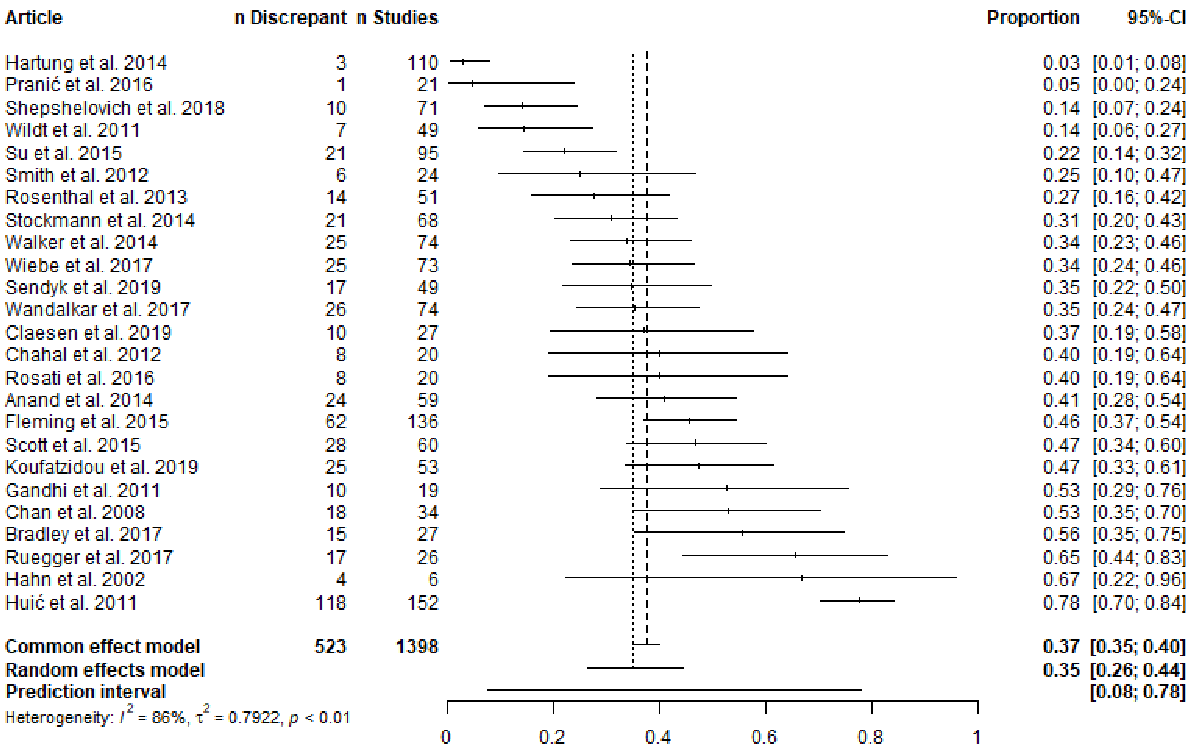


Supplementary Figure I2. Sample size.

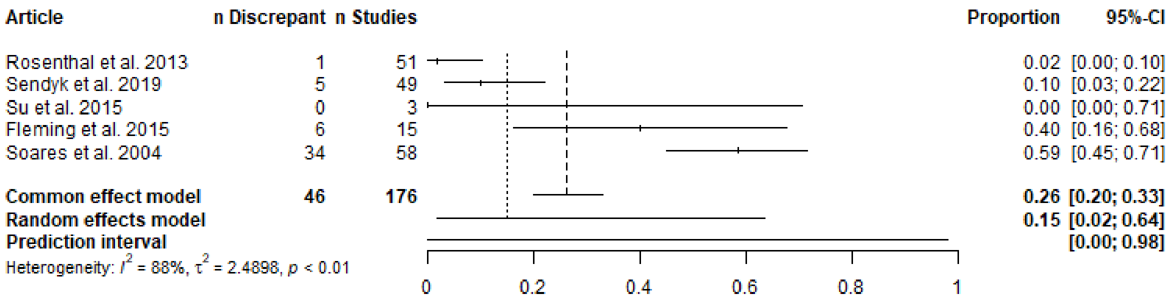


Supplementary Figure I3. Randomization.

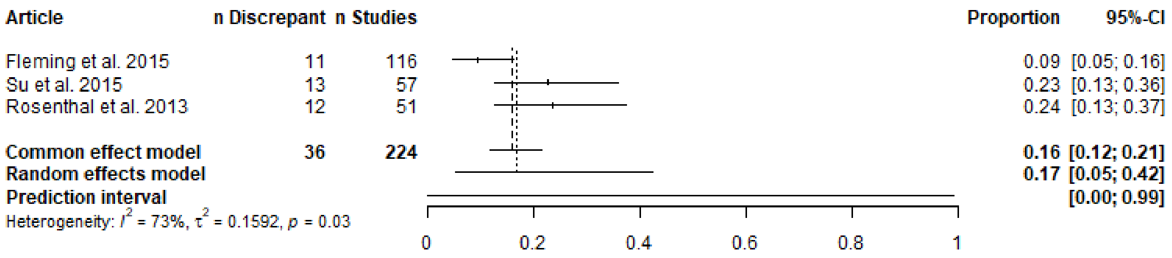


Supplementary Figure I4. Blinding.

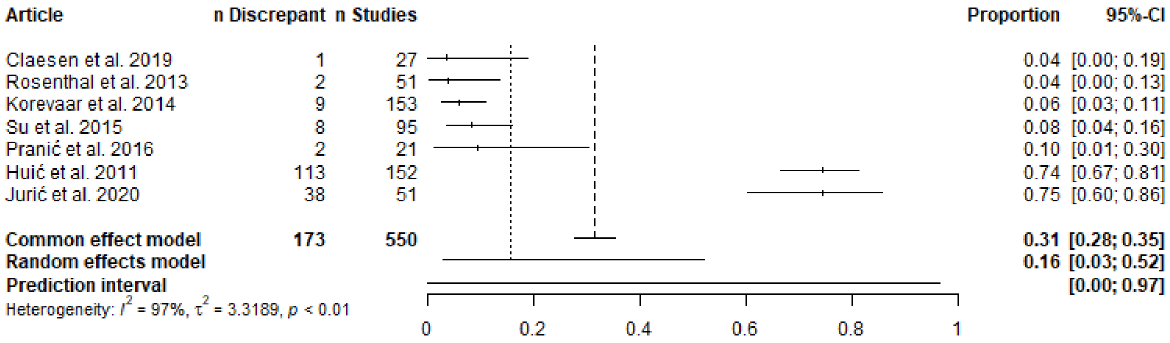


Supplementary Figure I5. Intervention.

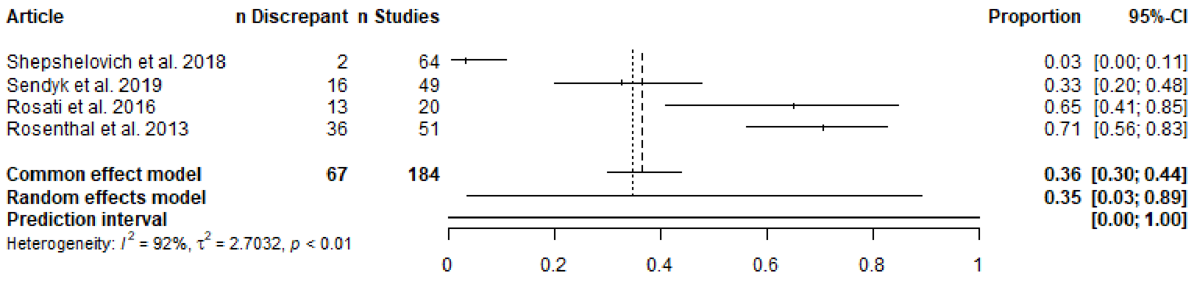


Supplementary Figure I6. Study Duration.

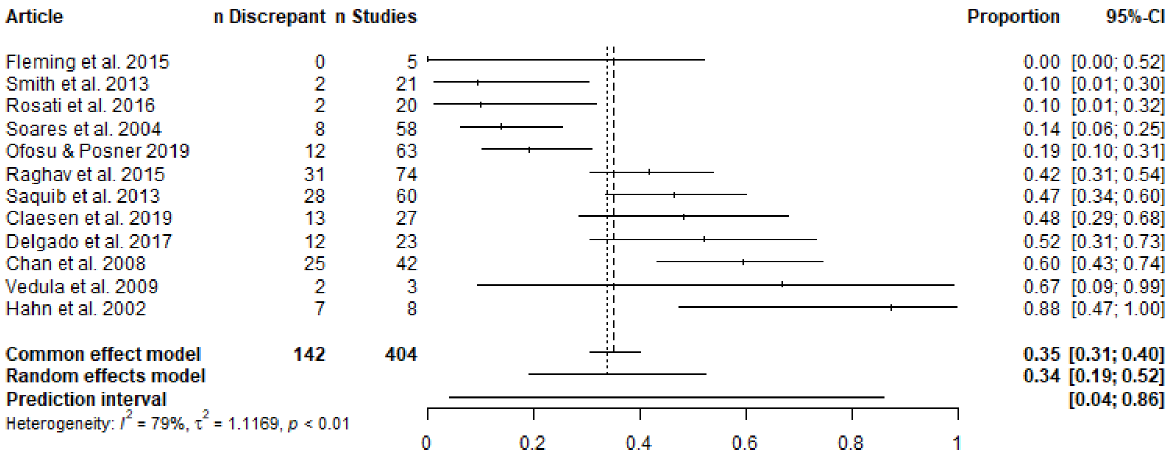


Supplementary Figure I7. Analyses.

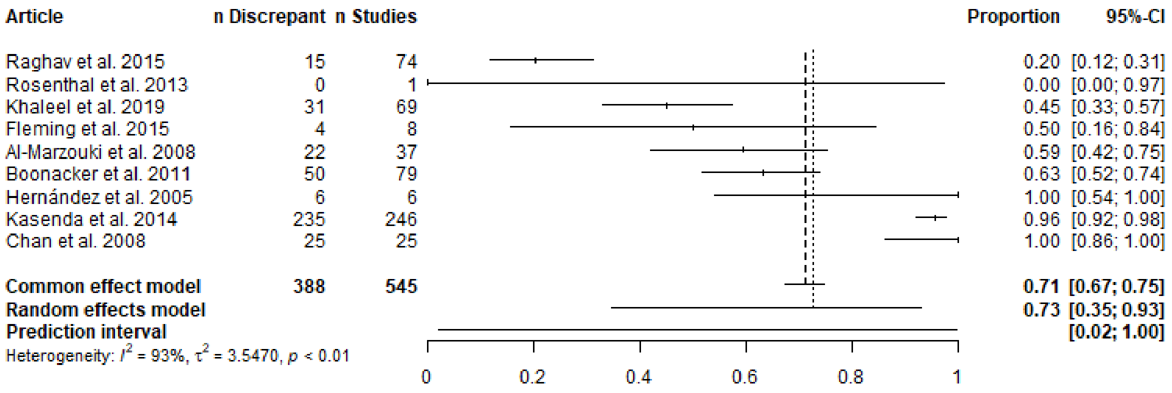


Supplementary Figure I8. Subgroup analyses.

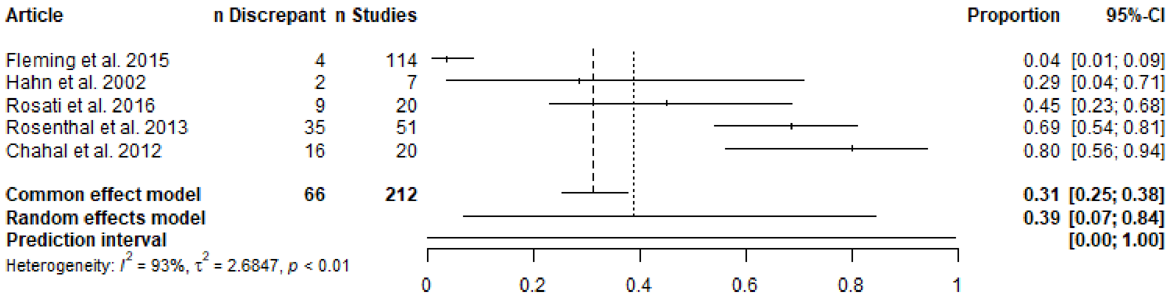


Supplementary Figure I9. Funding.

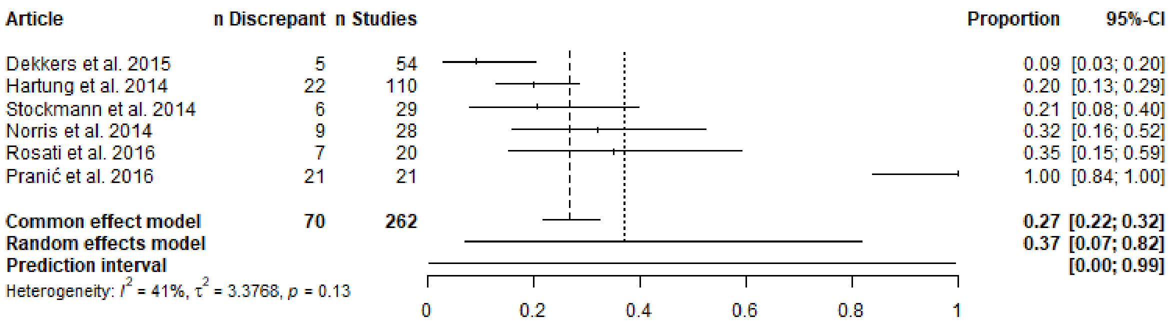


Supplementary Figure I10. Results.

#### J. Gaps in the literature

In addition to the quantitative data presented thus far, our reviewers took qualitative notes. These included a conclusion from each article, details regarding the source used to identify studies and the sub-disciplines they surveyed, and other *ad hoc* observations. One reviewer (RTT) extracted themes related to discrepancies and identified gaps in the literature using narrative synthesis. This process revealed several issues parallel to discrepancies: many studies are never registered, registrations are often imprecise, they rarely include information on analysis plans, and additional initiatives or enforcement may be necessary to improve registration in practice (see Supplementary Table J1 for a list of themes and article quotes).

We further identified at least four gaps in the literature: (1) discrepancy prevalence in fields other than clinical research, (2) discrepancy prevalence in a representative sample across clinical disciplines, (3) the level of detail in registrations, and (4) interventions that attempt to reduce undisclosed discrepancies.

Only one preprint surveyed each of psychology and economics, and no other article assessed preclinical or animal research, or sourced registrations from the Open Science Framework, AsPredicted.org, the American Economic Association (AEA) and Evidence in Governance and Politics (EGAP) registration platform, or the Registry for International Development Impact Evaluations (RIDIE). Notably, registration standards differ across disciplines and this necessitates different methods for identifying discrepancies. For example, pre-analysis plans on EGAP often contain over 10 hypotheses (Ofosu & Posner, 2019) and clinicaltrials.gov does not contain a question regarding an analysis plan.

Many articles we reviewed restricted their search to high-impact factor journals or a specific clinical discipline. Randomly sampling registrations and publications from the broader clinical literature could better estimate the prevalence of discrepancies at the level where some interventions could be applied (e.g., an ICMJE policy). With a sufficient sample size, random sampling could allow comparisons among clinical disciplines and the potential to identify best practices.

Most articles did not identify the level of specificity with which an outcome was registered. For example, outcomes can be specified at the level of domain (e.g., anxiety), specific measurement (e.g., Hamilton Anxiety Rating Scale), specific metric (e.g. change from baseline), and method of aggregation (e.g., proportion of participants with decrease >50%) (example taken from Deborah A. Zarin et al. (2011)). Outcomes registered with low specificity allow for a greater range of researcher degrees of freedom. Future research efforts can also benefit from identifying whether the registrations were prospective, which version of the registration was being assessed, and if discrepancies were disclosed. This information would help reveal how problematic the discrepancies are.

Finally, nearly all the articles focused on documenting discrepancies. Future efforts may be more fruitful if they attempt to understand the causes of discrepancies and seek to develop and evaluate solutions to the issue (e.g., following the framework for meta-research presented by Hardwicke, Serghiou, et al. (2020)).

Supplementary Table J1. Additional themes related to registration and discrepancies

| Theme | Quotes |
|---|---|
| Many studies are never registered | "The more concerning issue, however, is the lack of registration or provision of registration number for randomized controlled trials within these journals." (Wiebe et al., 2017)<br>"Only 16 per cent of the published orthodontic RCTs had been registered." (Koufatzidou et al., 2019) |
| Registrations are imprecise | "Our results suggest that [selective outcome reporting] might well be substantial; however, the bias can only be broadly identified as protocols are not sufficiently precise." (Hahn et al., 2002)<br>"Only 5 out of the 109 trials (5%) provided enough information for us to be confident that the outcomes reported in the published trial were consistent with the original registration" (Nankervis et al., 2012) |
| Consumers of research should be aware of outcome switching | "If clinicians base their decisions on evidence distorted by primary outcome switching, patient care could be negatively affected" (C. W. Jones et al., 2017)<br>"Readers of medical literature should not assume that selective outcome reporting is not present, even if the report has been published in a high IF journal." (Shinohara et al., 2015) |
| Registration should be mandated or enforced | "there is insufficient quality control of data by data providers, ClinicalTrials.gov administrators, and journal editors, emphasizing much-needed enforcement of complete descriptions of drug interventions." (Jurić et al., 2020)<br>"Journal editors, regulators, research ethics committees, funders, and sponsors should implement policies mandating prospective registration for all clinical trials" (Chan et al., 2017)<br>"Reporting completeness and consistency were significantly better after July 2005. The ICMJE requirement for mandatory registration was associated with significant improvement in reporting quality in infectious diseases trials." (Shepshelovich et al., 2017) |
| Additional initiatives are needed to reduce reporting discrepancies | "Efforts to prevent nonregistration of protocols and selective reporting are needed." (Krsticevic et al., 2019)<br>"To foster credible evidence-based medicine, additional initiatives are needed to minimize selective reporting." (Raghav et al., 2015)<br>"Novel approaches are required to address this problem." (Fleming et al., 2015) |
| Analysis plans should be registered | "More immediate and specific remedies for the deficiencies of diet trial registries include (1) posting a detailed final statistical analysis plan before unmasking random group assignments or beginning data analyses as a minimum quality criterion..." (Ludwig et al., 2019)<br>"Definitive judgments regarding credibility of claimed subgroup effects are not possible without access to protocols and analysis plans of randomised controlled trials." (Kasenda et al., 2014) |
